# Reconsider photoplethysmogram signal quality assessment in the free living environment

**DOI:** 10.1101/2024.02.26.24303386

**Authors:** Yan-Wei Su, Chia-Cheng Hao, Gi-Ren Liu, Yuan-Chung Sheu, Hau-Tieng Wu

## Abstract

**Background:** Assessing signal quality is crucial for biomedical signal processing, yet a precise mathematical model for defining signal quality is often lacking, posing challenges for experts in labeling signal qualities. The situation is even worse in the free living environment.

**Method:** We propose to model a PPG signal by the adaptive non-harmonic model (ANHM) and apply a decomposition algorithm to explore its structure, based on which we advocate a reconsideration of the concept of signal quality.

**Result:** We demonstrate the necessity of this reconsideration and highlight the relationship between signal quality and signal decomposition with examples recorded from the free living environment. We also demonstrate that relying on mean and instantaneous heart rates derived from PPG signals labeled as high quality by experts without proper reconsideration might be problematic.

**Conclusion:** A new method, distinct from visually inspecting the raw PPG signal to assess its quality, is needed. Our proposed ANHM model, combined with advanced signal processing tools, shows potential for establishing a systematic signal decomposition based signal quality assessment model.

## 1. Introduction

Various methods exist for assessing the quality of a photoplethysmogram (PPG) signal [16]. Fundamental criteria, such as the presence of clear pulse peaks, are essential for basic signal quality [23], facilitating heart rate (HR) extraction through peak detection algorithms [6]. Diagnostic quality demands clean pulse waveforms, or cardiac components, with visible systolic and diastolic waves [18]. Additional considerations include pulse amplitude and width consistency with adjacent pulses, and adherence to typical PPG pulse morphology (primary and secondary peaks, troughs, pulse amplitude, width, etc.) [33]. Alternatively, simultaneous recording of other signals, as demonstrated in [20], can be employed to define quality by comparing PPG-derived HR to electrocardiogram (ECG)-derived HR. To our knowledge, generally, experts seem to rely on the visibility of the cardiac component to label the quality of a PPG segment. To automatically quantify PPG quality, signal processing techniques are needed. This can be achieved through time-domain, frequency-domain, or hybrid approaches, guided by predefined rules or machine learning techniques. See [23, 16] for a review of available tools.

Despite implicitly consented PPG signal quality criteria among experts and numerous proposed PPG signal quality assessments (SQA) algorithms, there is, to our knowledge, no paper precisely defining what PPG signal quality really means with a mathematical model, particularly in the context of subjects moving in a free-living environment. As a result, PPG signals with multiple components, especially involving moving rhythms with the movement rhythm close to HR, may be considered of low or high quality due to the presence of “non-cardiac” artifacts depending on the potential phase synchronization between heart rate and moving rhythm. See Figure 1 for an example of a “low quality” PPG and Figure 4 for a “high quality” PPG, both are recorded during exercise, and experts’ label. Consider that various sources of “non-cardiac” artifacts can significantly alter the PPG waveform, such as sudden breathing changes, body movements, or talking. In the pursuit of *extracting cardiac information* from the PPG component, it is crucial to minimize irrelevant factors like respiration and motion artifacts, especially in a free-living environment. Addressing these artifacts is a challenging task. Since the respiration is usually of much lower frequency, it usually can be suppressed by a high-pass filter. Several algorithms have been proposed to reduce motion artifacts [10, 27, 13, 19, 26] as a pre-processing stage for robust heart rate detection. We could view some of these motion artifact removal algorithms as a special case of the general signal decomposition mission. To our knowledge, the relationship between signal decomposition and signal quality has not been well studied.

**Figure 1.**
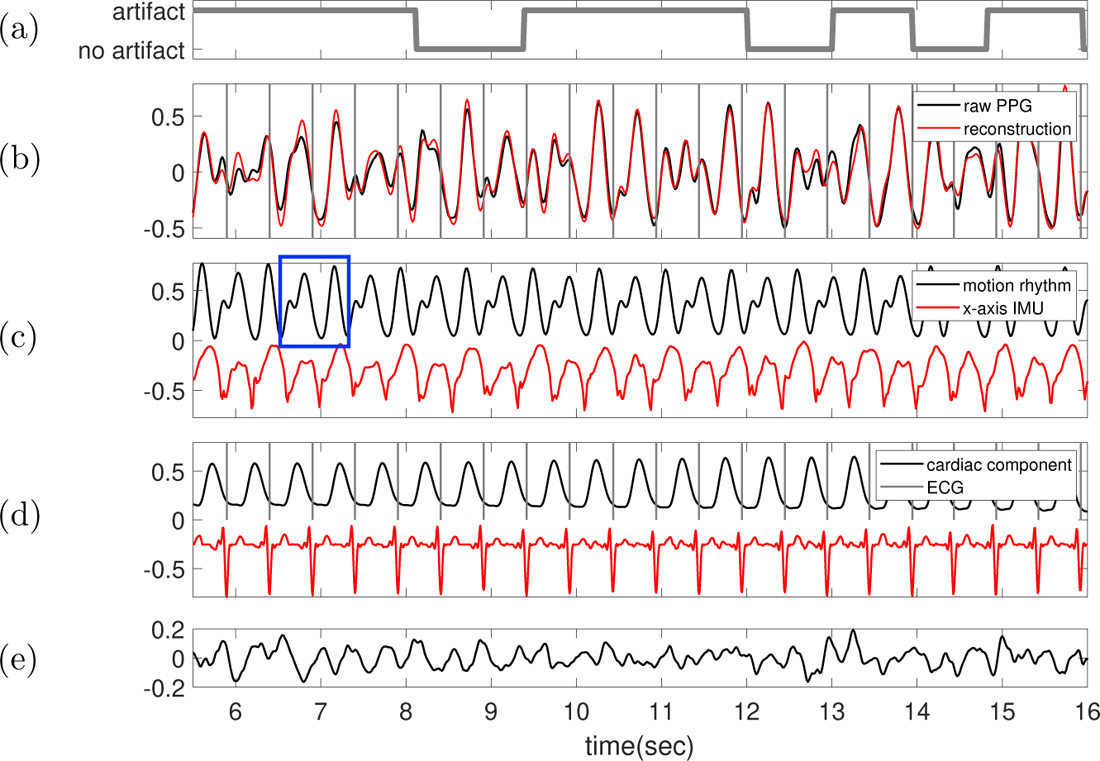
The 65.5th to the 76th second segment of the 6th subject in the TROIKA dataset, who is running at the speed of 6 kilometer per hour during this segment. **(a)** Label sequence (grey line) and the prediction result by the proposed SQA model (red-dashed line). **(b)** The raw PPG signal is shown in black, and the summation of the decomposed motion rhythm and cardiac component is superimposed in red. **(c)** The decomposed motion rhythm (black curve) and the synchronously recorded x-axis accelerometer signal (red curve). **(d)** The decomposed cardiac component (black curve), the synchronously recorded ECG (red curve) and the detected R-peaks (grey lines). **(e)** The difference of the raw PPG signal and the summation of the decomposed motion rhythm and cardiac component. The decomposition NRMSE is 0.138.

Motivated by the critical role played by the signal quality in biomedical signal processing, we advocate the necessity of reconsidering what it means by the signal quality using PPG, and the necessity for a precise definition of signal quality based on a well defined mathematical model that is oriented by the scientific target. We emphasize the close connection between signal quality definition and the signal decomposition problem, particularly in handling non-sinusoidal oscillatory patterns of respiration and motion in the PPG signal, by modeling the PPG signal with the *adaptive non-harmonic model*(ANHM). For instance, respiration-induced intensity variation (RIIV) and motion rhythm, being non-sinusoidal when exists, may introduce spectral interference with the cardiac component. Armed with this signal decomposition technique, we provide a detailed exploration of potential issues of existing signal quality labeling approach that may arise without a precise signal quality definition.

## 2. Mathematical model and signal decomposition

To model a PPG signal, we indicate some observations. First, due to the heart rate variability, the frequency of the cardiac component is time-varying. Second, the respiration may impact the oscillatory strength of the cardiac component or exist as an additional oscillatory component called RIIV [29]. Motion rhythm might exist if a subject is moving. Third, the trends, or direct currents (DCs), of red and infrared channels encode the oxygen saturation information [12]. Fourth, noise is inevitable. Jointly, we consider the *adaptive non-harmonic model* (ANHM) [34] to model the PPG signal. Fix small constants *E >* 0 and Δ *>* 0. A clean PPG is modeled by

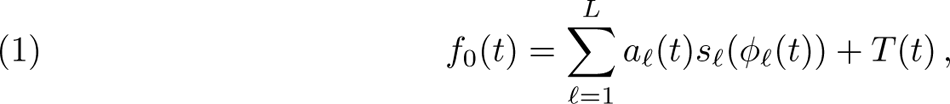

where *a_f_*(*t*)*s_f_*(*φ*(*t*)) is called the *intrinsic model type* (IMT) function fulfilling some conditions listed below and *T* (*·*) is a smooth function so that its Fourier transform *T*^^^ is compactly supported in [*−*Δ, Δ]. For each *f ∈ {*1*, …, L}*, we assume (C1) *φ_f_*(*t*) is a *C*^2^ function, which is called the *phase function* of the *f*-th IMT function; (C2) *φ,* (*t*) *>* 0, which is called the *instantaneous frequency (IF)* of the *f*-th IMT function.

We assume *|φ^,,^*(*t*)*| ≤ Eφ,_f_* (*t*) for all *t_f_ ∈* R; (C3) *a_f_*(*t*) *>* 0 is a *C*^1^ function, which is called the *amplitude modulation (AM)* of the *f*-th IMT function so that *|a,* (*t*)*| ≤ Eφ_f_,* (*t*) for all *t_f_ ∈* R; (C4) *s_f_*(*·*) is a smooth 1-period function with mean 0 and unit *L*^2^ norm so that *|s*^*_f_*(1)*| >* 0, which is called the *wave-shape function* (WSF) of the *f*-th IMT function; (C5) when *L >* 1, we assume *φ_f_,* (*t*) *− φ,_f-1_* (*t*) *≥* Δ for *f* = 2*, …, L* and *φ_1_,* (*t*) *≥* 2Δ.

In this model, *E* quantifies how fast the AM and IF could change from time to time, and Δ quantifies the spectral gap between two consecutive IMT functions and the first IMT function and the trend. The assumptions (C1)-(C5) are imposed to capture the commonly observed PPG behavior. Moreover, the performance of the decomposition algorithm described below is guaranteed under these assumptions. In practice, since we can easily remove the trend component *T* by applying a high-pass filter in the pre-processing step, from now on we assume *T* = 0 to simplify the discussion. A more complicated model could be considered [14], but to avoid distracting from the discussion of signal quality, we are satisfied with this ANHM model. See signals shown in Figures 1 and 4 for examples with *L* = 2. It is difficult to visualize the cardiac oscillatory pattern in the raw PPG signal. Thus, a tool that decomposes the signal into IMT components plays an important role when we want to analyze the oscillatory signal of the type (1).

For signal decomposition, we apply the recently developed algorithm Shape-Adaptive Mode Decomposition (SAMD) [2] to obtain each IMT function. In short, we estimate the IF of each IMT function with a preferred time-frequency (TF) analysis algorithm [7]. In this work we choose the second order synchrosqueezing transform (SST) [4, 21] with the recently developed ridge extraction algorithm, *multiple harmonics ridge detection* (MHRD) [32], to obtain a reliable ridge that represents the IF [5], and hence the phase and AM. SAMD is a nonlinear optimization algorithm that takes the estimated phase and AM of each IMT function to reconstruct each IMT function.

## 3. Reconsider what the signal quality should be

We shall now discuss a critical issue regarding what signal quality should be defined. We demonstrate the issue using signals from the publicly available dataset TROIKA [35] with experts’ annotation from [9]. In the TROIKA dataset, both the PPG signal and triaxial acceleration signal were recorded from the wrist. Subjects performed treadmill running with changing speeds during data collection. The labels assign binary values (1 for “artifact” or “low quality”, and 0 for “clean”, “no artifact” or “high quality”) to each sample point in the segment. The TROIKA dataset was pre-processed by its original author in [35] with bandpass from 0.4 Hz to 5 Hz.^1^ We keep this setting since we do not have an access to the original PPG signal. All the 30-second PPG segments are uniformly sampled at the sampling rate of 64 Hz, and the signal values are normalized to the range [0, 1]. We delineate the label generation process [9]. Binary labels were established by annotators through assessing the correlation between ECG and PPG heartbeats, and examining the regularity of PPG signals to identify artifacts. Two scenarios were considered: (1) If motion is detected in the accelerometer and irregularities align with the accelerometer data, the segment is marked as an artifact. (2) If no motion is detected in the accelerometer, ECG displays normal sinus rhythm, but irregularities are present in the PPG signal, the segment is marked as an artifact. Each signal was annotated by at least one annotator. Initially, fifty 30-second segments were randomly selected and independently annotated by three annotators. Annotations were compared and analyzed, and decisions were made collectively to improve agreement. Subsequent annotations were conducted by a single annotator.

See Figure 1 for an example PPG segment, the 65.5th second to the 76th second segment, from the 6th subject in the TROIKA database [35] (DATA 06 TYPE02.mat), when the subject is running at the speed of 6 kilometer per hour during this segment. The label sequence provided by experts [9] is depicted as the gray curve in the first row. Examining the raw signal in Figure 1(b) may encourage labeling the segment to have low quality due to its irregular oscillation pattern that is visually different from the ordinary PPG signal, and some subsegments are labeled “no artifact” (or high quality) based on visually identifiable “regular” oscillations mimicking cardiac patterns. However, is this label really what we want? To confirm if this label is authentic, we dig into the ground truth by inspecting the simultaneously recorded ECG signal (red curve in Figure 1(d)) and x-axis accelerometer signal (red curve in Figure 1(c)). To enhance the visualization, the R peak locations are superimposed on the second row as gray vertical lines. We find that the seemingly regular subsegments in PPG do not align with the cardiac oscillation. This raises concerns about determining signal quality from the raw PPG signal. While the provided label is not from only reading the raw PPG signal but taking other channels into account, we would like to bring to readers’ attention that the meaning of the signal quality might be reconsidered.

To dig deeper, we find that the PPG segment encompasses not only the cardiac component (shown in Figure 1(d)) but also a strong motion rhythm (shown in Figure 1(c)), where we apply SAMD [2] to get these components. The quality of the decomposition can be visualized by the summation of the decomposed cardiac component and motion rhythm that is superimposed as the red curve on the second row. The normalized root mean squared error (NRMSE) is 0.235. We may consider the decomposed cardiac component of high quality (black curve in Figure 1(d), with R peaks from the ECG shown as vertical gray lines for comparison).

The time-frequency representation (TFR) of the PPG segment determined by SST shown in Figure 2(a) provides additional insights. Examining multiple curves with a non-integer multiple relationship suggests the presence of multiple oscillatory components. Specifically, two nearly parallel lines appear at approximately 2 and 2.6 Hz, associated with the fundamental frequency of the cardiac component (heart rate) and the second harmonic of the motion rhythm, respectively. In other words, the motion rhythm is at about 1.3Hz, but its fundamental component is weak, or even missing, in the PPG signal and we only see the second, third and fourth harmonics indicated by blue arrows. See Figure 2(b) for the TFR of the motion rhythm, which comes from subtracting the cardiac component from the PPG signal, where the pink dashed box indicates the region around 1.3Hz. Also see Figure 3 for a concrete evidence that the motion rhythm is about 1.3Hz, where the TFRs of the simultaneously recorded Inertial measurement unit (IMU) signal analyzed from different aspects are shown. It is important to note that the TFRs vary based on the perspective from which the IMU signal, as a vector-valued function, is analyzed. In the TFR of the magnitude of the IMU signal, curves are visible at 2.6 and 5.2 Hz, but not at 1.3 Hz. However, in the TFR of each axis, a curve appears at 1.3 Hz. This disparity arises from different WSFs associated with how we analyze the IMU signal as a multivariate time series. It is also worth noting that the motion rhythm depends on both the running speed and stride length. On the other hand, the HR gradually increases from 1.99 Hz to 2.11 Hz, while the motion rhythm keeps constant. As a result, the frequencies of the second harmonic of the cardiac component and the third harmonic of motion rhythm depart as time proceeds, which leads to the spectral interference in the first half of the signal indicated by the pink dashed box in Figure 2(a). After removing the cardiac component, we can see three obvious lines indicated by blue arrows in Figure 2(b), which are associated with the second, third and fourth harmonics of the motion rhythm. This fact is reflected in the decomposed motion rhythm shown in Figure 1(c), where we can see a repeating pattern indicated by the blue box, which seems to have two cycles, but these two cycles are actually one thanks to the third harmonic. In this TFR, only two dominant harmonics of the cardiac component indicated by pink arrows are visible, likely due to the application of a strong low-pass filter with a cutoff frequency of 5 Hz to the database. This partially explains why the decomposed cardiac component differs from the typical PPG signal recorded during rest. Overall, through SAMD and TFR, we gain insight into the construction of the seemingly chaotic PPG signal, prompting a reconsideration of signal quality.

**Figure 2.**
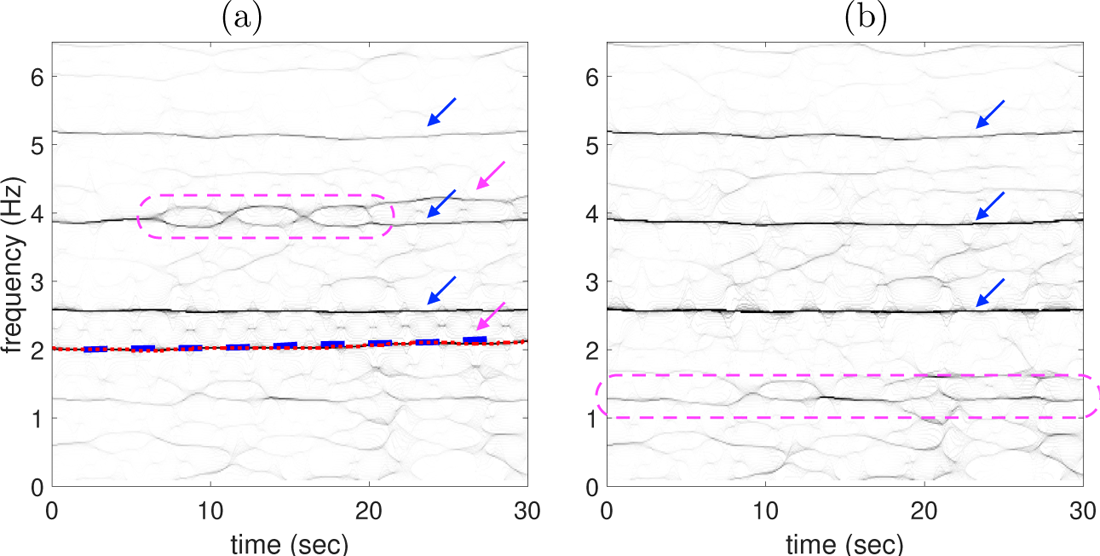
The TFR of the 60th to the 90th second PPG segment of the 6th subject in the TROIKA datasets, who is running at the speed of 6 kilometer per hour during this segment. We show the second-order SST with the Gaussian window with *σ* = 1.5 second. **(a)** The TFR of the original PPG signal, where the blue-dashed line is the ECG-derived IHR, and the red-dotted line is the result of the RD algorithm. **(b)** The TFR of the PPG signal after removing the cardiac component.

**Figure 3.**
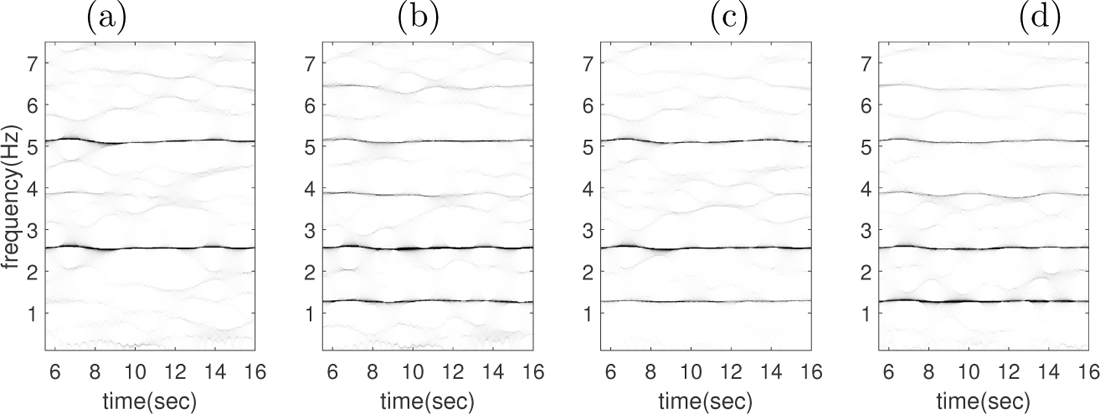
The TFR of the 60th to the 90th second IMU segment of the 6th subject in the TROIKA datasets, who is running at the speed of 6 kilometer per hour during this segment. We show the second-order SST with the Gaussian window with *σ* = 1.5 second. From (a) to (d): the TFR of the magnitude of the IMU signal, and the acceleration along the x-axis, y-axis and z-axis.

In another illustration (Figure 4) also from the TROIKA database [35], we present a PPG segment labeled “no-artifact” (or high quality, shown as the gray curve in Figure 4(a)). This segment is the 65.5th second to the 76th second segment from the recordings of the 12th subject in the TROIKA database (DATA 12 TYPE02.mat), and the subject is also running at the speed of 6 kilometer per hour in this segment. The label seems reasonable since the PPG segment mimics cardiac oscillations. However, these cycles do not align with true cardiac rhythm, which is confirmed by reading the simultaneously recorded ECG signal (red curve in Figure 4(d)), where again the R peak locations are superimposed on the second row as gray vertical lines. This segment includes multiple oscillatory components, including the cardiac component (Figure 4(d), with ECG in red) and the motion rhythm (Figure 4(c), with the x-axis accelerometer signal in red), which is guided by the TFR shown in Figure 5(a). Notably, the decomposed cardiac component reveals an oscillation pattern distinct from the raw signal, aligning well with the confirmed cardiac rhythm from the simultaneously recorded ECG. If our scientific interest lies in the motion rhythm or the cardiac component, the quality of this PPG should be considered high. While the experts’ label appears to align with this observation, it stems from a different perspective. It is worth mentioning that unlike the signal shown in Figure 1, the motion rhythm in this segment has a clear fundamental component, which is represented as a line at 1.3 Hz shown in both TFRs of Figure 5 indicated by a blue arrow in the green dashed box (also see Figure 6 for the TFRs of the simultaneously recorded IMU signal). This fundamental component results in the up-down pattern of the troughs in the reconstructed motion rhythm depicted in Figure 4(c), particularly by the blue dashed box.

**Figure 4.**
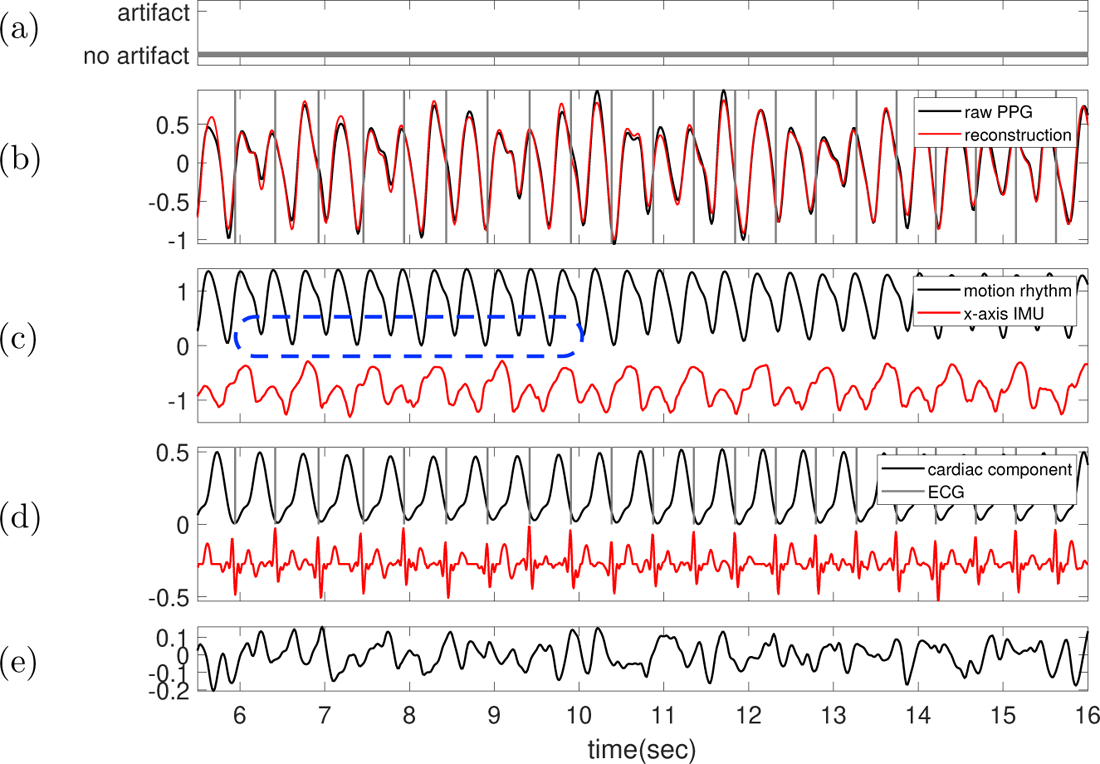
The 65.5th to 76th second segment of the 12th subject in the TROIKA datasets, who is running at the speed of 6 kilometer per hour during this segment. **(a)** Label sequence (grey line) and the prediction result by the proposed SQA model (red-dashed line). **(b)** The raw PPG signal is shown in black, and the summation of the decomposed motion rhythm and cardiac component is superimposed in red. **(c)** The decomposed motion component (black curve) and the synchronously recorded x-axis accelerometer signal (red curve). **(d)** The decomposed cardiac component (black curve), the synchronously recorded ECG (red curve) and the detected R-peaks (grey lines). **(e)** The difference of the raw PPG signal and the summation of the decomposed motion rhythm and cardiac component. The decomposition NRMSE is 0.116.

**Figure 5.**
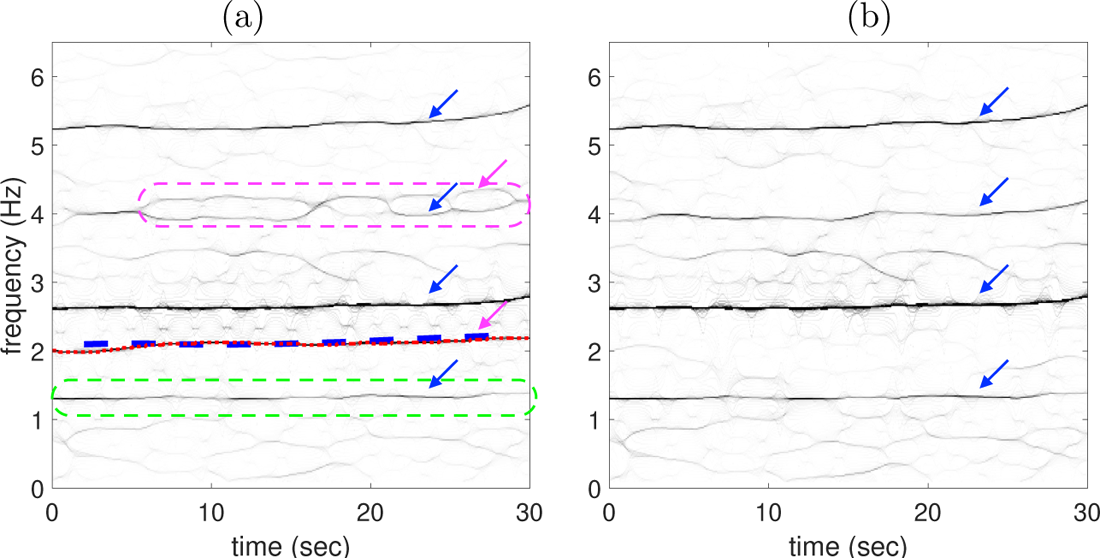
The TFR of the 60th to the 90th second PPG segment of the 12th subject in the TROIKA datasets, who is running at the speed of 6 kilometer per hour during this segment. We show the second-order SST with the Gaussian window with *σ* = 1.5 second. **(a)** The TFR of the original PPG signal, where the blue-dashed line is the ECG-derived IHR, and the red-dotted line is the result of the RD algorithm. **(b)** The TFR of the PPG signal after removing the cardiac component.

**Figure 6.**
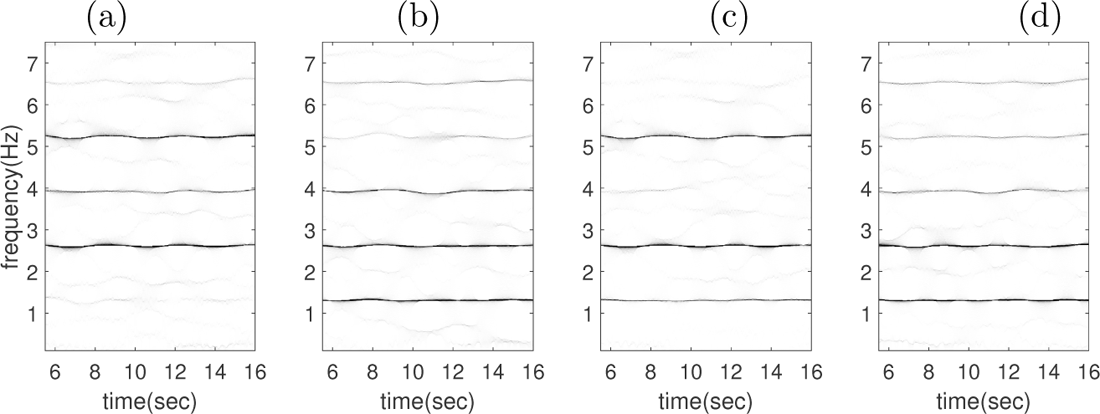
The TFR of the 60th to the 90th second IMU segment of the 12th subject in the TROIKA datasets, who is running at the speed of 6 kilometer per hour during this segment. We show the second-order SST with the Gaussian window with *σ* = 1.5 second. From (a) to (d): the TFR of the magnitude of the IMU signal, and the acceleration along the x-axis, y-axis and z-axis.

Based on the two examples above (and more shown in the appendix), our conclusion is that relying on the raw PPG signal for signal quality assessment may introduce bias, even when the label is determined by a human expert with other channels available. We thus advocate a reconsideration of what constitutes “high quality” when analyzing PPG signals. Consider Figures 1 and 4 again. Both PPG signals not only exhibit a clear cardiac component (in the subplot (d)) but also a distinct motion rhythm, displaying different patterns. Therefore, if the scientific focus is on cardiac dynamics like pulse rate, the signal should be deemed of high quality, though this fact is not evident from the raw PPG signal, even if other channels are considered to help the annotation.

Taking the pulse rate as an example, we selected “high-quality” segments from the TROIKA database, where “high quality” is defined as having more than 70% of subsegments labeled as non-artifact by experts. After excluding one subject exhibiting a strong coupling between motion rhythm and cardiac component, we identified 34 high-quality segments for analysis. We treated the ECG-derived mean heart rate (HR) and instantaneous heart rate (IHR) as the ground truth for a comparison. The summary statistics HR evaluated from ECG (ECG-HR), the mean pulse rate, as a surrogate for mean heart rates, evaluated from PPG (PPG-HR), and the extracted cardiac component (Cardiac-HR), are presented in Table 1. We utilize the Wilcoxon signed-rank test with the null hypothesis stating that two HRs share the same median, and we consider *p <* 0.05 to indicate statistical significance. The null hypothesis is rejected (not rejected respectively) when comparing p-value of 0.174, respectively). Next, we compare the IHR derived from ECG (ECG-IHR), PPG (PPG-IHR) and the extracted cardiac component (Cardiac-IHR). The IHR is evaluated by interpolating the inverse of the peak-to-peak intervals by cubic interpolation and sampled at 4 Hz. The normalized root mean square error (NRMSE) between PPG-IHR (or Cardiac-IHR) and ECG-IHR is assessed, and the null hypothesis, positing that these two NRMSEs share the same median, is rejected with a p-value of 2.48 *×* 10*^−^*^7^ through the utilization of the Wilcoxon signed-rank test. See the appendix for more details. It is worth noting that in practical research scenarios, researchers might opt to use segments recognized as high quality, especially those identified by experts, and proceed directly with analysis, such as evaluating heart rate for scientific exploration. However, this result suggests that such an approach may yield problematic results.

**Table 1.**
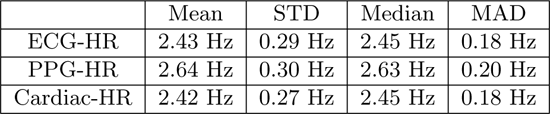
The summary statistics of the derived mean heart rates (HR).

In general, we may have interest in more complicated information from the cardiac component like pulse rate variability (PRV) [15] from the instantaneous pulse rate, and reflective index (RI) [17], stiffness index (SI) [8] and dicrotic notch analysis [22] from the cardiac component morphological analysis, etc, but the analysis shown above indicates that without a proper quantification of signal quality, even pulse rate information could be wrong. Similarly, if the goal is to study the motion rhythm, this PPG should *also* be considered “high quality” since it contains a component oscillating at the same frequency as the simultaneously recorded accelerometer signal. Once again, however, this fact is not apparent from the raw PPG signal alone.

We finally could provide our answer to the question “what PPG signal quality really means” – the signal quality depends on the quantities we want to extract from the PPG signal for the pre-defined scientific interest, and it should be determined after removing deterministic information that is irrelevant to the scientific interest. For example, if we want to study cardiac component related information, we take the ANHM (1) to model the clean PPG signal so that the recorded PPG signal is a realization of *Y* (*t*) = *f*_0_(*t*) + *ξ*(*t*) = *a_f_*(*t*)*s_f_*(*φ_f_*(*t*))+*T* (*t*)+*ξ*(*t*), where *ξ*(*t*) is a random process modeling the noise and *L >* 1 with *a_k_*(*t*)*s_k_*(*φ_k_*(*t*)) modeling the cardiac component. Under this model, we advocate that the signal quality should be determined using the realization of *Y*_1_(*t*) = *a_k_*(*t*)*s_k_*(*φ_k_*(*t*))+*ξ*(*t*), which is the signal after removing irrelevant deterministic information. For example, we can determine the signal quality of the cardiac component by using the energy ratio between the signal and noise [1] or existing signal quality assessment algorithms [6, 24, 31, 28]. Since we do not have a proper database with expert-determined labels under this consideration, we postpone the exploration of designing a proper signal quality index to our future work. Moreover, the signal quality quantification should be chosen based on which index we are after. In particular, the signal quality quantification for pulse rate should be different from RI, since more morphological features are needed for RI. This decomposition idea has been implicitly utilized in the literature. For example, when the scientific interest is evaluating the oxygen level from the PPG sensor installed on the forehead, it has been known that the existence of venous pulsation is a possible source of error [30], which suggests that the venous pulsation waveform should be removed before obtaining a reliable oxygen level. As a result, an alternative and systematic approach based on signal decomposition for determining signal quality adapted to scientific purposes, including how the label should be provided, is warranted, and it will be our future direction.

## 4. Conclusion

The main contribution is showing the necessity of reconsidering the meaning of high quality PPG signals by providing evidence based on the ANHM for the PPG signal and a signal decomposition algorithm. The same consideration can be extended to other biomedical signals that we do not explore in this paper. We shall emphasize that the proposed ANHM model and the decomposition algorithm SAMD do not conflict with existing signal quality evaluation procedures. For example, we could apply existing signal quality assessment algorithms [6, 24, 31, 28] to the decomposed cardiac component if our scientific interest is related to the cardiac component. Moreover, after obtaining the high quality signal of interest, we could proceed to the ordinary signal processing procedure. For example, if estimating the blood pressure [11], pulse decomposition analysis (PDA) [3], or detecting arrhythmia [25] is of interest, we could apply existing algorithms on the decomposed cardiac component. The relationship SAMD and PDA deserves clarification. Though distinct, they are related as follows: According to the ANHM, PDA is an algorithm designed to decompose a *single* cardiac cycle into various template functions, such as Gaussian or Gamma functions, to enhance the quantification of cardiac features. On the other hand, SAMD is a decomposition algorithm used to break down the raw PPG signal into different oscillatory components; see Figures 1 and 4 for an example of extracting the cardiac component.

In conclusion, the exploration presented in this paper underscores the necessity for designing a new signal quality evaluation system, especially when dealing with PPG signals recorded in the free-living environment. Our proposed ANHM model, in conjunction with advanced signal decomposition tools, holds promise for establishing such a system by incorporating the signal decomposition step. The development of this system will be reported in our future work. With labels provided by this system, we believe we can advance towards establishing a more dependable SQA model, particularly for scientific research.

## Data Availability

All data produced in the present work are available at https://github.com/chengstark/Segade/tree/main/data

## Acknowledgement

The authors express gratitude to the authors in [9] for their assistance in providing details regarding the labeled databases they shared.

## Appendix A. More numerical results

We utilized segments from the TROIKA database [35] that have been labeled as high quality by experts, following the criteria outlined in the main article or [9]. Here, “high quality by experts” is defined as having more than 70% of subsegments labeled as “non-artifact” by experts. There are a total of 12 subjects and 113 annotated 30-second-long segments in the TROIKA database. The distribution of the ratio of non-artifact subsegments in a 30-second segment is illustrated in Figure 7. Among these segments, 42 segments contain less than 30% of subsegments labeled as “non-artifact”.

**Figure 7.**
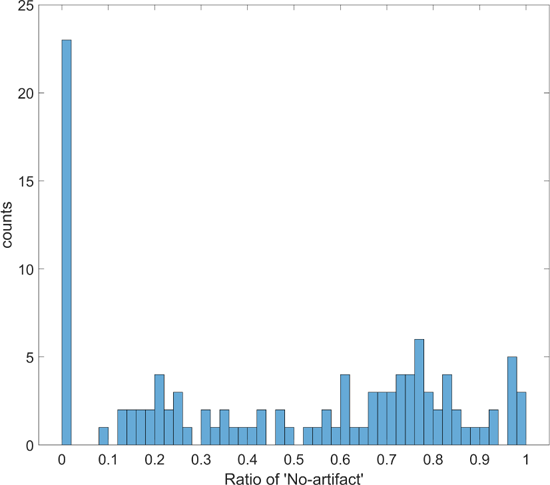
The histogram of the ratio of non-artifact subsegments in a 30-second segment.

One subject exhibits a strong coupling between motion rhythm and the cardiac component, wherein the instantaneous heart rate is twice the instantaneous frequency of the motion rhythm. Since decomposing the cardiac component and motion rhythm from this signal type surpasses the capability of both the model and the SAMD algorithm, this subject is excluded from the analysis. However, this intriguing case will be investigated in future work. Consequently, there remain a total of 34 high-quality segments for analysis. Below, we treated the ECG-derived mean heart rate (HR) and instantaneous heart rate (IHR) as the ground truth for a comparison.

The boxplots of the HR evaluated from ECG (ECG-HR), the mean pulse rate, as a surrogate for mean heart rates, evaluated from PPG (PPG-HR), and the extracted cardiac component (Cardiac-HR), are depicted in Figure 8. Here, we employ a standard peak detection algorithm to identify R peaks in ECG and peaks in both PPG and the extracted cardiac component. Subsequently, we calculate the mean of the inverse of the peak-to-peak interval to determine HR. We utilize the Wilcoxon signed-rank test with the null hypothesis stating that two HRs share the same median, and we consider *p <* 0.05 to indicate statistical significance. The null hypothesis is rejected (not rejected respectively) when comparing ECG-HR and PPG-HR (Cardiac-HR respectively) with a p-value of 1.15*×*10*^−^*^6^ (1.74*×*10*^−^*^1^, respectively). Next, we compare the IHR derived from ECG (ECG-IHR), PPG (PPG-IHR) and the extracted cardiac component (Cardiac-IHR). The IHR is evaluated by interpolating the inverse of the peak-to-peak intervals by cubic interpolation and sampled at 4 Hz. The normalized root mean square error (NRMSE) between PPG-IHR (or Cardiac-IHR) and ECG-IHR is assessed, and their boxplots are depicted in Figure 9. The null hypothesis, positing that these two NRMSEs share the same median, is rejected with a p-value of 2.48 *×* 10*^−^*^7^ through the utilization of the Wilcoxon signed-rank test.

**Figure 8.**
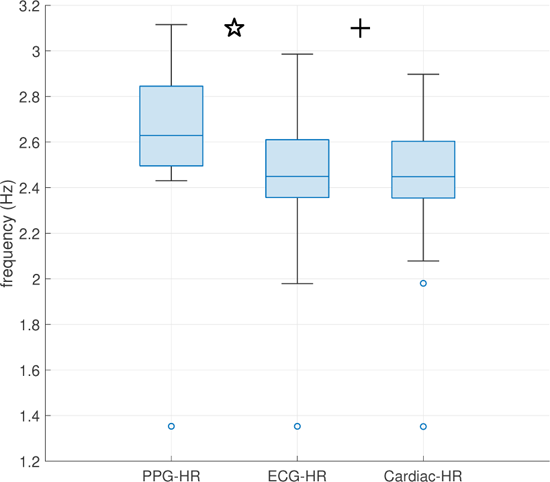
The boxplot of the mean heart rate (HR) derived from different signals, including ECG, denoted as ECG-HR, PPG, denoted as PPG-HR, and the decomposed cardiac component, denoted as cardiac-HR. The star sign indicates that the null hypothesis is rejected with a p-value of 1.15 *×* 10*^−^*^6^. The plus sign indicates that the null hypothesis cannot be rejected with a p-value of 1.74 *×* 10*^−^*^1^.

**Figure 9.**
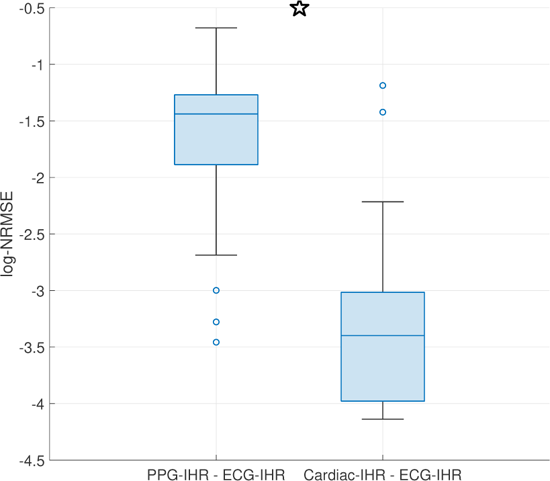
The boxplot illustrates the log-normalized root mean square error (log-NRMSE) when comparing various instantaneous heart rates (IHR), including those derived from ECG (ECG-IHR), PPG (PPG-IHR), and the extracted cardiac component (Cardiac-IHR). On the left-hand side are the log-NRMSE values comparing ECG-IHR and PPG-IHR, while on the right-hand side are the log-NRMSE values comparing ECG-IHR and Cardiac-IHR. The star sign indicates that the null hypothesis is rejected with a p-value of 2.48 *×* 10*^−^*^7^.

Figures 10–77 showcase results of these 34 segments, including the time-frequency representations and decomposed signals, along with the simultaneously recorded electrocardiogram and accelerometer signal.

**Figure 10.**
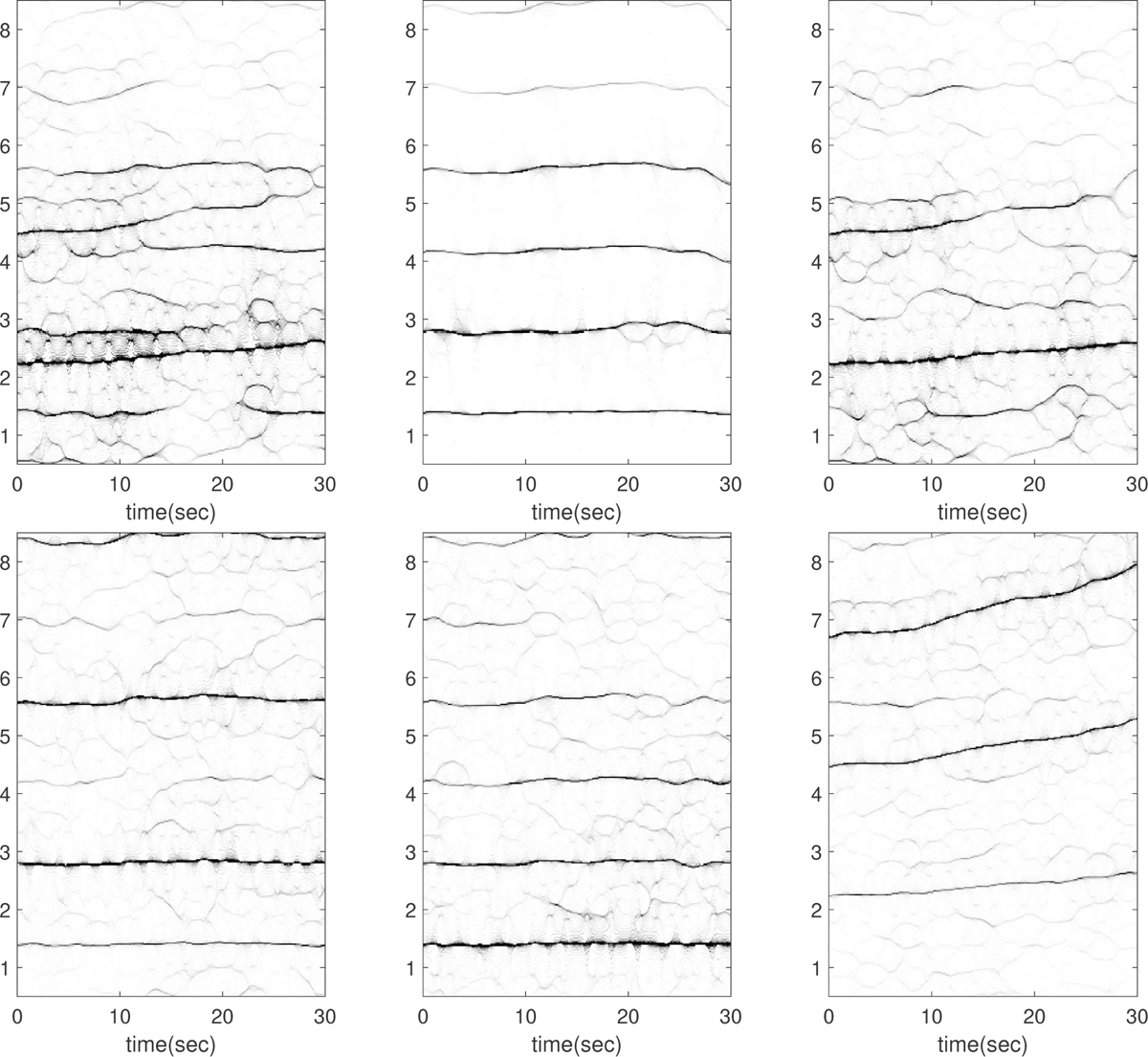
Subject 4 between 210 and 240 seconds in the TROIKA dataset. The subject is running at the speed of 12 kilometer per hour during this segment. The first row, from left to right: the time-frequency representations (TFRs) of the PPG, the decomposed motion rhythm component, and the PPG after removing the decomposed motion rhythm component. The second row, from left to right: the TFRs of the simultaneously recorded accelerometer magnitude signal, the simultaneously recorded x-axis of the accelerometer signal, and the simultaneously recorded ECG signal. Over this segment, the mean heart rate derived from the simultaneously recorded electrocardiogram (resp. raw PPG and extracted cardiac component) is 2.41 Hz (resp. 2.53 Hz, 2.40 Hz). In this case, the instantaneous heart rate increases from approximately 2.2 Hz in the beginning to approximately 3.6 Hz in the end. In the TFR of the ECG, there is a plausible component oscillating at approximately 5.6 Hz, aligning with the fourth harmonic of the motion rhythm.

**Figure 11.**
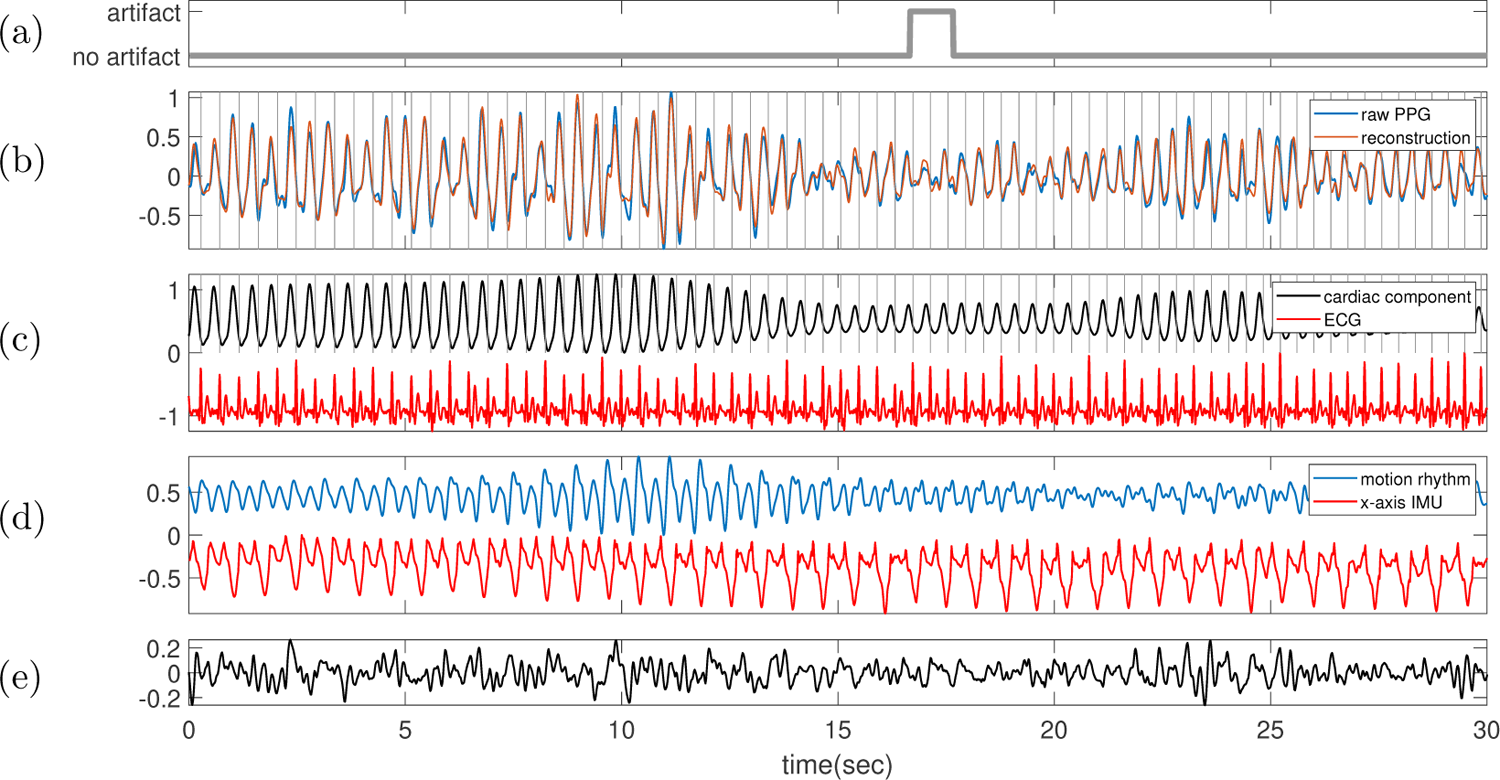
Subject 4 between 210 and 240 seconds in the TROIKA dataset. The subject is running at the speed of 12 kilometer per hour during this segment. **(a)** Label sequence (grey line) and the prediction result by the proposed SQA model (red-dashed line). **(b)** The raw PPG signal is shown in black, and the summation of the decomposed motion rhythm and cardiac component is super-imposed in red. The detected R-peaks from the simultaneously recorded ECG are superimposed as vertical grey lines. **(c)** The decomposed cardiac component (black curve), the simultaneously recorded ECG (red curve) and the detected R-peaks (vertical grey lines). **(d)** The decomposed motion rhythm (blue curve) and the x-axis of the accelerometer signal recorded simultaneously (red curve). **(e)** The difference of the PPG signal and the summation of the decomposed motion rhythm and cardiac component. The normalized root mean square error (NRMSE) between the PPG signal and the summation of the decomposed motion rhythm and cardiac component reconstruction is 0.25.

**Figure 12.**
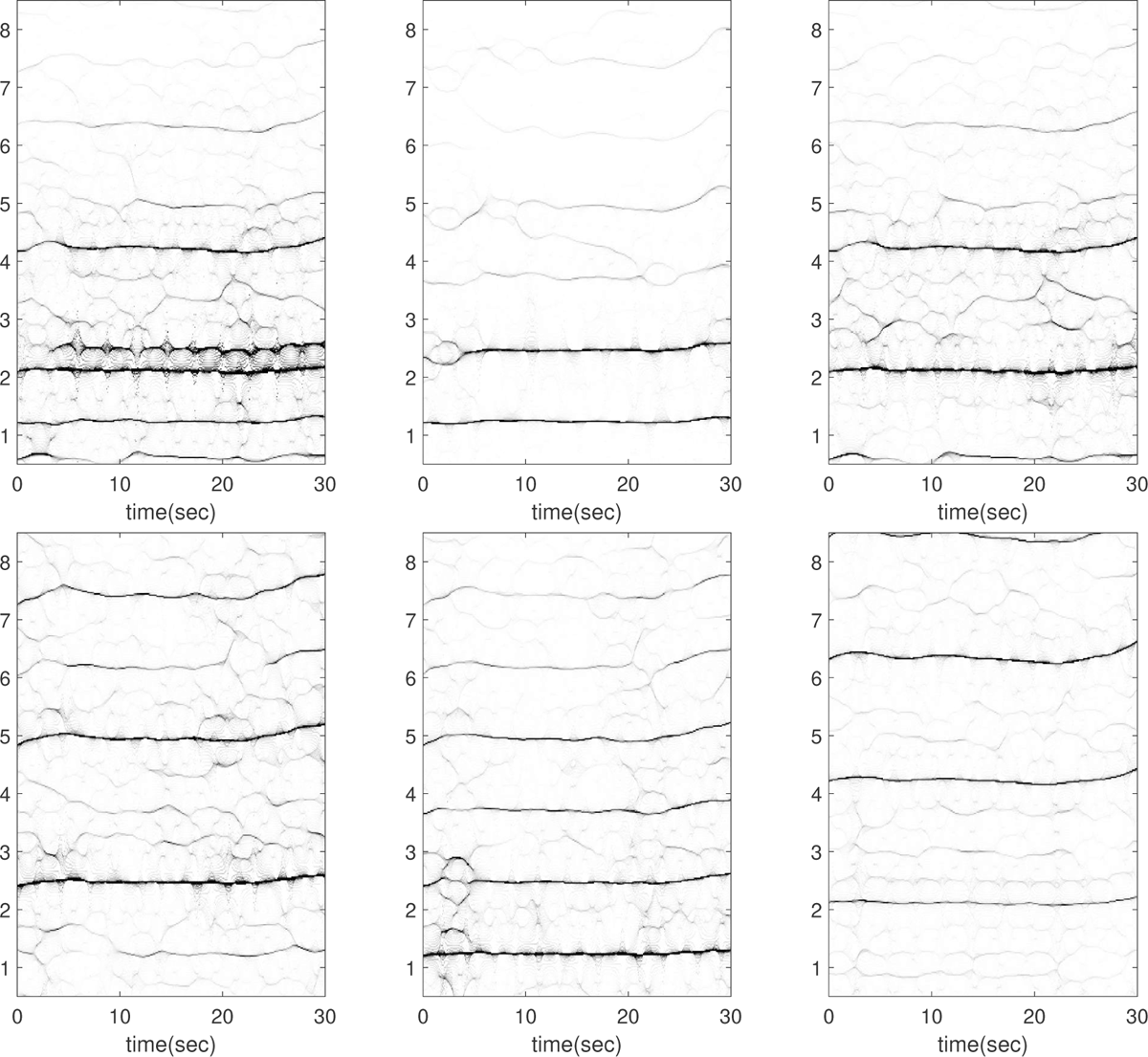
Subject 5 between 60 and 90 seconds in the TROIKA dataset. The subject is running at the speed of 6 kilometer per hour during this segment. The first row, from left to right: the time-frequency representations (TFRs) of the PPG, the decomposed motion rhythm component, and the PPG after removing the decomposed motion rhythm component. The second row, from left to right: the TFRs of the simultaneously recorded accelerometer magnitude signal, the simultaneously recorded x-axis of the accelerometer signal, and the simultaneously recorded ECG signal. Over this segment, the mean heart rate derived from the simultaneously recorded electrocardiogram (resp. raw PPG and extracted cardiac component) is 2.13 Hz (resp. 2.49 Hz, 2.12 Hz).

**Figure 13.**
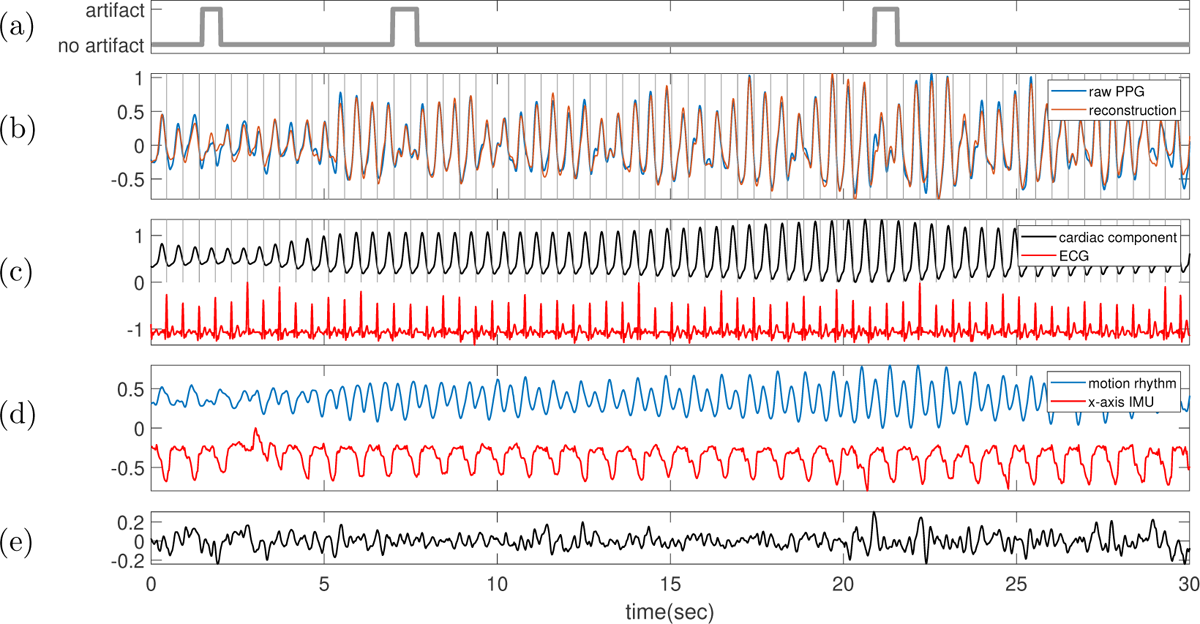
Subject 5 between 60 and 90 seconds in the TROIKA dataset. The subject is running at the speed of 6 kilometer per hour during this segment. **(a)** Label sequence (grey line) and the prediction result by the proposed SQA model (red-dashed line). **(b)** The raw PPG signal is shown in black, and the summation of the decomposed motion rhythm and cardiac component is superimposed in red. The detected R-peaks from the simultaneously recorded ECG are superimposed as vertical grey lines. **(c)** The decomposed cardiac component (black curve), the simultaneously recorded ECG (red curve) and the detected R-peaks (vertical grey lines). **(d)** The decomposed motion rhythm (blue curve) and the x-axis of the accelerometer signal recorded simultaneously (red curve). **(e)** The difference of the PPG signal and the summation of the decomposed motion rhythm and cardiac component. The normalized root mean square error (NRMSE) between the PPG signal and the summation of the decomposed motion rhythm and cardiac component reconstruction is 0.20.

**Figure 14.**
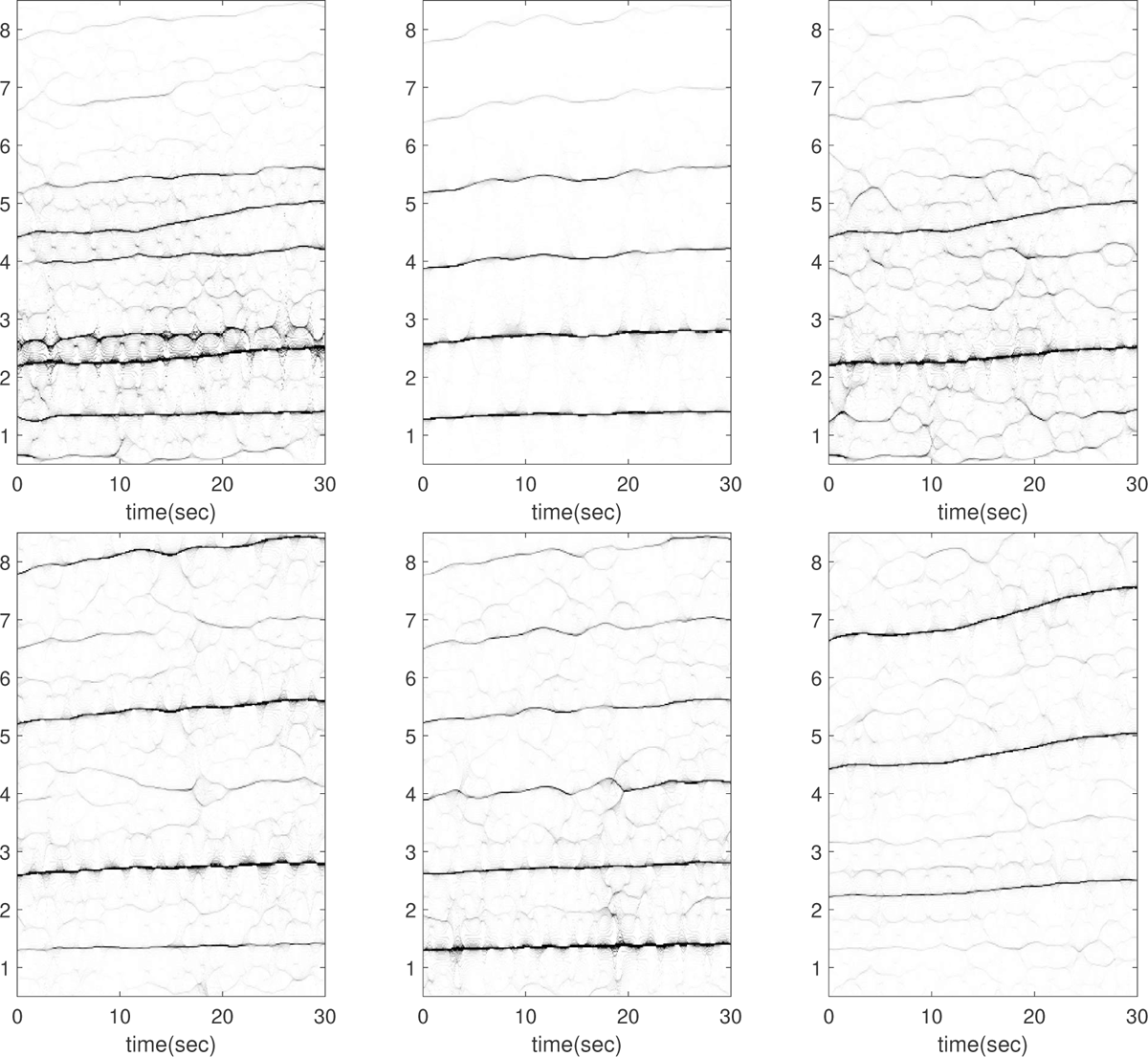
Subject 5 between 90 and 120 seconds in the TROIKA dataset. The subject is running at the speed of 12 kilometer per hour during this segment. The first row, from left to right: the time-frequency representations (TFRs) of the PPG, the decomposed motion rhythm component, and the PPG after removing the decomposed motion rhythm component. The second row, from left to right: the TFRs of the simultaneously recorded accelerometer magnitude signal, the simultaneously recorded x-axis of the accelerometer signal, and the simultaneously recorded ECG signal. Over this segment, the mean heart rate derived from the simultaneously recorded electrocardiogram (resp. raw PPG and extracted cardiac component) is 2.35 Hz (resp. 2.63 Hz, 2.35 Hz). In this case, the instantaneous frequency of the motion rhythm and the instantaneous heart rate both increase as time goes. From the TFR of the ECG, there are plausible components oscillating at approximately 1.3 Hz, 2.6 Hz, and 3.3 Hz in the ECG. Among these, the components oscillating at approximately 1.3 Hz and 2.6 Hz align with the first and second harmonics of the motion rhythm, respectively. However, the source of the oscillation at approximately 3.3 Hz remains unknown.

**Figure 15.**
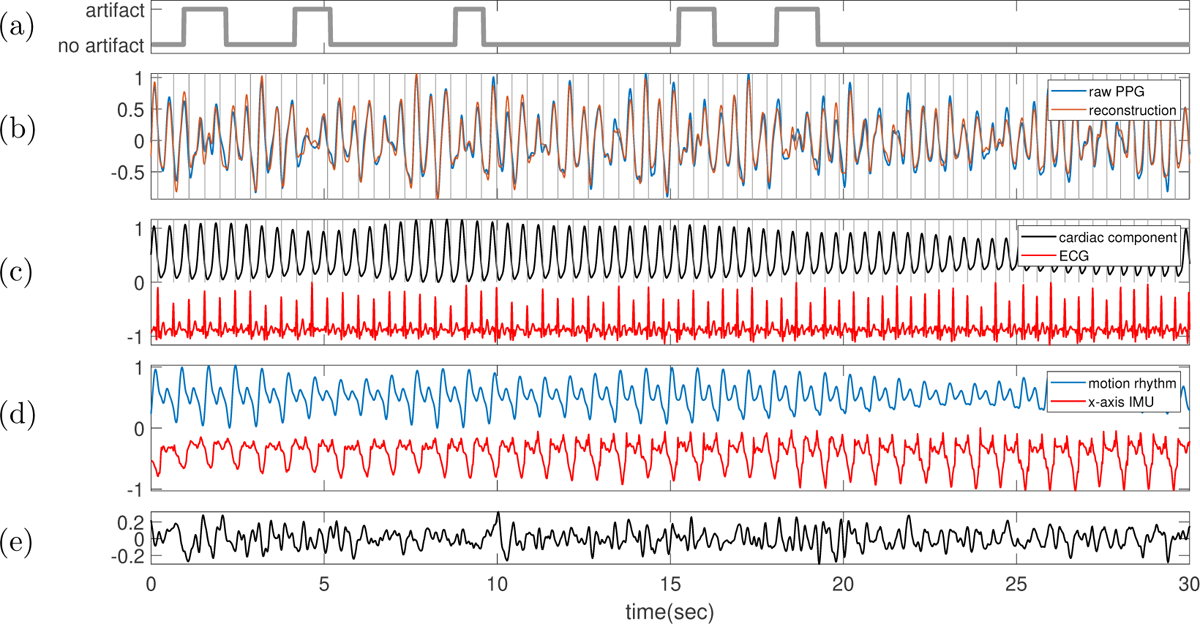
Subject 5 between 90 and 120 seconds in the TROIKA dataset. The subject is running at the speed of 12 kilometer per hour during this segment. **(a)** Label sequence (grey line) and the prediction result by the proposed SQA model (red-dashed line). **(b)** The raw PPG signal is shown in black, and the summation of the decomposed motion rhythm and cardiac component is superimposed in red. The detected R-peaks from the simultaneously recorded ECG are superimposed as vertical grey lines. **(c)** The decomposed cardiac component (black curve), the simultaneously recorded ECG (red curve) and the detected R-peaks (vertical grey lines). **(d)** The decomposed motion rhythm (blue curve) and the x-axis of the accelerometer signal recorded simultaneously (red curve). **(e)** The difference of the PPG signal and the summation of the decomposed motion rhythm and cardiac component. The normalized root mean square error (NRMSE) between the PPG signal and the summation of the decomposed motion rhythm and cardiac component reconstruction is 0.26.

**Figure 16.**
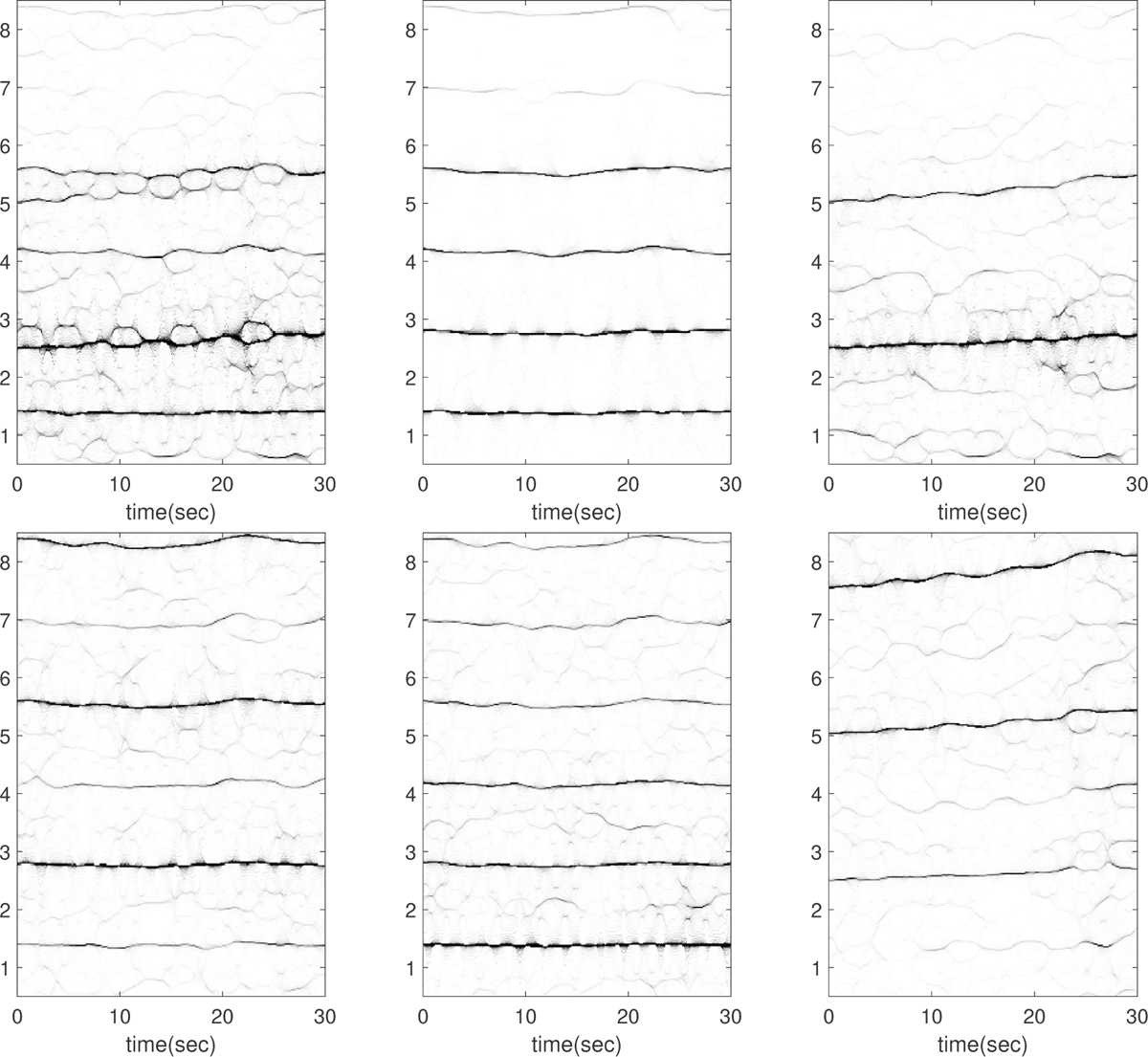
Subject 5 between 120 and 150 seconds in the TROIKA dataset. The subject is running at the speed of 12 kilometer per hour during this segment. The first row, from left to right: the time-frequency representations (TFRs) of the PPG, the decomposed motion rhythm component, and the PPG after removing the decomposed motion rhythm component. The second row, from left to right: the TFRs of the simultaneously recorded accelerometer magnitude signal, the simultaneously recorded x-axis of the accelerometer signal, and the simultaneously recorded ECG signal. Over this segment, the mean heart rate derived from the simultaneously recorded electrocardiogram (resp. raw PPG and extracted cardiac component) is 2.62 Hz (resp. 2.85 Hz, 2.62 Hz).

**Figure 17.**
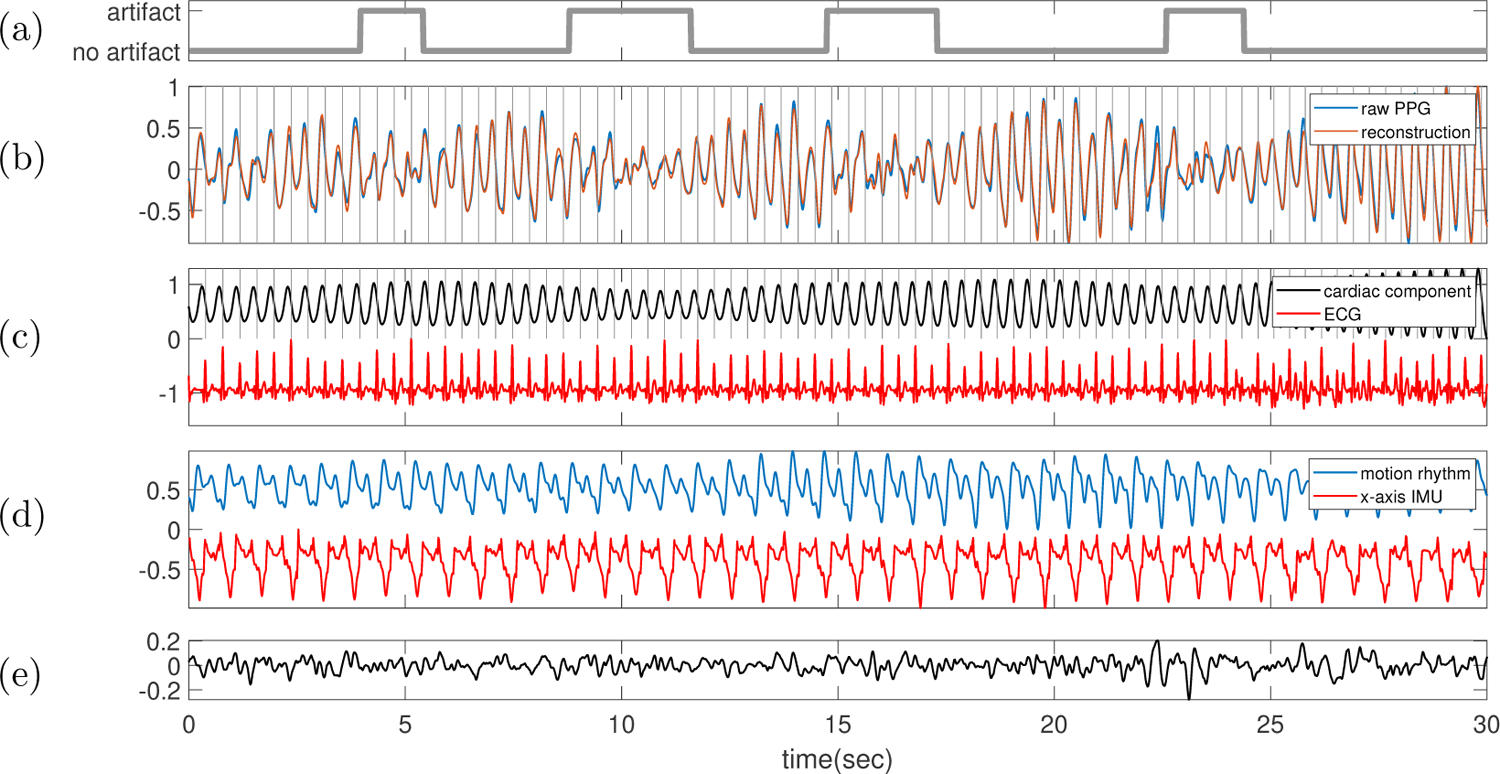
Subject 5 between 120 and 150 seconds in the TROIKA dataset. The subject is running at the speed of 12 kilometer per hour during this segment. **(a)** Label sequence (grey line) and the prediction result by the proposed SQA model (red-dashed line). **(b)** The raw PPG signal is shown in black, and the summation of the decomposed motion rhythm and cardiac component is super-imposed in red. The detected R-peaks from the simultaneously recorded ECG are superimposed as vertical grey lines. **(c)** The decomposed cardiac component (black curve), the simultaneously recorded ECG (red curve) and the detected R-peaks (vertical grey lines). **(d)** The decomposed motion rhythm (blue curve) and the x-axis of the accelerometer signal recorded simultaneously (red curve). **(e)** The difference of the PPG signal and the summation of the decomposed motion rhythm and cardiac component. The normalized root mean square error (NRMSE) between the PPG signal and the summation of the decomposed motion rhythm and cardiac component reconstruction is 0.16.

**Figure 18.**
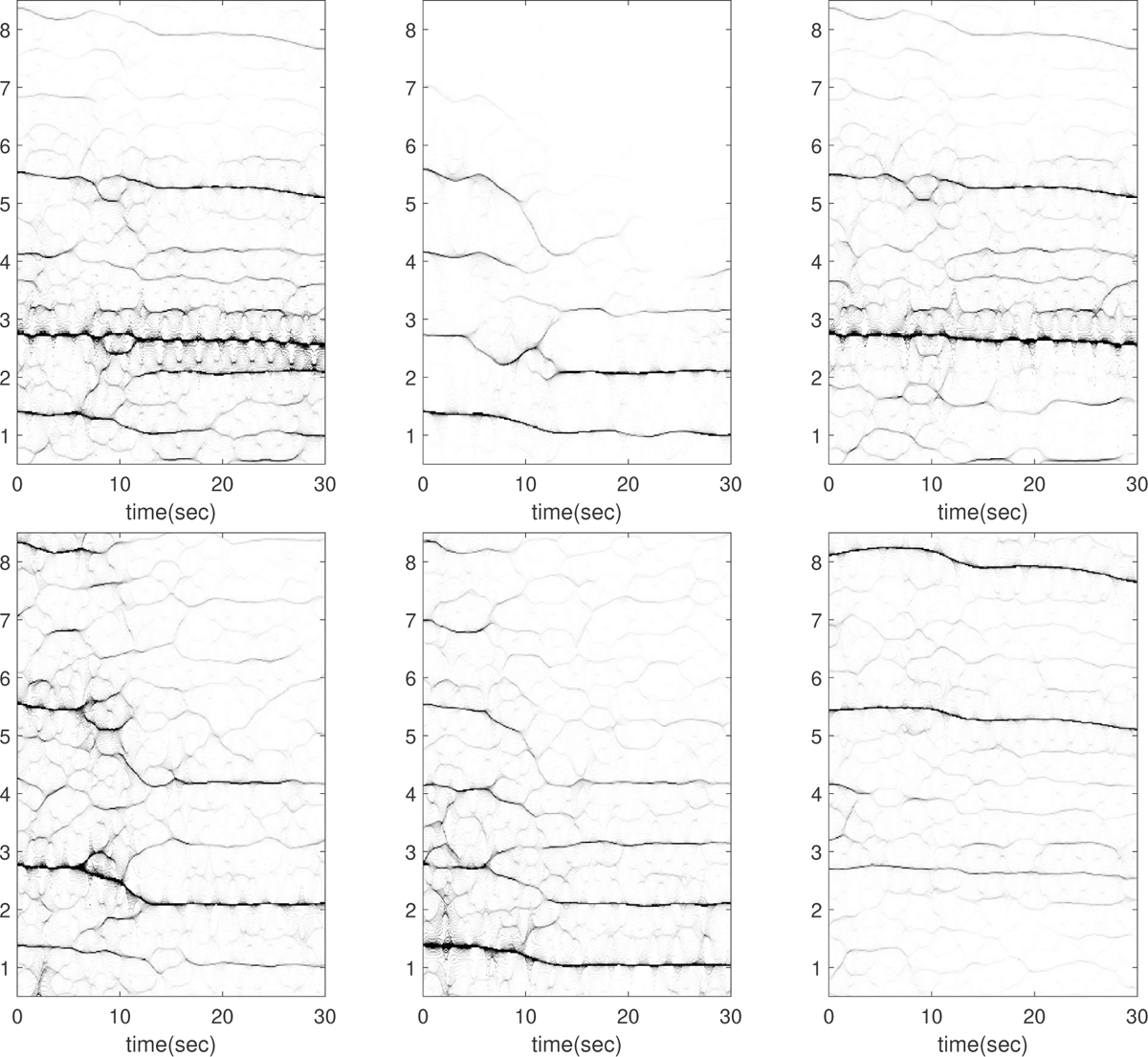
Subject 5 between 150 and 180 seconds in the TROIKA dataset. The subject is running at the speed of 6 kilometer per hour during this segment. The first row, from left to right: the time-frequency representations (TFRs) of the PPG, the decomposed motion rhythm component, and the PPG after removing the decomposed motion rhythm component. The second row, from left to right: the TFRs of the simultaneously recorded accelerometer magnitude signal, the simultaneously recorded x-axis of the accelerometer signal, and the simultaneously recorded ECG signal. Over this segment, the mean heart rate derived from the simultaneously recorded electrocardiogram (resp. raw PPG and extracted cardiac component) is 2.65 Hz (resp. 2.67 Hz, 2.67 Hz). The accelerometer is relatively noisy with a higher instantaneous frequency (*∼* 1.4 Hz) before the 10th second, compared with the signal after the 10th second,

**Figure 19.**
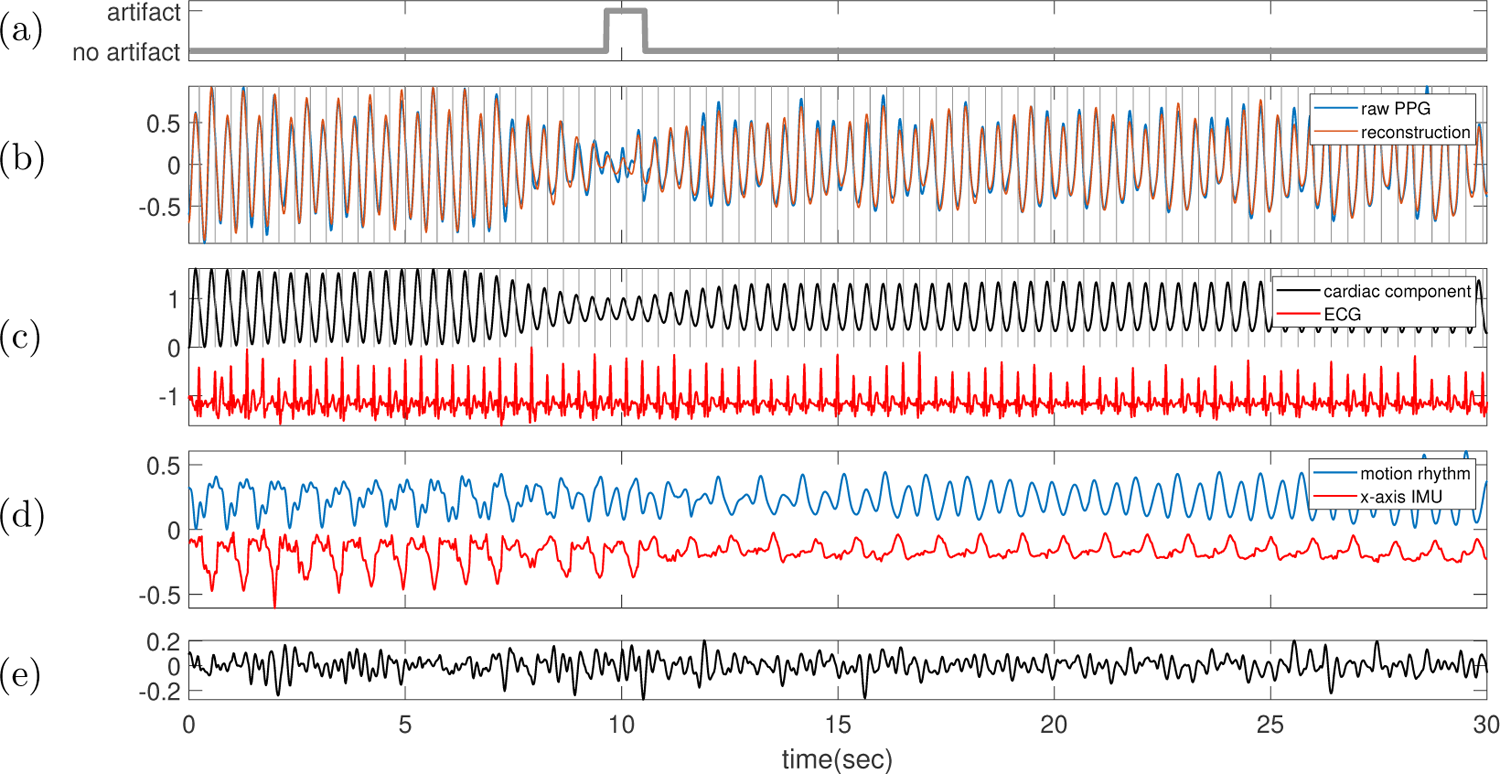
Subject 5 between 150 and 180 seconds in the TROIKA dataset. The subject is running at the speed of 6 kilometer per hour during this segment. **(a)** Label sequence (grey line) and the prediction result by the proposed SQA model (red-dashed line). **(b)** The raw PPG signal is shown in black, and the summation of the decomposed motion rhythm and cardiac component is super-imposed in red. The detected R-peaks from the simultaneously recorded ECG are superimposed as vertical grey lines. **(c)** The decomposed cardiac component (black curve), the simultaneously recorded ECG (red curve) and the detected R-peaks (vertical grey lines). **(d)** The decomposed motion rhythm (blue curve) and the x-axis of the accelerometer signal recorded simultaneously (red curve). **(e)** The difference of the PPG signal and the summation of the decomposed motion rhythm and cardiac component. The normalized root mean square error (NRMSE) between the PPG signal and the summation of the decomposed motion rhythm and cardiac component reconstruction is 0.17.

**Figure 20.**
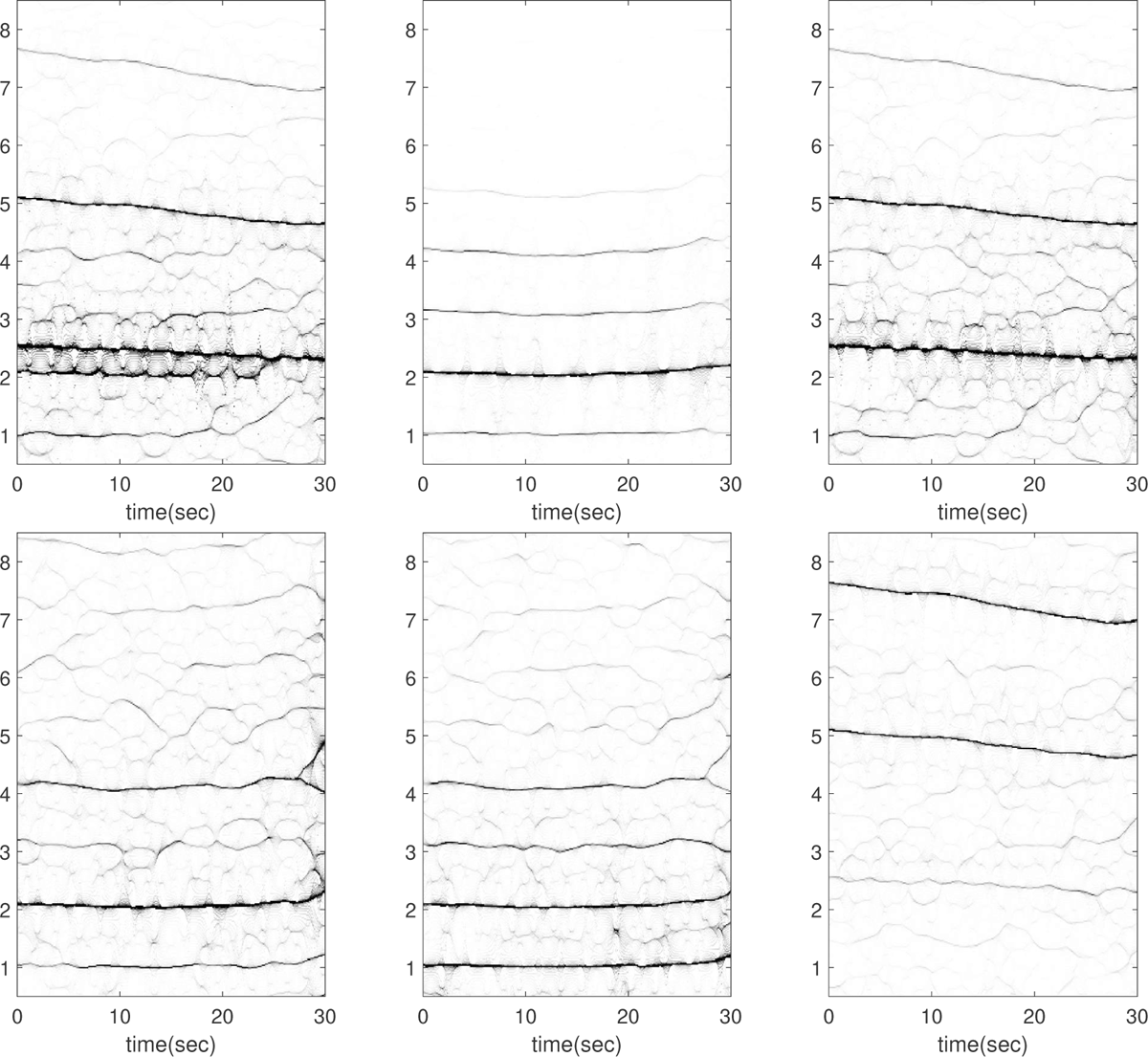
Subject 5 between 180 and 210 seconds in the TROIKA dataset. The subject is running at the speed of 6 kilometer per hour during this segment. The first row, from left to right: the time-frequency representations (TFRs) of the PPG, the decomposed motion rhythm component, and the PPG after removing the decomposed motion rhythm component. The second row, from left to right: the TFRs of the simultaneously recorded accelerometer magnitude signal, the simultaneously recorded x-axis of the accelerometer signal, and the simultaneously recorded ECG signal. Over this segment, the mean heart rate derived from the simultaneously recorded electrocardiogram (resp. raw PPG and extracted cardiac component) is 2.44 Hz (resp. 2.43 Hz, 2.43 Hz). Note that the fundamental component of the ECG signal (*∼* 2.5 Hz) is relatively weak in this segment.

**Figure 21.**
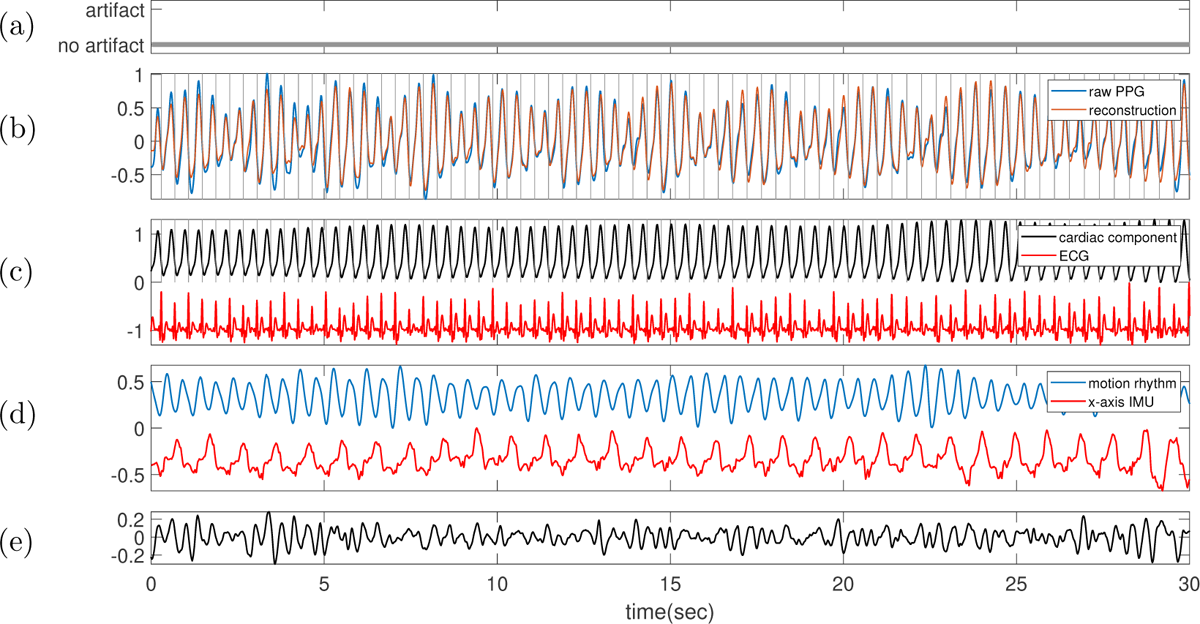
Subject 5 between 180 and 210 seconds in the TROIKA dataset. The subject is running at the speed of 6 kilometer per hour during this segment. **(a)** Label sequence (grey line) and the prediction result by the proposed SQA model (red-dashed line). **(b)** The raw PPG signal is shown in black, and the summation of the decomposed motion rhythm and cardiac component is super-imposed in red. The detected R-peaks from the simultaneously recorded ECG are superimposed as vertical grey lines. **(c)** The decomposed cardiac component (black curve), the simultaneously recorded ECG (red curve) and the detected R-peaks (vertical grey lines). **(d)** The decomposed motion rhythm (blue curve) and the x-axis of the accelerometer signal recorded simultaneously (red curve). **(e)** The difference of the PPG signal and the summation of the decomposed motion rhythm and cardiac component. The normalized root mean square error (NRMSE) between the PPG signal and the summation of the decomposed motion rhythm and cardiac component reconstruction is 0.22.

**Figure 22.**
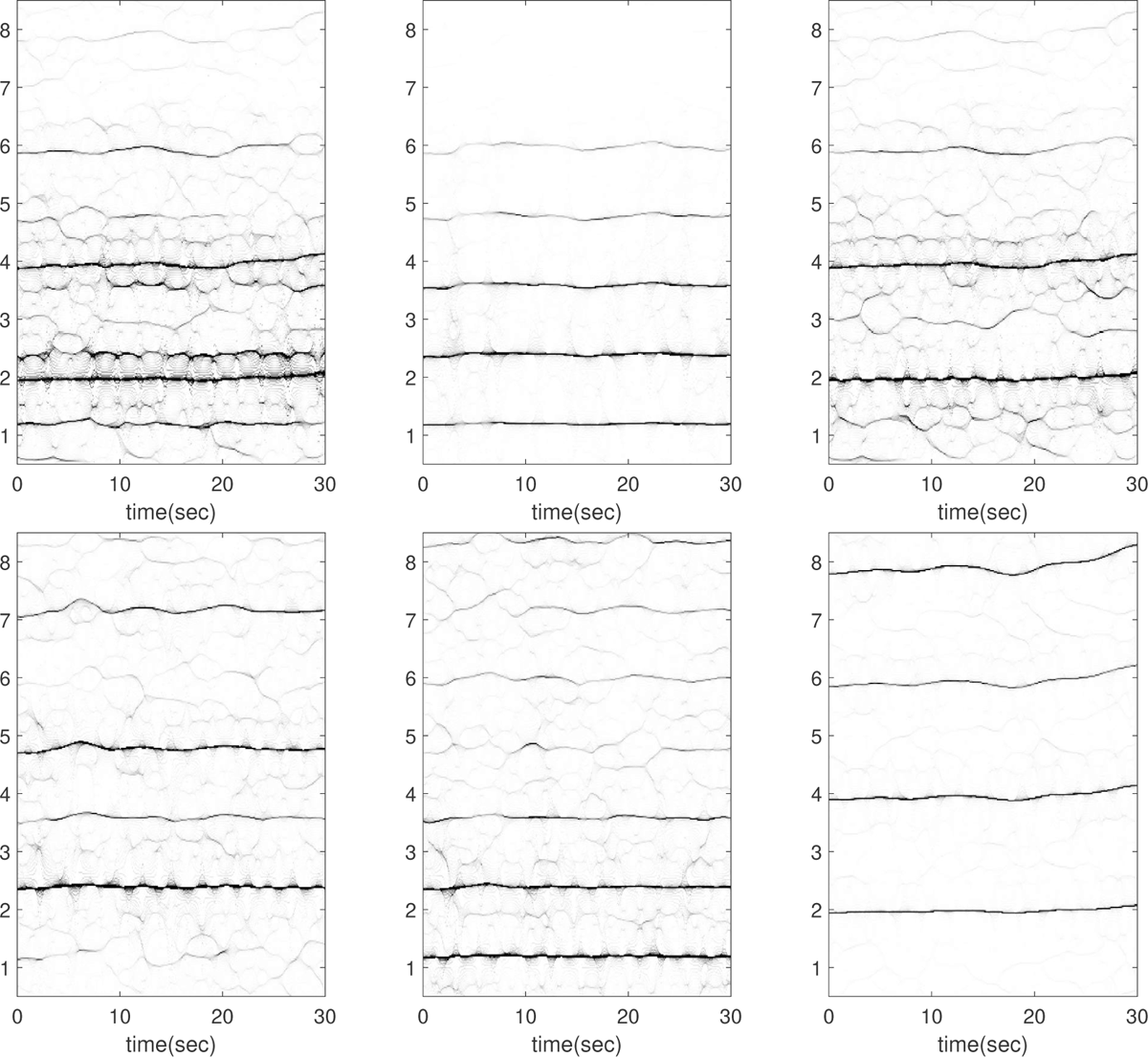
Subject 7 between 60 and 90 seconds in the TROIKA dataset. The subject is running at the speed of 6 kilometer per hour during this segment. The first row, from left to right: the time-frequency representations (TFRs) of the PPG, the decomposed motion rhythm component, and the PPG after removing the decomposed motion rhythm component. The second row, from left to right: the TFRs of the simultaneously recorded accelerometer magnitude signal, the simultaneously recorded x-axis of the accelerometer signal, and the simultaneously recorded ECG signal. Over this segment, the mean heart rate derived from the simultaneously recorded electrocardiogram (resp. raw PPG and extracted cardiac component) is 1.99 Hz (resp. 2.66 Hz, 1.98 Hz).

**Figure 23.**
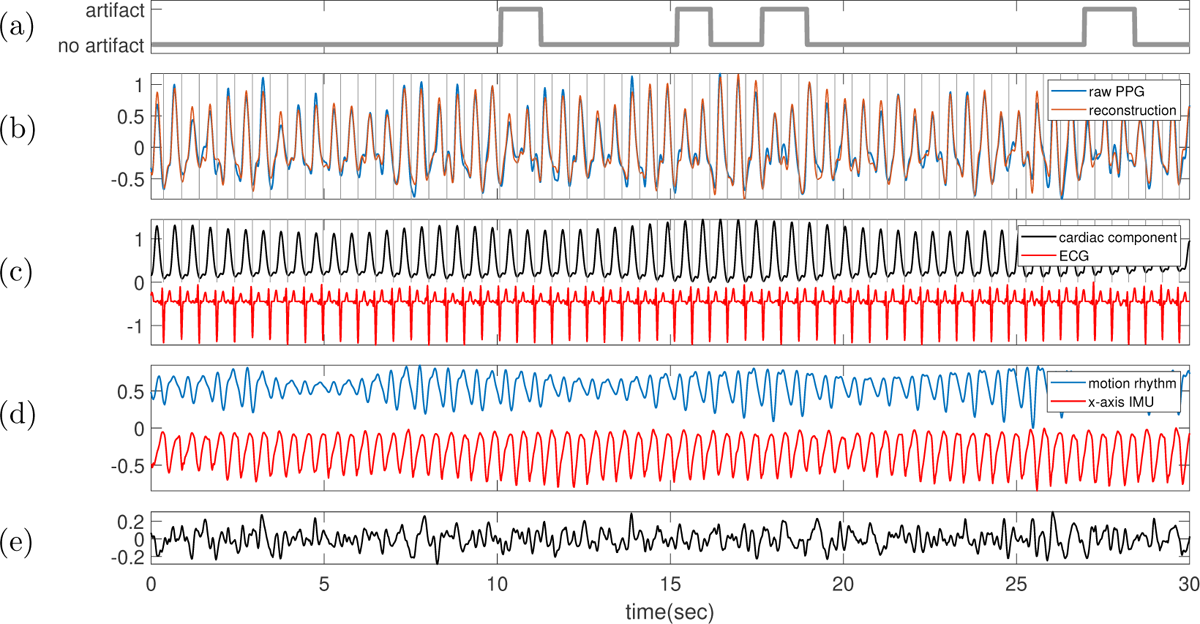
Subject 7 between 60 and 90 seconds in the TROIKA dataset. The subject is running at the speed of 6 kilometer per hour during this segment. **(a)** Label sequence (grey line) and the prediction result by the proposed SQA model (red-dashed line). **(b)** The raw PPG signal is shown in black, and the summation of the decomposed motion rhythm and cardiac component is superimposed in red. The detected R-peaks from the simultaneously recorded ECG are superimposed as vertical grey lines. **(c)** The decomposed cardiac component (black curve), the simultaneously recorded ECG (red curve) and the detected R-peaks (vertical grey lines). **(d)** The decomposed motion rhythm (blue curve) and the x-axis of the accelerometer signal recorded simultaneously (red curve). **(e)** The difference of the PPG signal and the summation of the decomposed motion rhythm and cardiac component. The normalized root mean square error (NRMSE) between the PPG signal and the summation of the decomposed motion rhythm and cardiac component reconstruction is 0.23.

**Figure 24.**
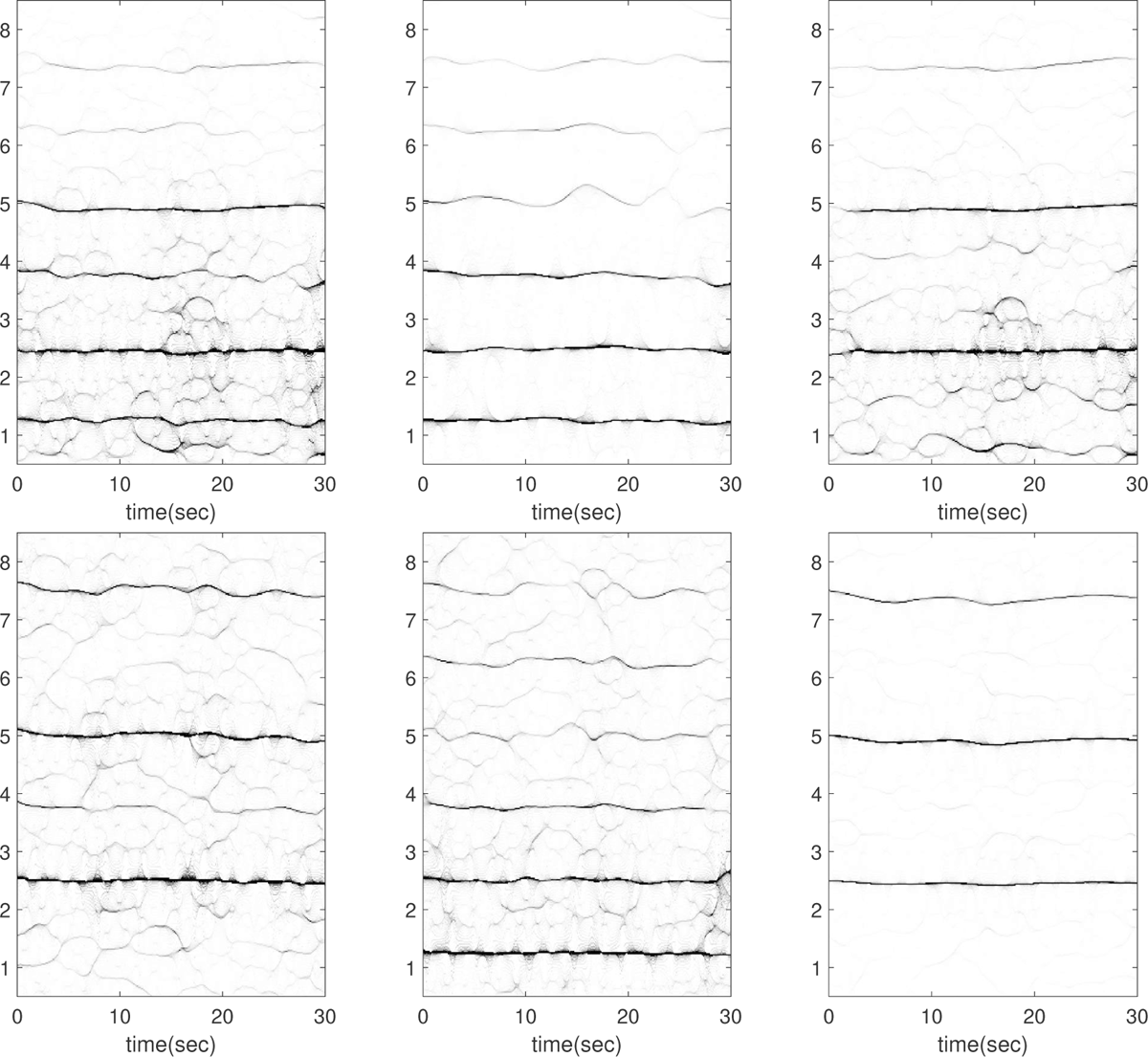
Subject 7 between 150 and 180 seconds in the TROIKA dataset. The subject is running at the speed of 6 kilometer per hour during this segment. The first row, from left to right: the time-frequency representations (TFRs) of the PPG, the decomposed motion rhythm component, and the PPG after removing the decomposed motion rhythm component. The second row, from left to right: the TFRs of the simultaneously recorded accelerometer magnitude signal, the simultaneously recorded x-axis of the accelerometer signal, and the simultaneously recorded ECG signal. Over this segment, the mean heart rate derived from the simultaneously recorded electrocardiogram (resp. raw PPG and extracted cardiac component) is 2.46 Hz (resp. 2.51 Hz, 2.46 Hz). In this case, the fundamental component of the accelerometer magnitude signal cannot be visualized. This segment is noteworthy because the instantaneous frequency (IF) of the second harmonic of the IMU signal matches exactly with that of the fundamental component of ECG in the first and last 10 seconds (see the top row of Figure 25); indicating a strong 2:1 coupling between heart rate and motion rhythm. The current algorithm is limited in decomposing such signals, and the decomposition is not ideal as illustrated in Figure 25.

**Figure 25.**
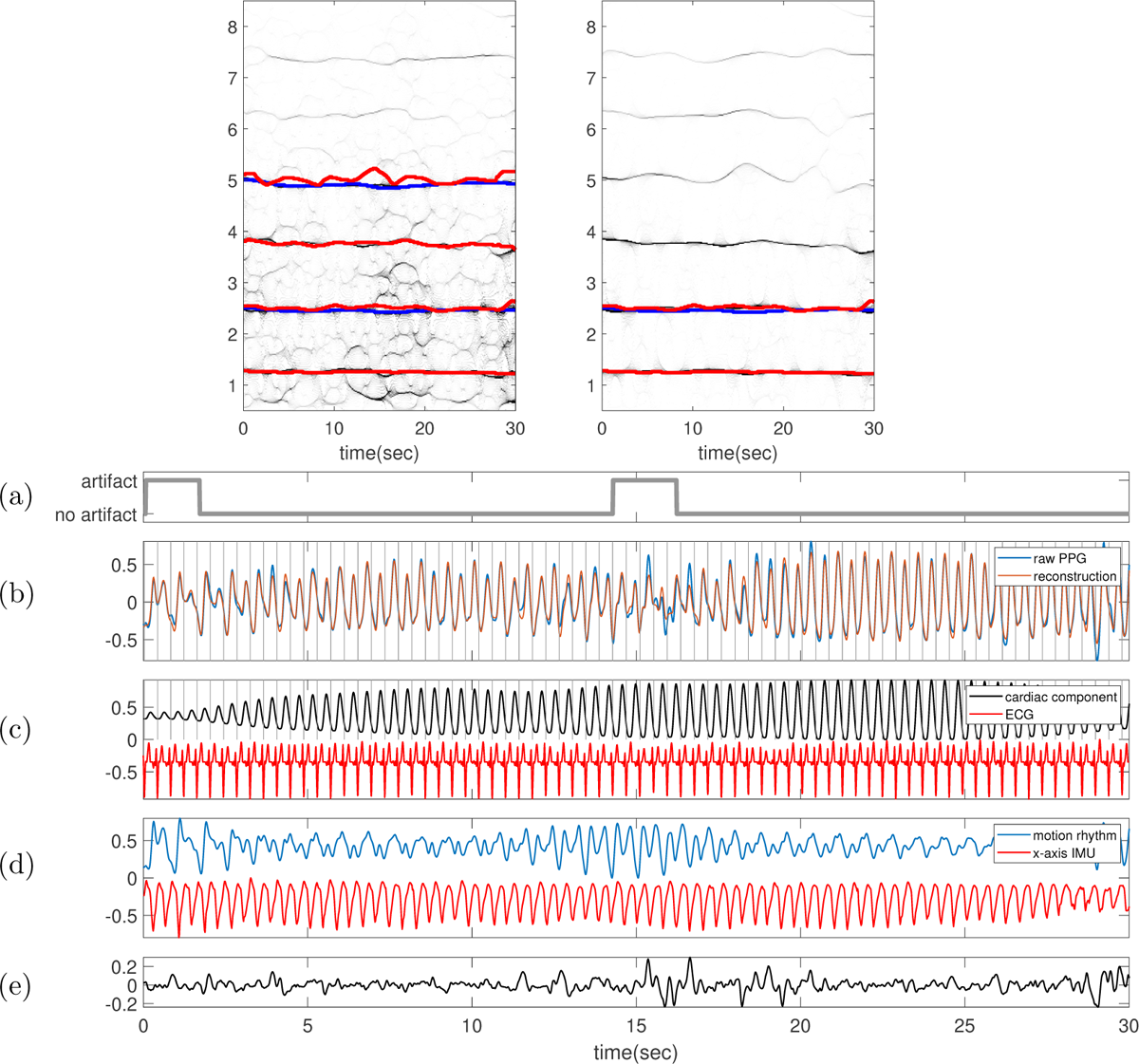
Subject 7 between 150 and 180 seconds in the TROIKA dataset. The subject is running at the speed of 6 kilometer per hour during this segment. In the top row, the TFR of the PPG with the IFs of the first two (four respectively) harmonics of ECG (IMU respectively) superimposed as blue (red respectively) curves are shown on the left-hand side. On the right-hand side, the TFR of the decomposed motion rhythm component with the IF of the first (first two respectively) harmonic of ECG (IMU respectively) superimposed as the blue (red respectively) curve are displayed. **(a)** Label sequence (grey line) and the prediction result by the proposed SQA model (red-dashed line). **(b)** The raw PPG signal is shown in black, and the summation of the decomposed motion rhythm and cardiac component is superimposed in red. The detected R-peaks from the simultaneously recorded ECG are superimposed as vertical grey lines. **(c)** The decomposed cardiac component (black curve), the simultaneously recorded ECG (red curve) and the detected R-peaks (vertical grey lines). **(d)** The decomposed motion rhythm (blue curve) and the x-axis of the accelerometer signal recorded simultaneously (red curve). **(e)** The difference of the PPG signal and the summation of the decomposed motion rhythm and cardiac component. The normalized root mean square error (NRMSE) between the PPG signal and the summation of the decomposed motion rhythm and cardiac component reconstruction is 0.20.

**Figure 26.**
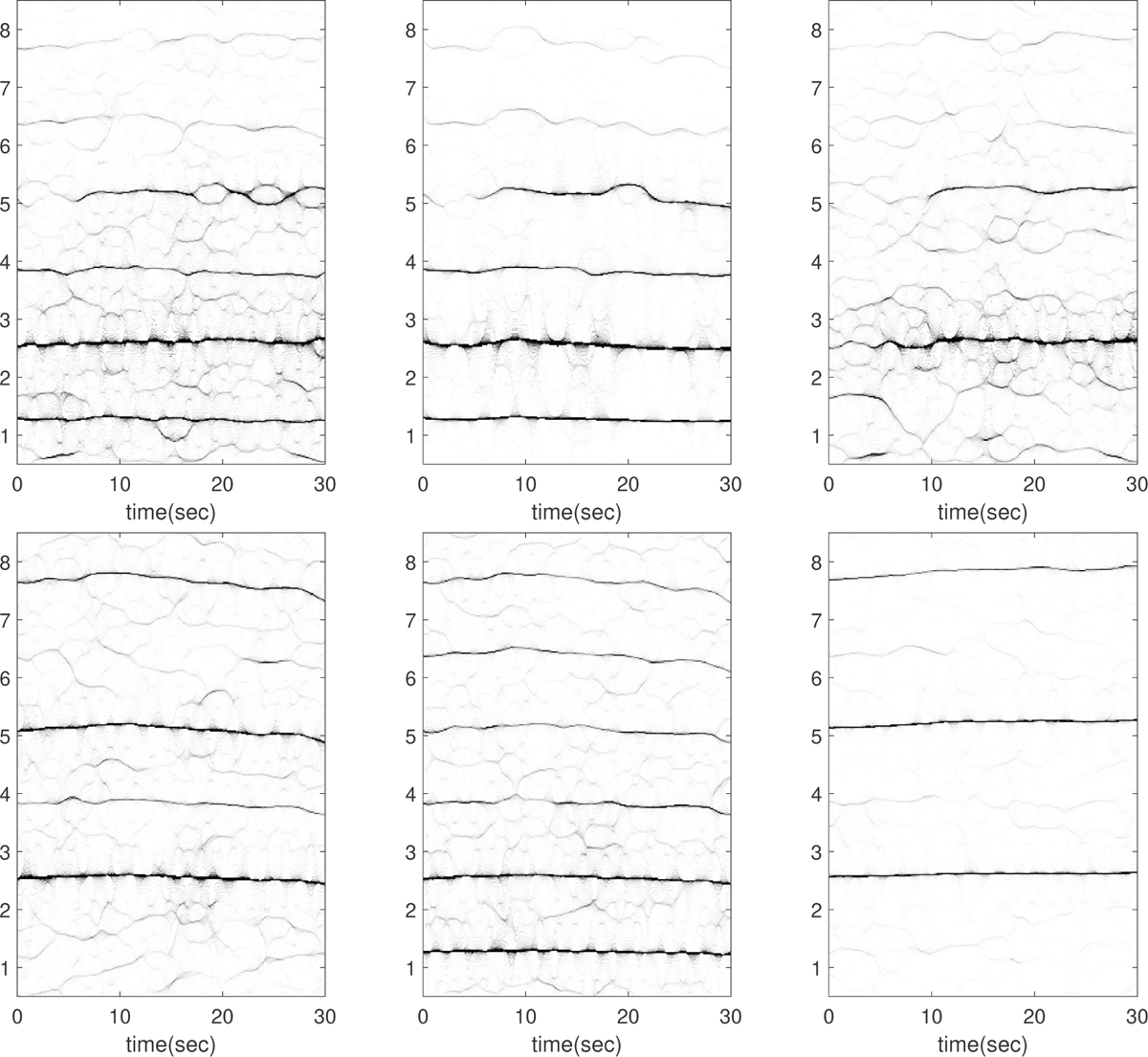
Subject 7 between 240 and 270 seconds in the TROIKA dataset. The subject is running at the speed of 12 kilometer per hour during this segment. The first row, from left to right: the time-frequency representations (TFRs) of the PPG, the decomposed motion rhythm component, and the PPG after removing the decomposed motion rhythm component. The second row, from left to right: the TFRs of the simultaneously recorded accelerometer magnitude signal, the simultaneously recorded x-axis of the accelerometer signal, and the simultaneously recorded ECG signal. Over this segment, the mean heart rate derived from the simultaneously recorded electrocardiogram (resp. raw PPG and extracted cardiac component) is 2.60 Hz (resp. 2.75 Hz, 2.62 Hz). This segment is noteworthy because the instantaneous frequency (IF) of the second harmonic of the IMU signal matches exactly with that of the fundamental component of ECG in the first 10 seconds (see the top row of Figure 27); indicating a strong 2:1 coupling between heart rate and motion rhythm. The current algorithm is limited in decomposing such signals, and the decomposition is not ideal in the first 10 seconds, as illustrated in Figure 27.

**Figure 27.**
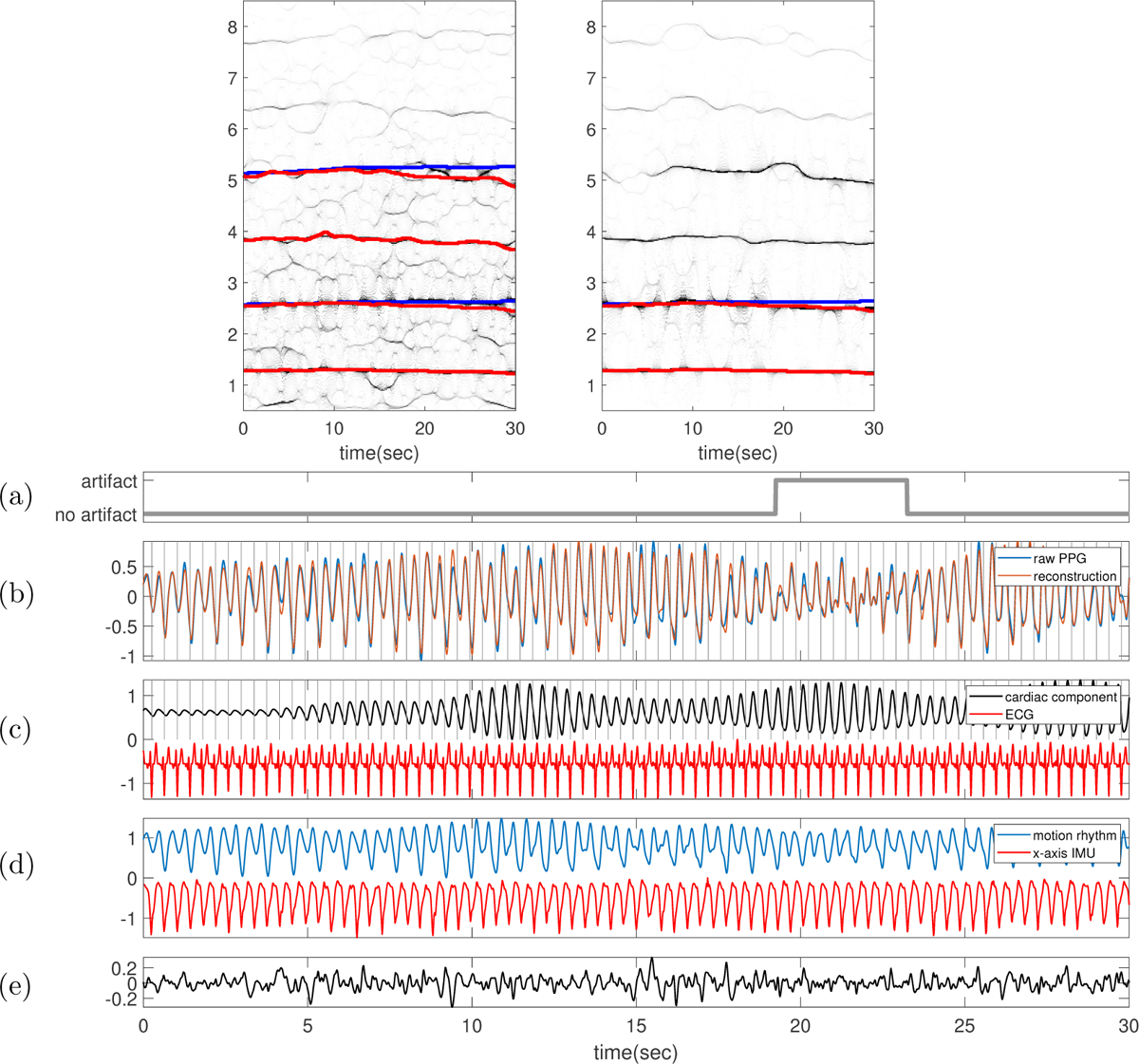
Subject 7 between 240 and 270 seconds in the TROIKA dataset. The subject is running at the speed of 12 kilometer per hour during this segment. In the top row, the TFR of the PPG with the IFs of the first two (four respectively) harmonics of ECG (IMU respectively) superimposed as blue (red respectively) curves are shown on the left-hand side. On the right-hand side, the TFR of the decomposed motion rhythm component with the IF of the first (first two respectively) harmonic of ECG (IMU respectively) superimposed as the blue (red respectively) curve are displayed. **(a)** Label sequence (grey line) and the prediction result by the proposed SQA model (red-dashed line). **(b)** The raw PPG signal is shown in black, and the summation of the decomposed motion rhythm and cardiac component is superimposed in red. The detected R-peaks from the simultaneously recorded ECG are superimposed as vertical grey lines. **(c)** The decomposed cardiac component (black curve), the simultaneously recorded ECG (red curve) and the detected R-peaks (vertical grey lines). **(d)** The decomposed motion rhythm (blue curve) and the x-axis of the accelerometer signal recorded simultaneously (red curve). **(e)** The difference of the PPG signal and the summation of the decomposed motion rhythm and cardiac component. The normalized root mean square error (NRMSE) between the PPG signal and the summation of the decomposed motion rhythm and cardiac component reconstruction is 0.20.

**Figure 28.**
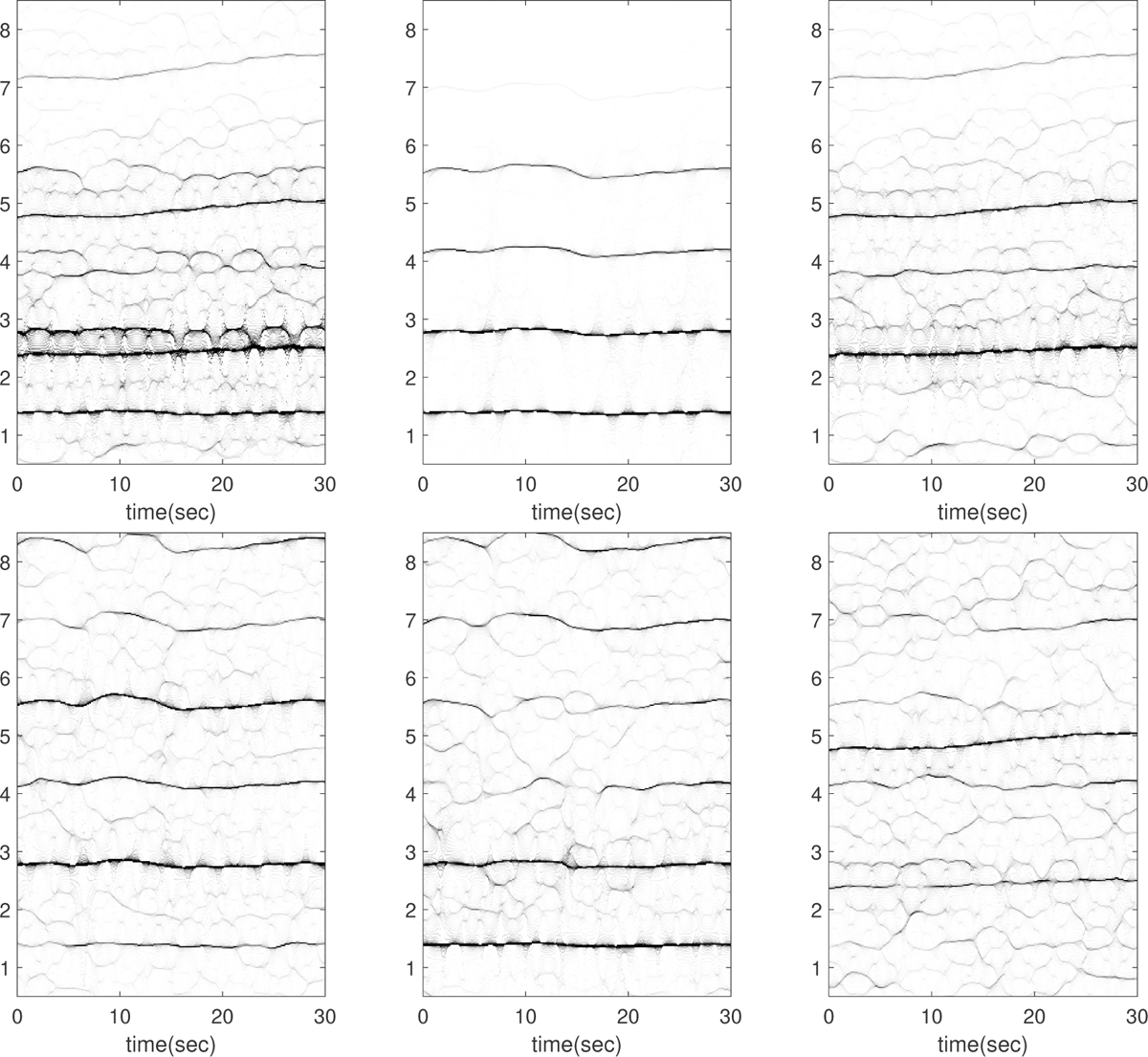
Subject 9 between 240 and 270 seconds in the TROIKA dataset. The subject is running at the speed of 12 kilometer per hour during this segment. The first row, from left to right: the time-frequency representations (TFRs) of the PPG, the decomposed motion rhythm component, and the PPG after removing the decomposed motion rhythm component. The second row, from left to right: the TFRs of the simultaneously recorded accelerometer magnitude signal, the simultaneously recorded x-axis of the accelerometer signal, and the simultaneously recorded ECG signal. Over this segment, the mean heart rate derived from the simultaneously recorded electrocardiogram (resp. raw PPG and extracted cardiac component) is 2.45 Hz (resp. 2.57 Hz, 2.44 Hz). From the TFR of the ECG, there are plausible components oscillating at *∼*2.8 Hz, *∼*4.2 Hz and *∼*5.5 Hz in the ECG, which coincide with the second, third and fourth harmonics of the motion rhythm.

**Figure 29.**
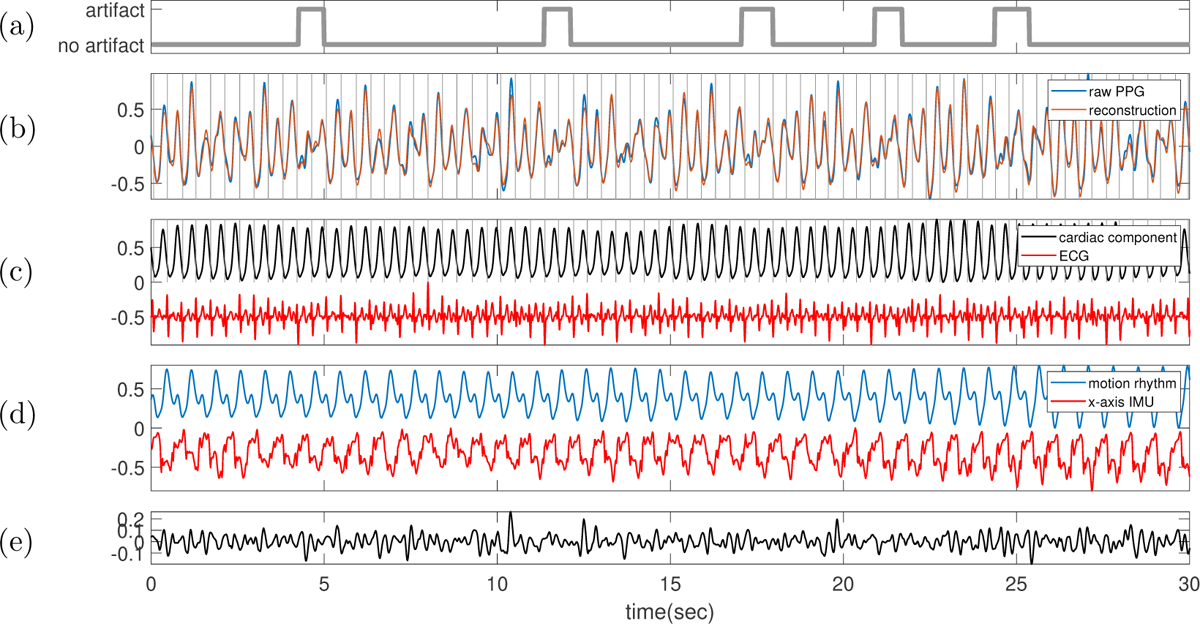
Subject 9 between 240 and 270 seconds in the TROIKA dataset. The subject is running at the speed of 12 kilometer per hour during this segment. **(a)** Label sequence (grey line) and the prediction result by the proposed SQA model (red-dashed line). **(b)** The raw PPG signal is shown in black, and the summation of the decomposed motion rhythm and cardiac component is superimposed in red. The detected R-peaks from the simultaneously recorded ECG are superimposed as vertical grey lines. **(c)** The decomposed cardiac component (black curve), the simultaneously recorded ECG (red curve) and the detected R-peaks (vertical grey lines). **(d)** The decomposed motion rhythm (blue curve) and the x-axis of the accelerometer signal recorded simultaneously (red curve). **(e)** The difference of the PPG signal and the summation of the decomposed motion rhythm and cardiac component. The normalized root mean square error (NRMSE) between the PPG signal and the summation of the decomposed motion rhythm and cardiac component reconstruction is 0.18.

**Figure 30.**
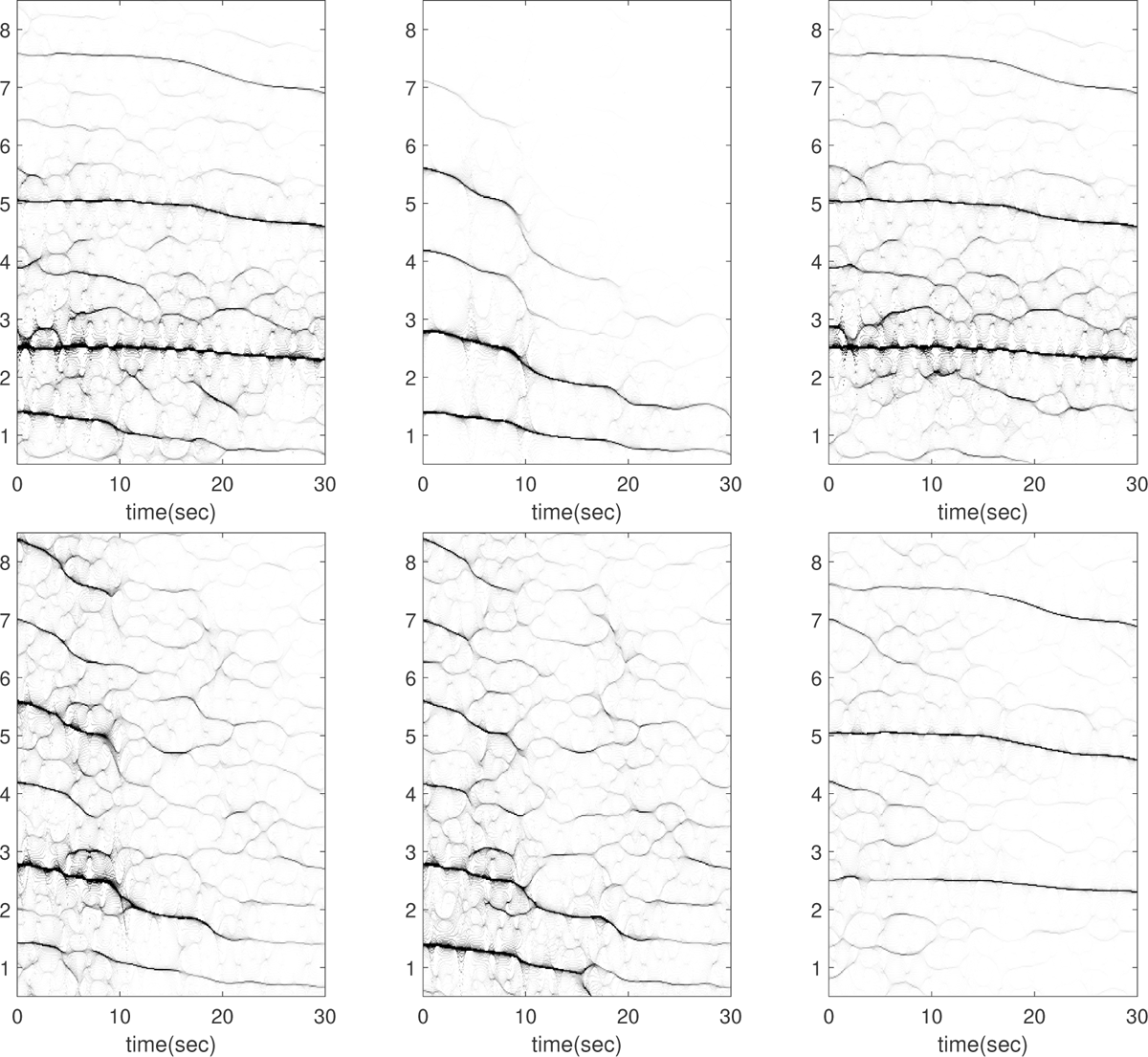
Subject 9 between 270 and 300 seconds in the TROIKA dataset. The subject is resting during this segment. The first row, from left to right: the time-frequency representations (TFRs) of the PPG, the decomposed motion rhythm component, and the PPG after removing the decomposed motion rhythm component. The second row, from left to right: the TFRs of the simultaneously recorded accelerometer magnitude signal, the simultaneously recorded x-axis of the accelerometer signal, and the simultaneously recorded ECG signal. Over this segment, the mean heart rate derived from the simultaneously recorded electrocardiogram (resp. raw PPG and extracted cardiac component) is 2.44 Hz (resp. 2.46 Hz, 2.45 Hz). The instantaneous frequency of the accelerometer magnitude signal (in the PPG signal as well) in this case decreases from *∼* 1.4 Hz to *∼* 0.7 Hz, and the amplitude modulation (strength) also decreases. While we lack detailed information about the recording environment, we hypothesize that the subject may have decreased their running speed, possibly transitioning to a walking mode, during the recording of this segment.

**Figure 31.**
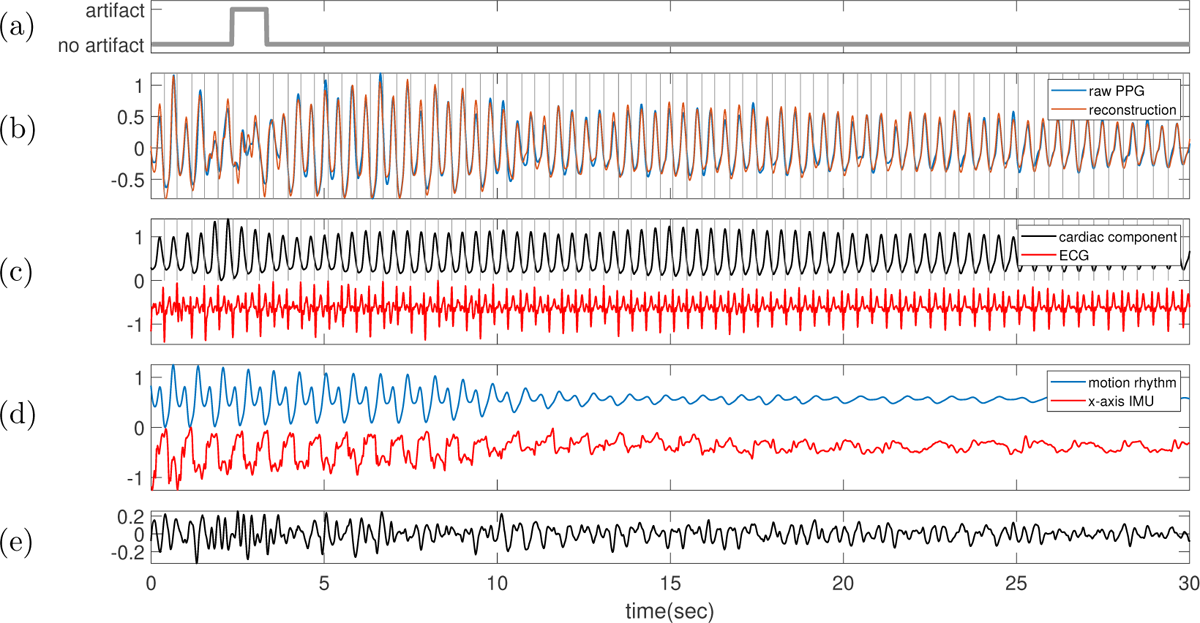
Subject 9 between 270 and 300 seconds in the TROIKA dataset. The subject is resting during this segment. **(a)** Label sequence (grey line) and the prediction result by the proposed SQA model (red-dashed line). **(b)** The raw PPG signal is shown in black, and the summation of the decomposed motion rhythm and cardiac component is superimposed in red. The detected R-peaks from the simultaneously recorded ECG are superimposed as vertical grey lines. **(c)** The decomposed cardiac component (black curve), the simultaneously recorded ECG (red curve) and the detected R-peaks (vertical grey lines). **(d)** The decomposed motion rhythm (blue curve) and the x-axis of the accelerometer signal recorded simultaneously (red curve). **(e)** The difference of the PPG signal and the summation of the decomposed motion rhythm and cardiac component. The normalized root mean square error (NRMSE) between the PPG signal and the summation of the decomposed motion rhythm and cardiac component reconstruction is 0.23. Note that the instantaneous frequency and amplitude modulation of the motion rhythm in the PPG and the accelerometer magnitude signal both decrease since the 10th second. While we lack detailed information about the recording environment, we hypothesize that the subject may have decreased their running speed, possibly transitioning to a walking mode, during the recording of this segment.

**Figure 32.**
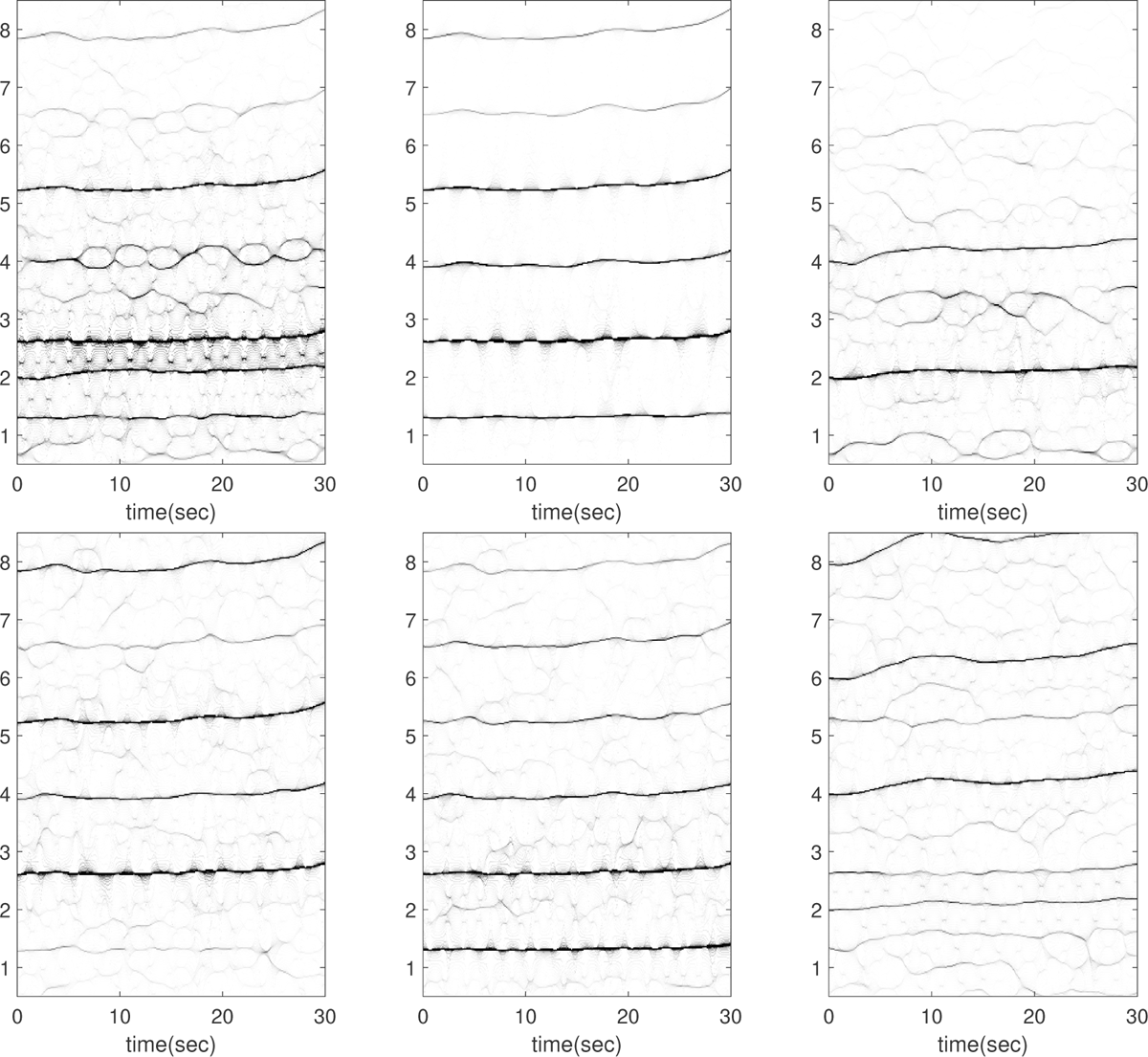
Subject 12 between 60 and 90 seconds in the TROIKA dataset. The subject is running at the speed of 6 kilometer per hour during this segment. The first row, from left to right: the time-frequency representations (TFRs) of the PPG, the decomposed motion rhythm component, and the PPG after removing the decomposed motion rhythm component. The second row, from left to right: the TFRs of the simultaneously recorded accelerometer magnitude signal, the simultaneously recorded x-axis of the accelerometer signal, and the simultaneously recorded ECG signal. Over this segment, the mean heart rate derived from the simultaneously recorded electrocardiogram (resp. raw PPG and extracted cardiac component) is 2.11 Hz (resp. 2.66 Hz, 2.10 Hz). In this case, there are plausible components oscillating at approximately 2.7 Hz and 5.3 Hz in the ECG, aligning with the second and fourth harmonics of the motion rhythm. While the presence of the third harmonic is conceivable, it may not be visible as it overlaps with the second harmonic of the ECG signal and is overshadowed in strength.

**Figure 33.**
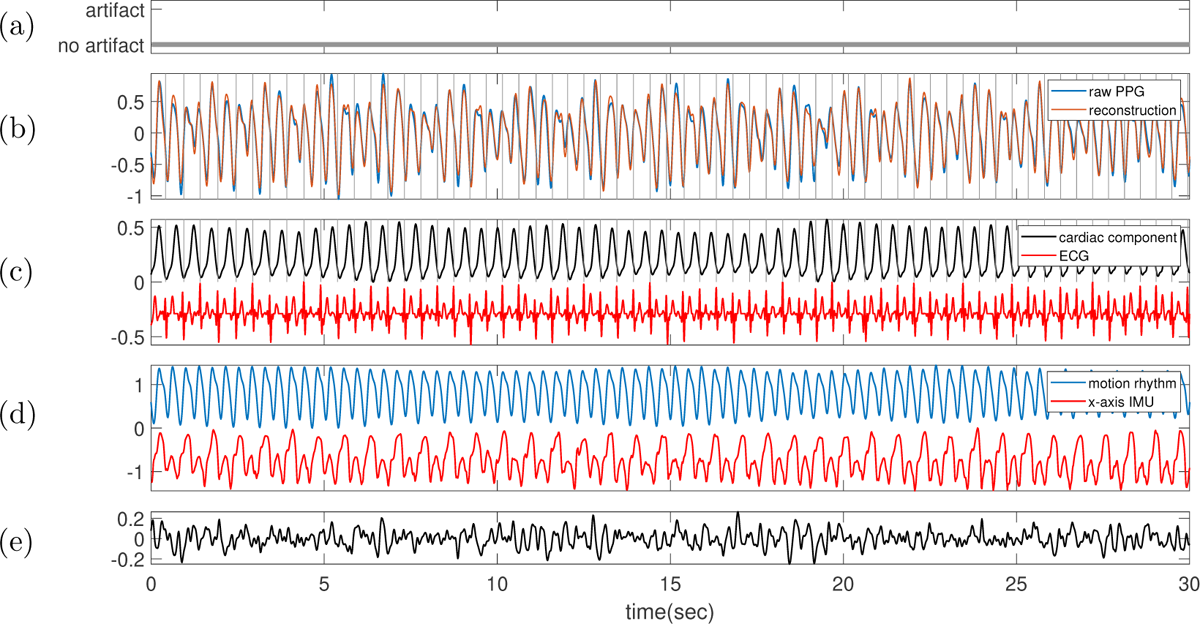
Subject 12 between 60 and 90 seconds in the TROIKA dataset. The subject is running at the speed of 6 kilometer per hour during this segment. **(a)** Label sequence (grey line) and the prediction result by the proposed SQA model (red-dashed line). **(b)** The raw PPG signal is shown in black, and the summation of the decomposed motion rhythm and cardiac component is superimposed in red. The detected R-peaks from the simultaneously recorded ECG are superimposed as vertical grey lines. **(c)** The decomposed cardiac component (black curve), the simultaneously recorded ECG (red curve) and the detected R-peaks (vertical grey lines). **(d)** The decomposed motion rhythm (blue curve) and the x-axis of the accelerometer signal recorded simultaneously (red curve). **(e)** The difference of the PPG signal and the summation of the decomposed motion rhythm and cardiac component. The normalized root mean square error (NRMSE) between the PPG signal and the summation of the decomposed motion rhythm and cardiac component reconstruction is 0.18.

**Figure 34.**
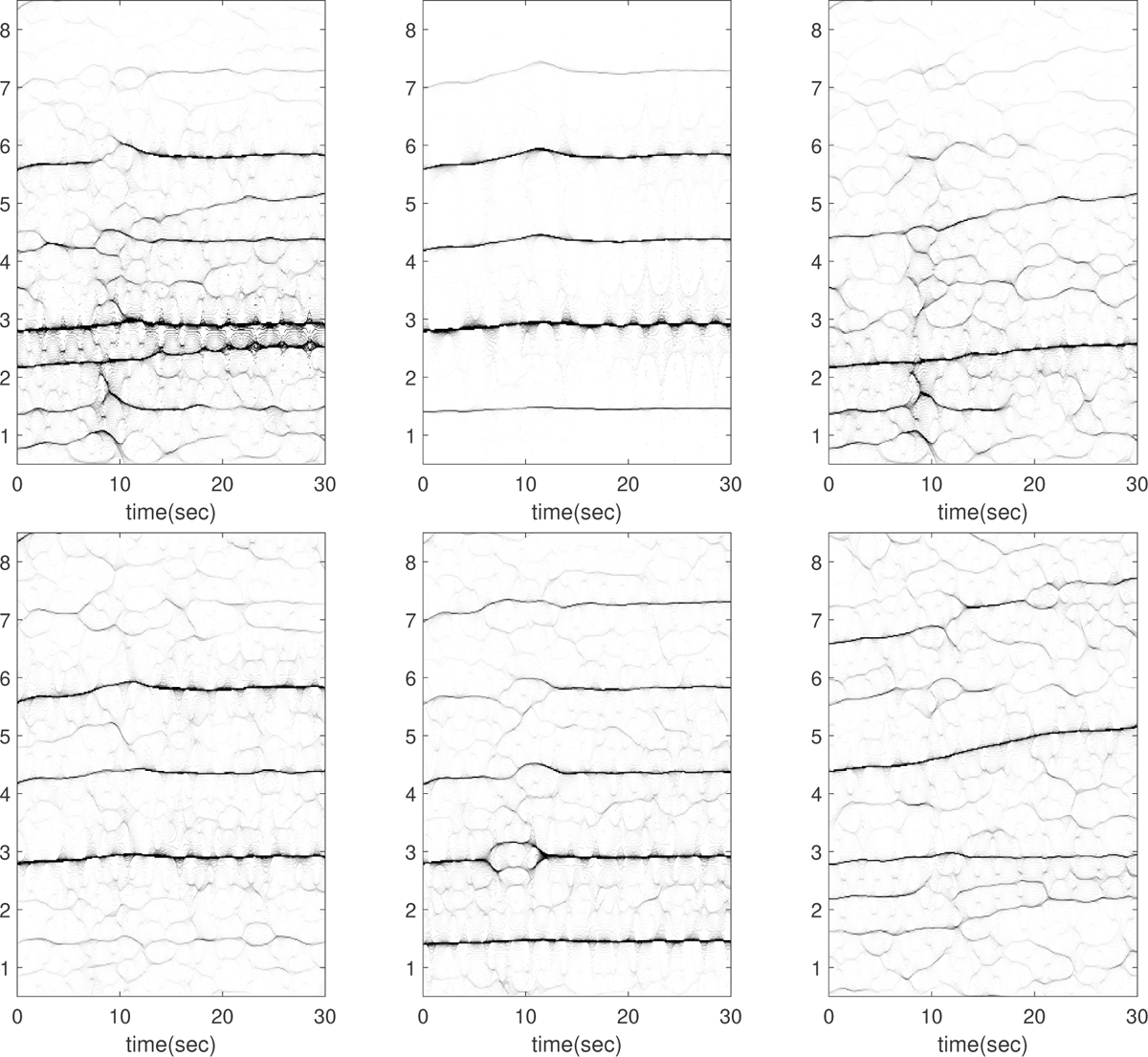
Subject 12 between 90 and 120 seconds in the TROIKA dataset. The subject is running at the speed of 12 kilometer per hour during this segment. The first row, from left to right: the time-frequency representations (TFRs) of the PPG, the decomposed motion rhythm component, and the PPG after removing the decomposed motion rhythm component. The second row, from left to right: the TFRs of the simultaneously recorded accelerometer magnitude signal, the simultaneously recorded x-axis of the accelerometer signal, and the simultaneously recorded ECG signal. Over this segment, the mean heart rate derived from the simultaneously recorded electrocardiogram (resp. raw PPG and extracted cardiac component) is 2.40 Hz (resp. 2.90 Hz, 2.39 Hz). The TFR of the ECG signal presents an intriguing observation: the fundamental component of the ECG is affected by contamination from the second harmonic of the motion rhythm. This contamination could potentially mislead one to conclude that the instantaneous frequency of the second harmonic of the ECG deviates from twice the instantaneous frequency of the fundamental component.

**Figure 35.**
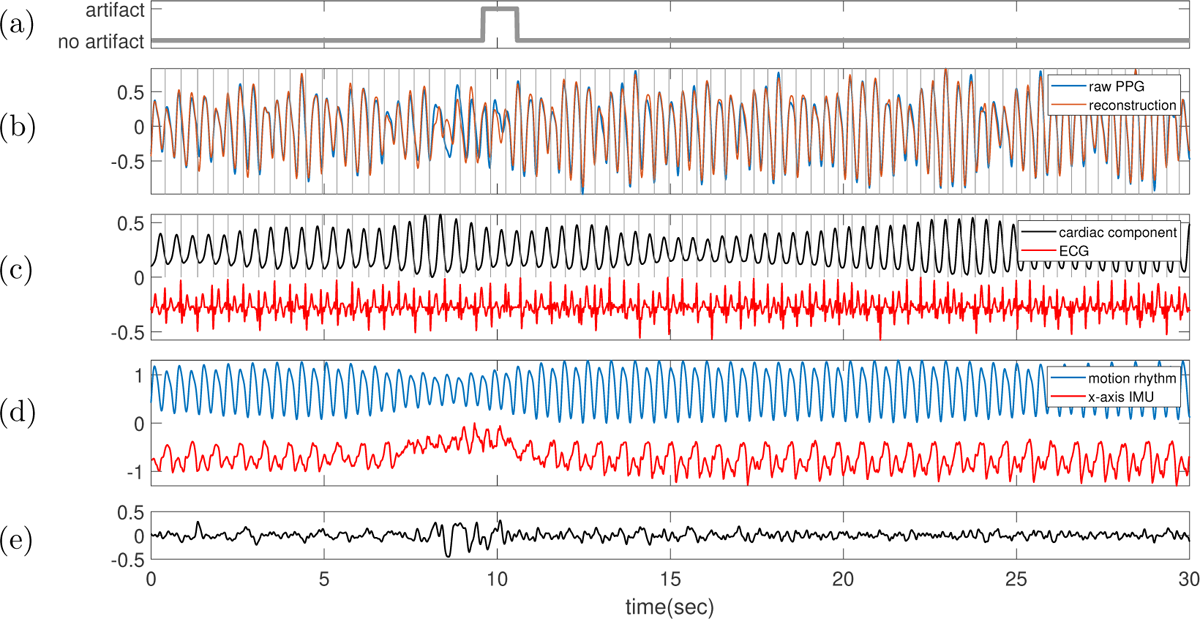
Subject 12 between 90 and 120 seconds in the TROIKA dataset. The subject is running at the speed of 12 kilometer per hour during this segment. **(a)** Label sequence (grey line) and the prediction result by the proposed SQA model (red-dashed line). **(b)** The raw PPG signal is shown in black, and the summation of the decomposed motion rhythm and cardiac component is superimposed in red. The detected R-peaks from the simultaneously recorded ECG are superimposed as vertical grey lines. **(c)** The decomposed cardiac component (black curve), the simultaneously recorded ECG (red curve) and the detected R-peaks (vertical grey lines). **(d)** The decomposed motion rhythm (blue curve) and the x-axis of the accelerometer signal recorded simultaneously (red curve). **(e)** The difference of the PPG signal and the summation of the decomposed motion rhythm and cardiac component. The normalized root mean square error (NRMSE) between the PPG signal and the summation of the decomposed motion rhythm and cardiac component reconstruction is 0.22.

**Figure 36.**
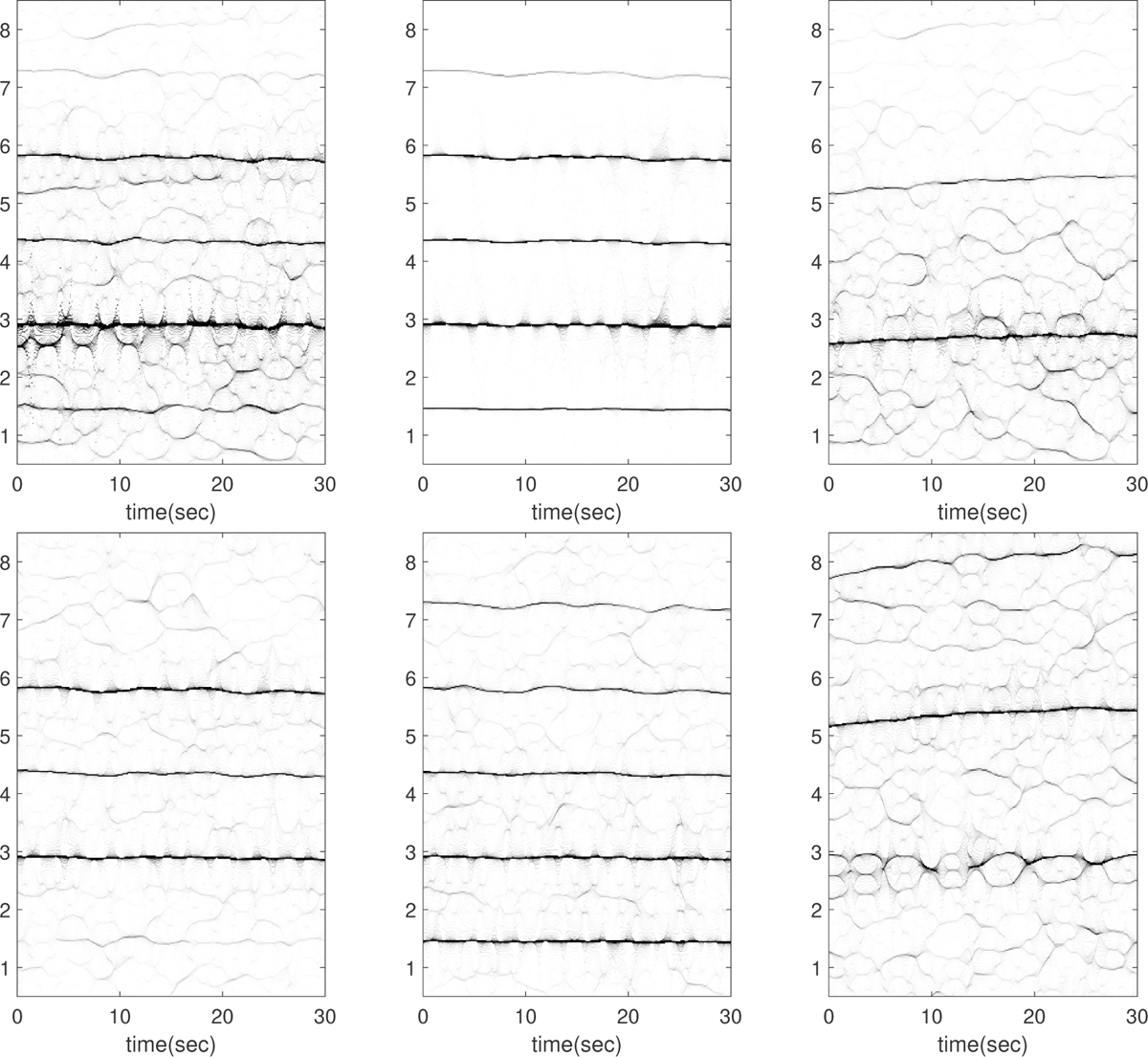
Subject 12 between 120 and 150 seconds in the TROIKA dataset. The subject is running at the speed of 12 kilometer per hour during this segment. The first row, from left to right: the time-frequency representations (TFRs) of the PPG, the decomposed motion rhythm component, and the PPG after removing the decomposed motion rhythm component. The second row, from left to right: the TFRs of the simultaneously recorded accelerometer magnitude signal, the simultaneously recorded x-axis of the accelerometer signal, and the simultaneously recorded ECG signal. Over this segment, the mean heart rate derived from the simultaneously recorded electrocardiogram (resp. raw PPG and extracted cardiac component) is 2.67 Hz (resp. 2.97 Hz, 2.68 Hz). In the TFR of the ECG signal, we can see a dominant pattern around 2.8–3Hz, which possibly comes from the interference of the second harmonic of motion rhythm and the fundamental component of the ECG signal.

**Figure 37.**
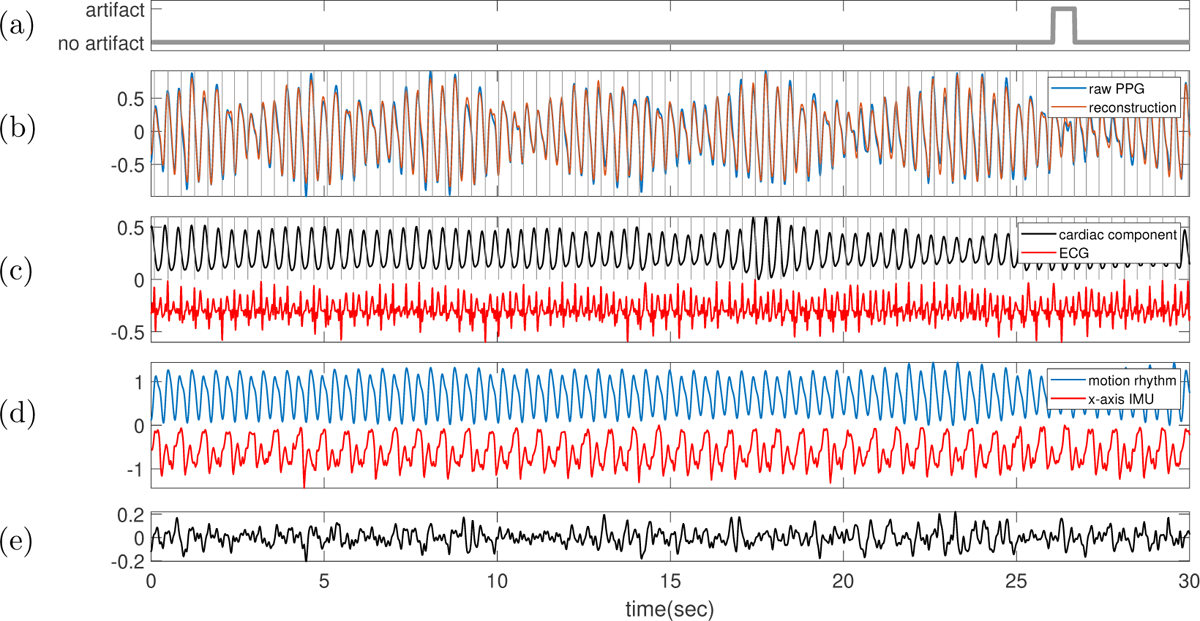
Subject 12 between 120 and 150 seconds in the TROIKA dataset. The subject is running at the speed of 12 kilometer per hour during this segment. **(a)** Label sequence (grey line) and the prediction result by the proposed SQA model (red-dashed line). **(b)** The raw PPG signal is shown in black, and the summation of the decomposed motion rhythm and cardiac component is superimposed in red. The detected R-peaks from the simultaneously recorded ECG are superimposed as vertical grey lines. **(c)** The decomposed cardiac component (black curve), the simultaneously recorded ECG (red curve) and the detected R-peaks (vertical grey lines). **(d)** The decomposed motion rhythm (blue curve) and the x-axis of the accelerometer signal recorded simultaneously (red curve). **(e)** The difference of the PPG signal and the summation of the decomposed motion rhythm and cardiac component. The normalized root mean square error (NRMSE) between the PPG signal and the summation of the decomposed motion rhythm and cardiac component reconstruction is 0.16.

**Figure 38.**
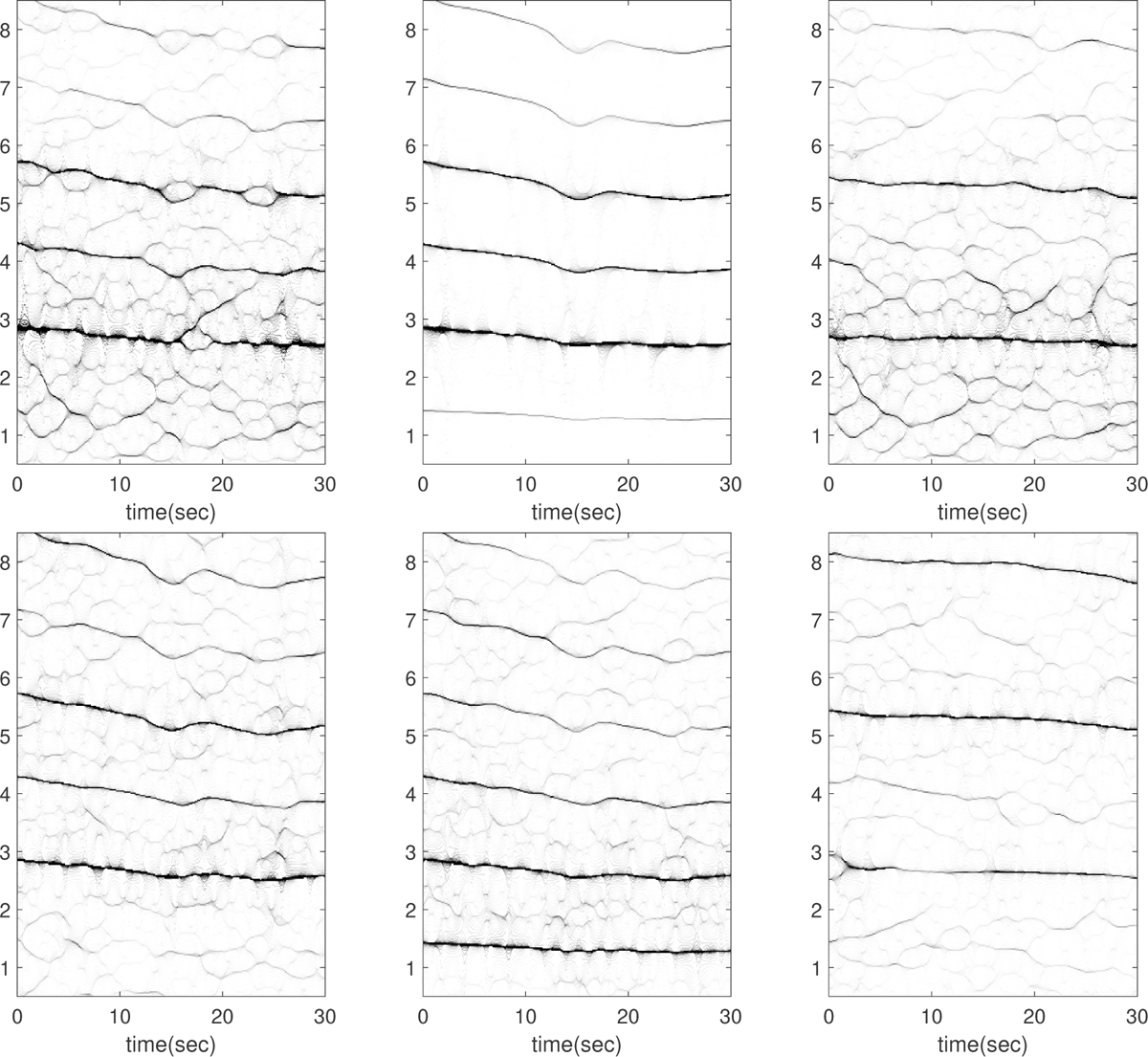
Subject 12 between 150 and 180 seconds in the TROIKA dataset. The subject is running at the speed of 6 kilometer per hour during this segment. The first row, from left to right: the time-frequency representations (TFRs) of the PPG, the decomposed motion rhythm component, and the PPG after removing the decomposed motion rhythm component. The second row, from left to right: the TFRs of the simultaneously recorded accelerometer magnitude signal, the simultaneously recorded x-axis of the accelerometer signal, and the simultaneously recorded ECG signal. Over this segment, the mean heart rate derived from the simultaneously recorded electrocardiogram (resp. raw PPG and extracted cardiac component) is 2.65 Hz (resp. 3.07 Hz, 2.65 Hz). In this case, the fundamental component of the accelerometer magnitude signal cannot be visualized. Moreover, this segment displays an interesting “frequency crossover” phenomenon in addition to having similar instantaneous frequency (IFs); that is, the IF of the second harmonic of the IMU signal decreases and cross that of the fundamental component of ECG at around the 10th second (see the top row of Figure 39). This is different from the 2:1 coupling between heart rate and motion rhythm. The current algorithm is also limited in decomposing such signals, and the decomposition is not ideal around the 10th seconds, as illustrated in Figure 39.

**Figure 39.**
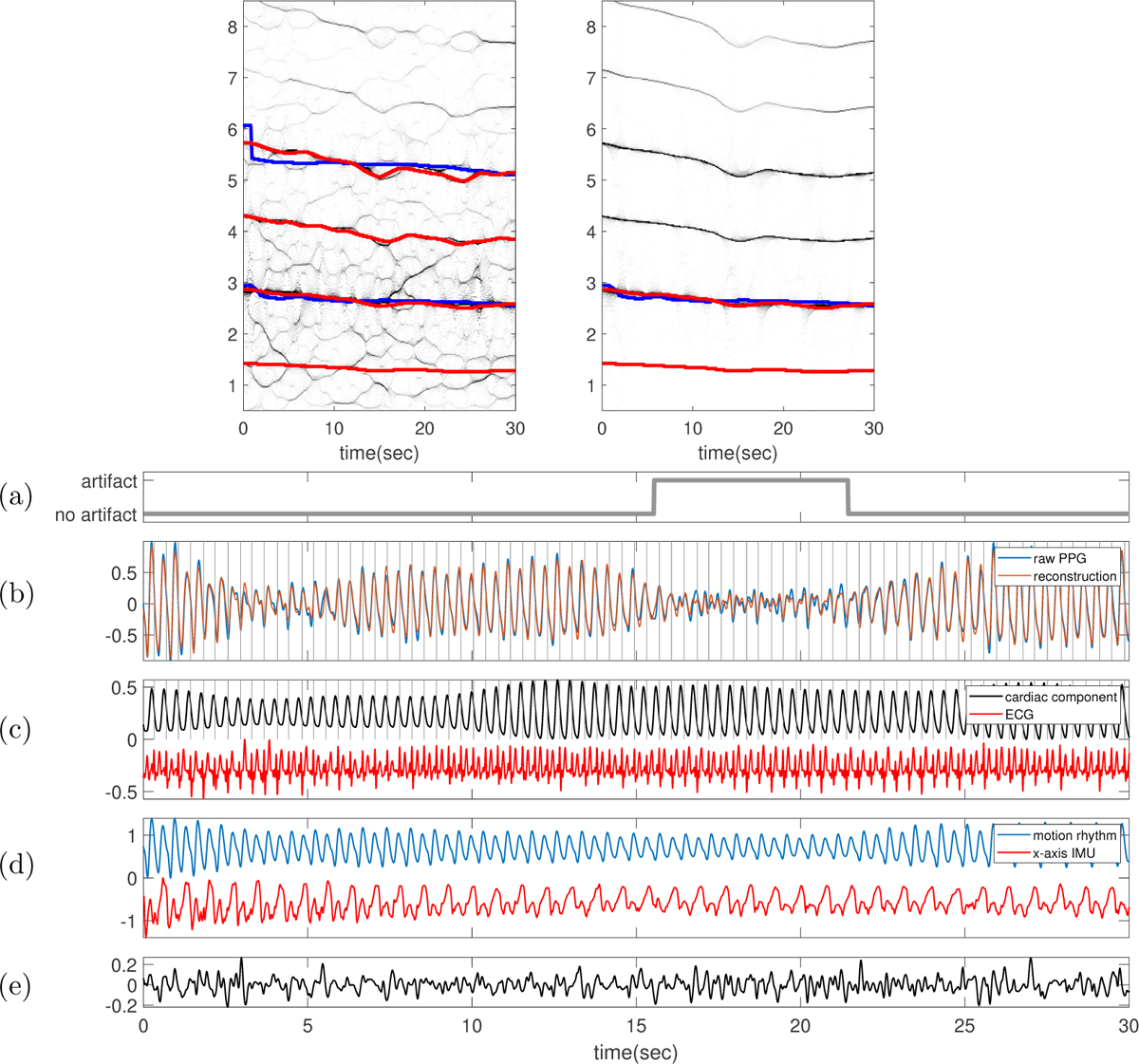
Subject 12 between 150 and 180 seconds in the TROIKA dataset. The subject is running at the speed of 6 kilometer per hour during this segment. In the top row, the TFR of the PPG with the IFs of the first two (four respectively) harmonics of ECG (IMU respectively) superimposed as blue (red respectively) curves are shown on the left-hand side. On the right-hand side, the TFR of the decomposed motion rhythm component with the IF of the first (first two respectively) harmonic of ECG (IMU respectively) superimposed as the blue (red respectively) curve are displayed. **(a)** Label sequence (grey line) and the prediction result by the proposed SQA model (red-dashed line). **(b)** The raw PPG signal is shown in black, and the summation of the decomposed motion rhythm and cardiac component is superimposed in red. The detected R-peaks from the simultaneously recorded ECG are superimposed as vertical grey lines. **(c)** The decomposed cardiac component (black curve), the simultaneously recorded ECG (red curve) and the detected R-peaks (vertical grey lines). **(d)** The decomposed motion rhythm (blue curve) and the x-axis of the accelerometer signal recorded simultaneously (red curve). **(e)** The difference of the PPG signal and the summation of the decomposed motion rhythm and cardiac component. The normalized root mean square error (NRMSE) between the PPG signal and the summation of the decomposed motion rhythm and cardiac component reconstruction is 0.20.

**Figure 40.**
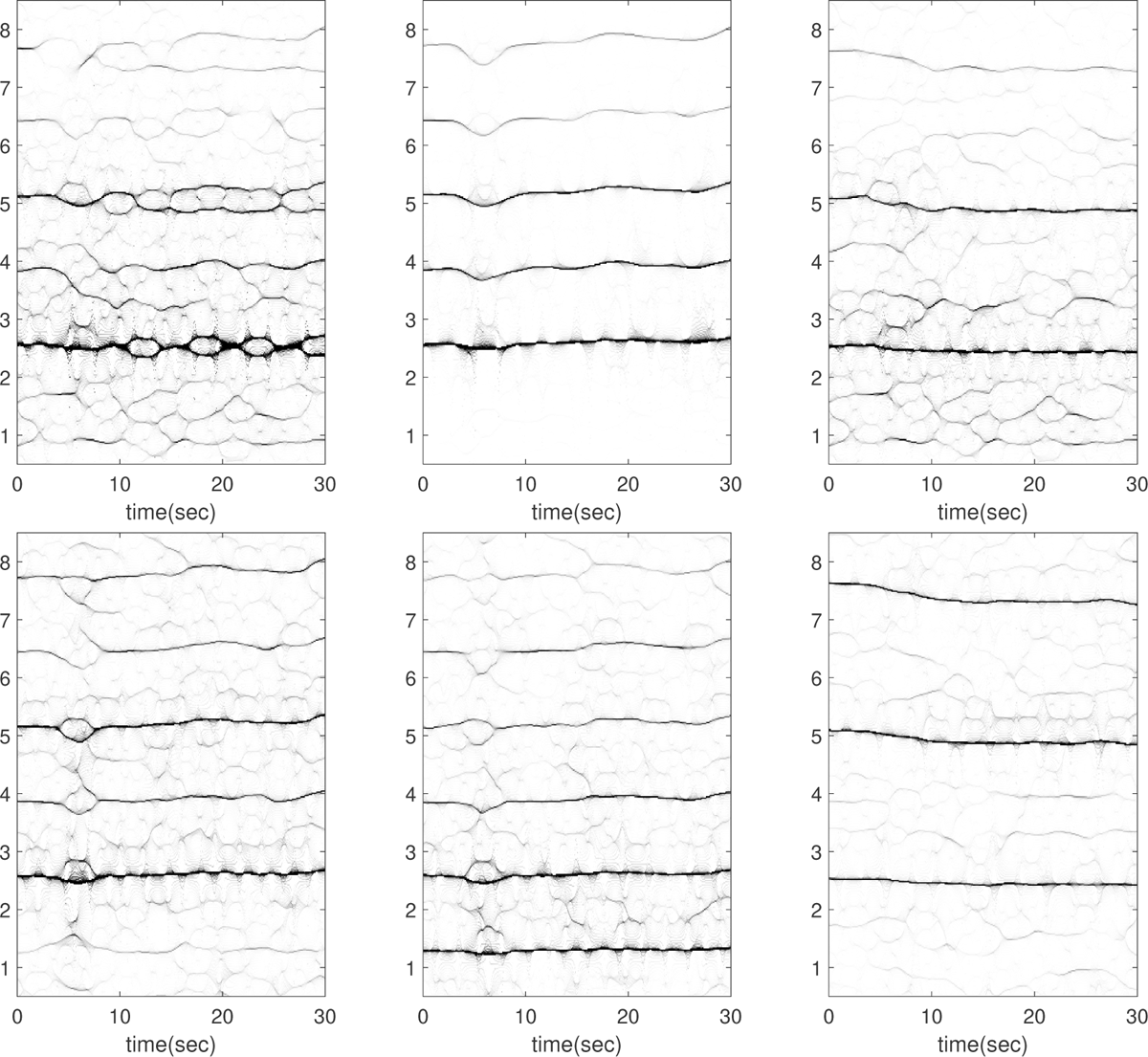
Subject 12 between 180 and 210 seconds in the TROIKA dataset. The subject is running at the speed of 6 kilometer per hour during this segment. The first row, from left to right: the time-frequency representations (TFRs) of the PPG, the decomposed motion rhythm component, and the PPG after removing the decomposed motion rhythm component. The second row, from left to right: the TFRs of the simultaneously recorded accelerometer magnitude signal, the simultaneously recorded x-axis of the accelerometer signal, and the simultaneously recorded ECG signal. Over this segment, the mean heart rate derived from the simultaneously recorded electrocardiogram (resp. raw PPG and extracted cardiac component) is 2.47 Hz (resp. 2.84 Hz, 2.54 Hz). This segment is noteworthy because the instantaneous frequency (IF) of the second harmonic of the IMU signal matches exactly with that of the fundamental component of ECG in the first 5 seconds (see the top row of Figure 41); indicating a strong 2:1 coupling between heart rate and motion rhythm. The current algorithm is limited in decomposing such signals, and the decomposition is not ideal in the first 10 seconds, as illustrated in Figure 41.

**Figure 41.**
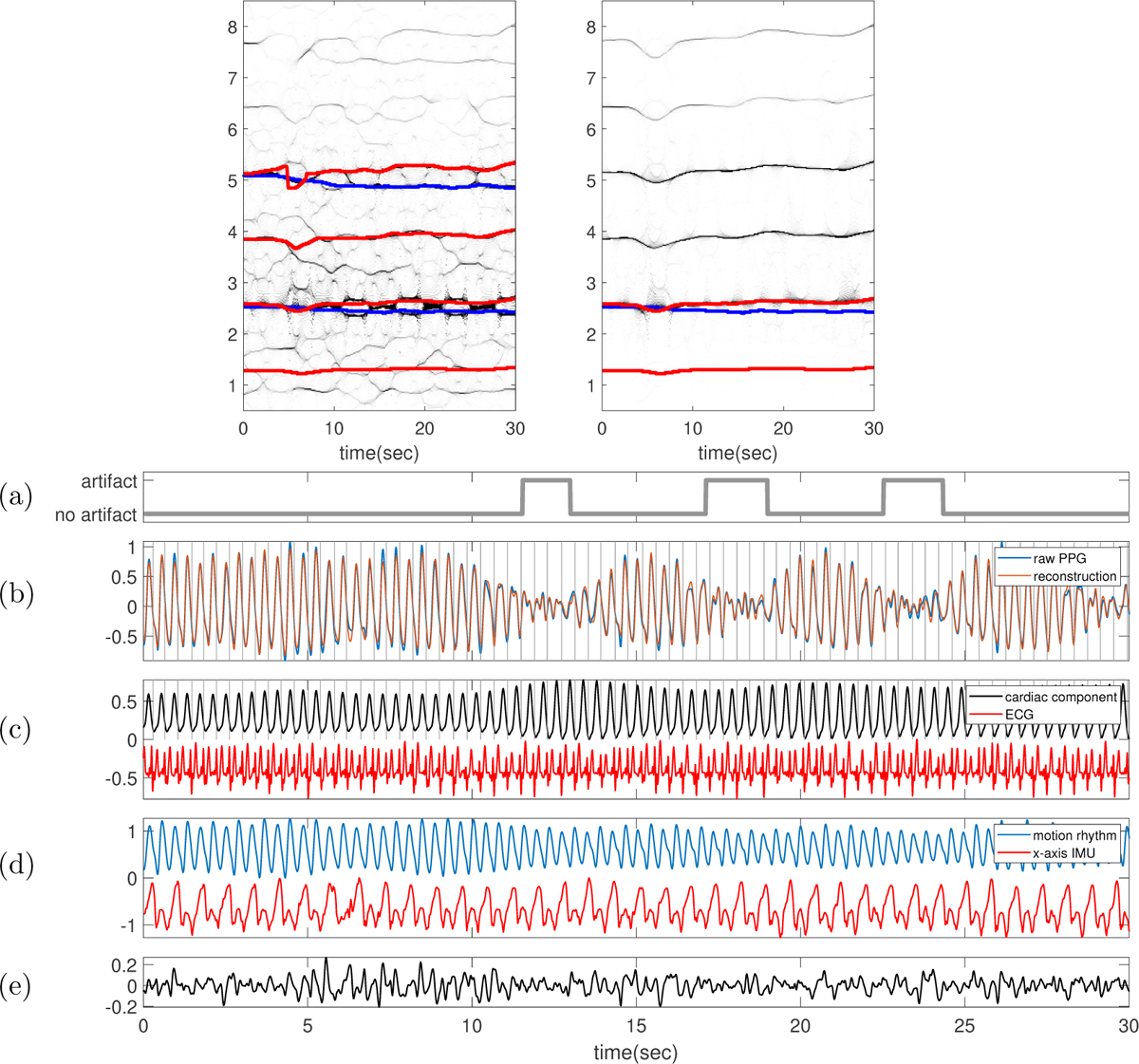
Subject 12 between 180 and 210 seconds in the TROIKA dataset. The subject is running at the speed of 6 kilometer per hour during this segment. In the top row, the TFR of the PPG with the IFs of the first two (four respectively) harmonics of ECG (IMU respectively) superimposed as blue (red respectively) curves are shown on the left-hand side. On the right-hand side, the TFR of the decomposed motion rhythm component with the IF of the first (first two respectively) harmonic of ECG (IMU respectively) superimposed as the blue (red respectively) curve are displayed. **(a)** Label sequence (grey line) and the prediction result by the proposed SQA model (red-dashed line). **(b)** The raw PPG signal is shown in black, and the summation of the decomposed motion rhythm and cardiac component is superimposed in red. The detected R-peaks from the simultaneously recorded ECG are superimposed as vertical grey lines. **(c)** The decomposed cardiac component (black curve), the simultaneously recorded ECG (red curve) and the detected R-peaks (vertical grey lines). **(d)** The decomposed motion rhythm (blue curve) and the x-axis of the accelerometer signal recorded simultaneously (red curve). **(e)** The difference of the PPG signal and the summation of the decomposed motion rhythm and cardiac component. The normalized root mean square error (NRMSE) between the PPG signal and the summation of the decomposed motion rhythm and cardiac component reconstruction is 0.16.

**Figure 42.**
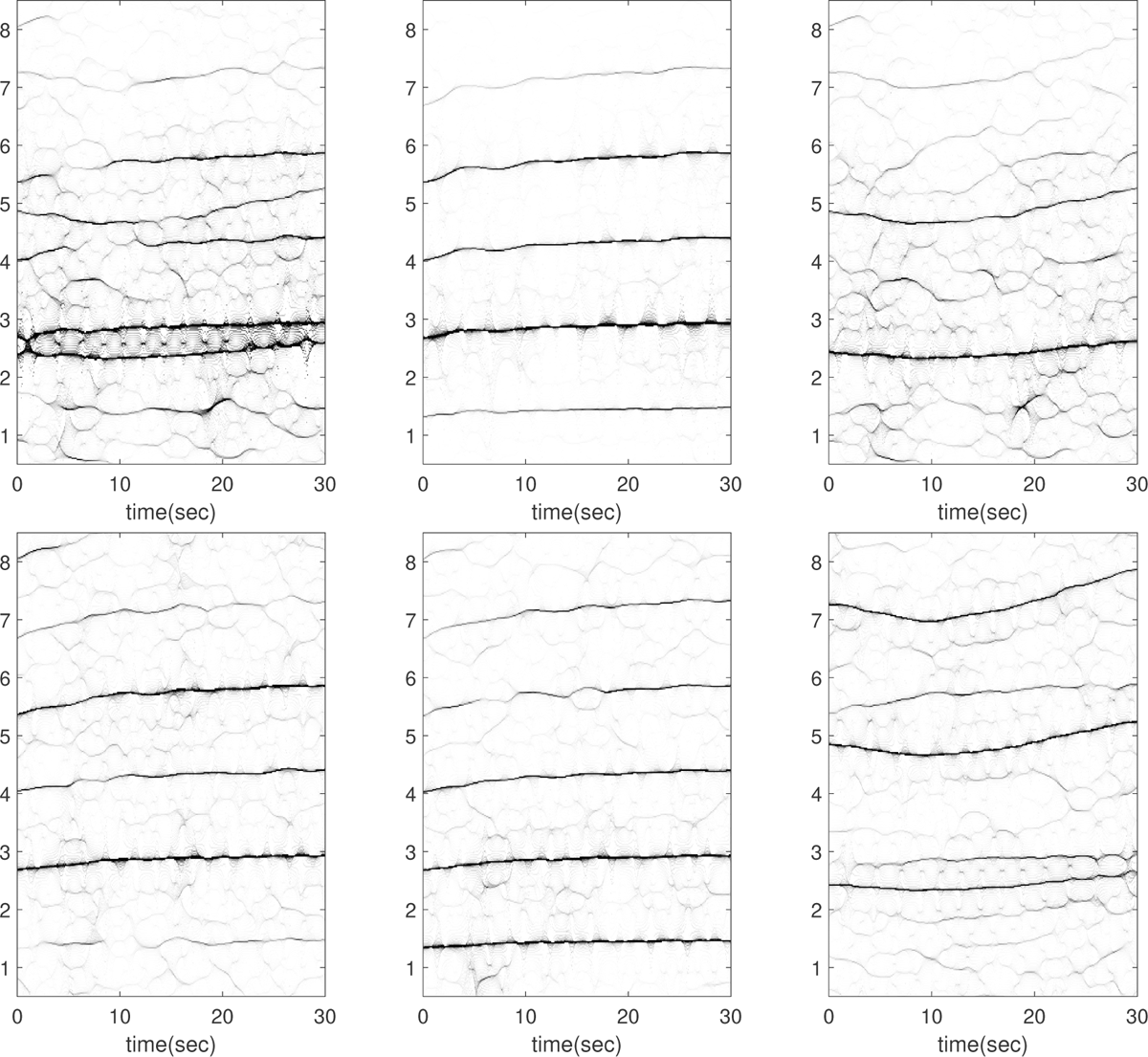
Subject 12 between 210 and 240 seconds in the TROIKA dataset. The subject is running at the speed of 12 kilometer per hour during this segment. The first row, from left to right: the time-frequency representations (TFRs) of the PPG, the decomposed motion rhythm component, and the PPG after removing the decomposed motion rhythm component. The second row, from left to right: the TFRs of the simultaneously recorded accelerometer magnitude signal, the simultaneously recorded x-axis of the accelerometer signal, and the simultaneously recorded ECG signal. Over this segment, the mean heart rate derived from the simultaneously recorded electrocardiogram (resp. raw PPG and extracted cardiac component) is 2.47 Hz (resp. 2.95 Hz, 2.43 Hz). In this case, there are plausible components oscillating at approximately 2.9 Hz and 5.6 Hz in the ECG, aligning with the second and fourth harmonics of the motion rhythm.

**Figure 43.**
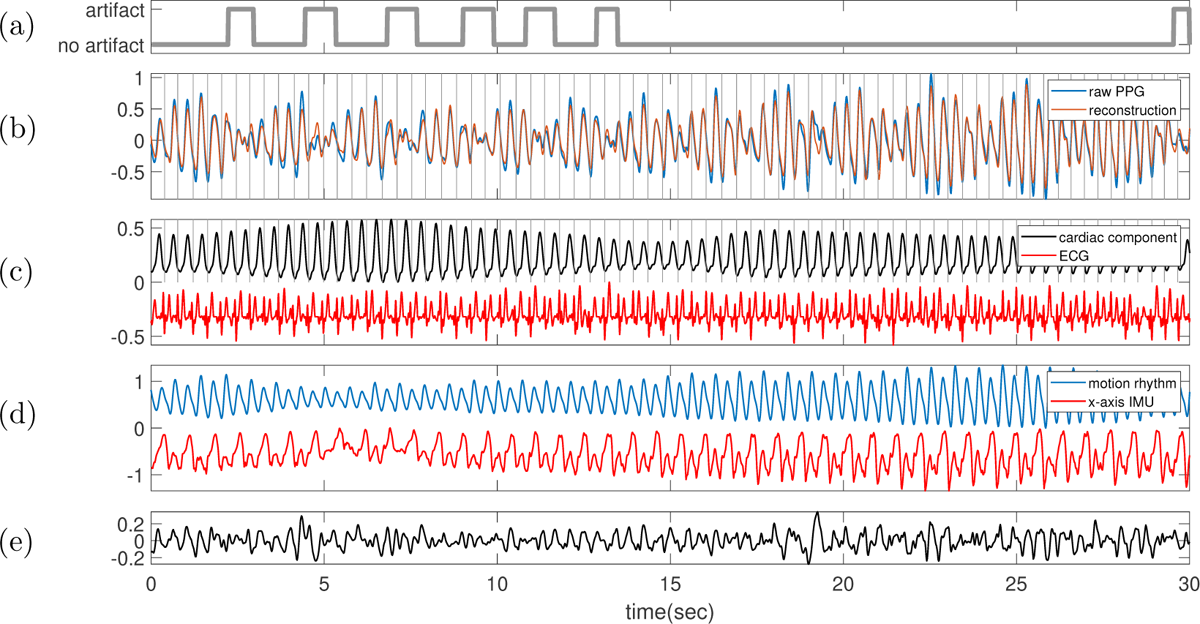
Subject 12 between 210 and 240 seconds in the TROIKA dataset. The subject is running at the speed of 12 kilometer per hour during this segment. **(a)** Label sequence (grey line) and the prediction result by the proposed SQA model (red-dashed line). **(b)** The raw PPG signal is shown in black, and the summation of the decomposed motion rhythm and cardiac component is superimposed in red. The detected R-peaks from the simultaneously recorded ECG are superimposed as vertical grey lines. **(c)** The decomposed cardiac component (black curve), the simultaneously recorded ECG (red curve) and the detected R-peaks (vertical grey lines). **(d)** The decomposed motion rhythm (blue curve) and the x-axis of the accelerometer signal recorded simultaneously (red curve). **(e)** The difference of the PPG signal and the summation of the decomposed motion rhythm and cardiac component. The normalized root mean square error (NRMSE) between the PPG signal and the summation of the decomposed motion rhythm and cardiac component reconstruction is 0.26.

**Figure 44.**
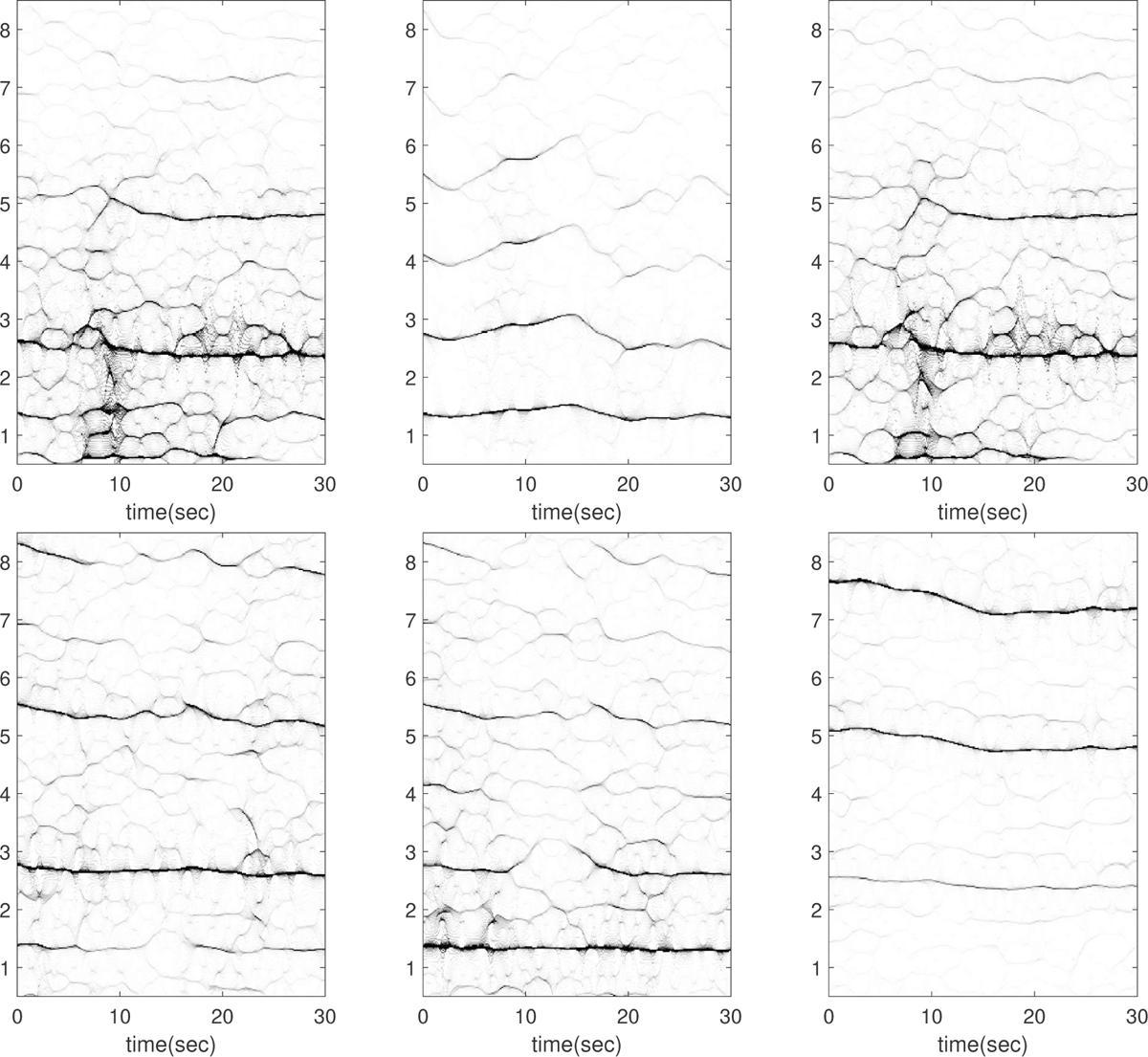
Subject 4 between 150 and 180 seconds in the TROIKA dataset. The subject is running at the speed of 6 kilometer per hour during this segment. The first row, from left to right: the time-frequency representations (TFRs) of the PPG, the decomposed motion rhythm component, and the PPG after removing the decomposed motion rhythm component. The second row, from left to right: the TFRs of the simultaneously recorded accelerometer magnitude signal, the simultaneously recorded x-axis of the accelerometer signal, and the simultaneously recorded ECG signal. Over this segment, the mean heart rate derived from the simultaneously recorded electrocardiogram (resp. raw PPG and extracted cardiac component) is 2.44 Hz (resp. 2.48 Hz, 2.48 Hz). There is an artifact around the 10th second in the PPG, evident in the TFR and confirmed by the x-axis IMU signal.

**Figure 45.**
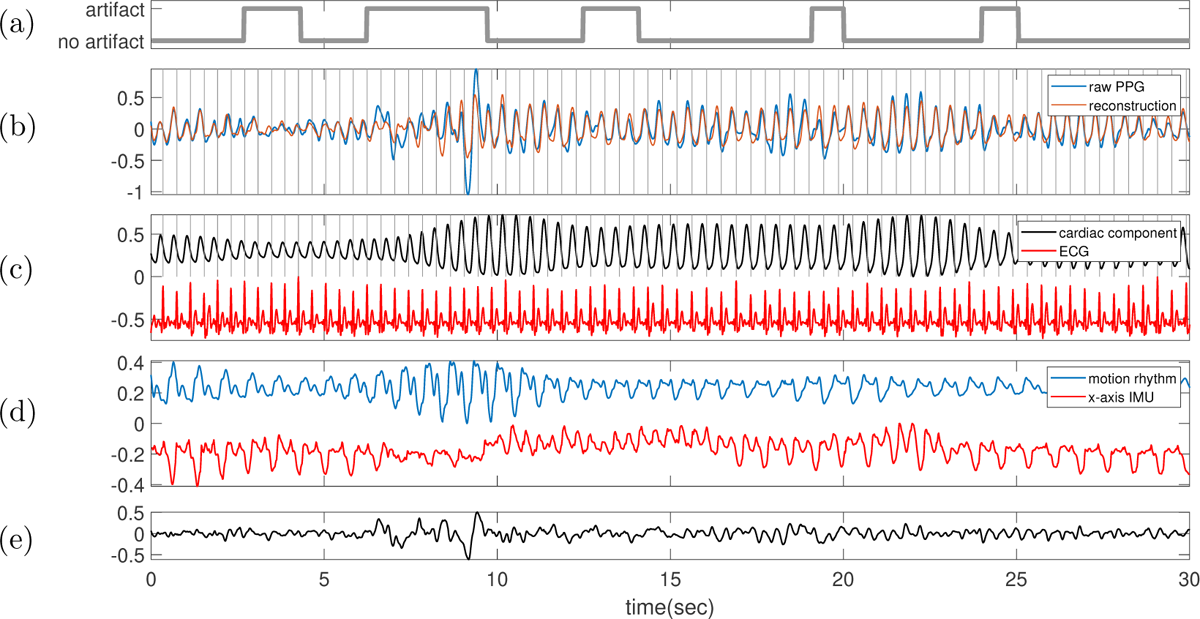
Subject 4 between 150 and 180 seconds in the TROIKA dataset. The subject is running at the speed of 6 kilometer per hour during this segment. **(a)** Label sequence (grey line) and the prediction result by the proposed SQA model (red-dashed line). **(b)** The raw PPG signal is shown in black, and the summation of the decomposed motion rhythm and cardiac component is superimposed in red. The detected R-peaks from the simultaneously recorded ECG are superimposed as vertical grey lines. **(c)** The decomposed cardiac component (black curve), the simultaneously recorded ECG (red curve) and the detected R-peaks (vertical grey lines). **(d)** The decomposed motion rhythm (blue curve) and the x-axis of the accelerometer signal recorded simultaneously (red curve). **(e)** The difference of the PPG signal and the summation of the decomposed motion rhythm and cardiac component. The normalized root mean square error (NRMSE) between the PPG signal and the summation of the decomposed motion rhythm and cardiac component reconstruction is 0.51.

**Figure 46.**
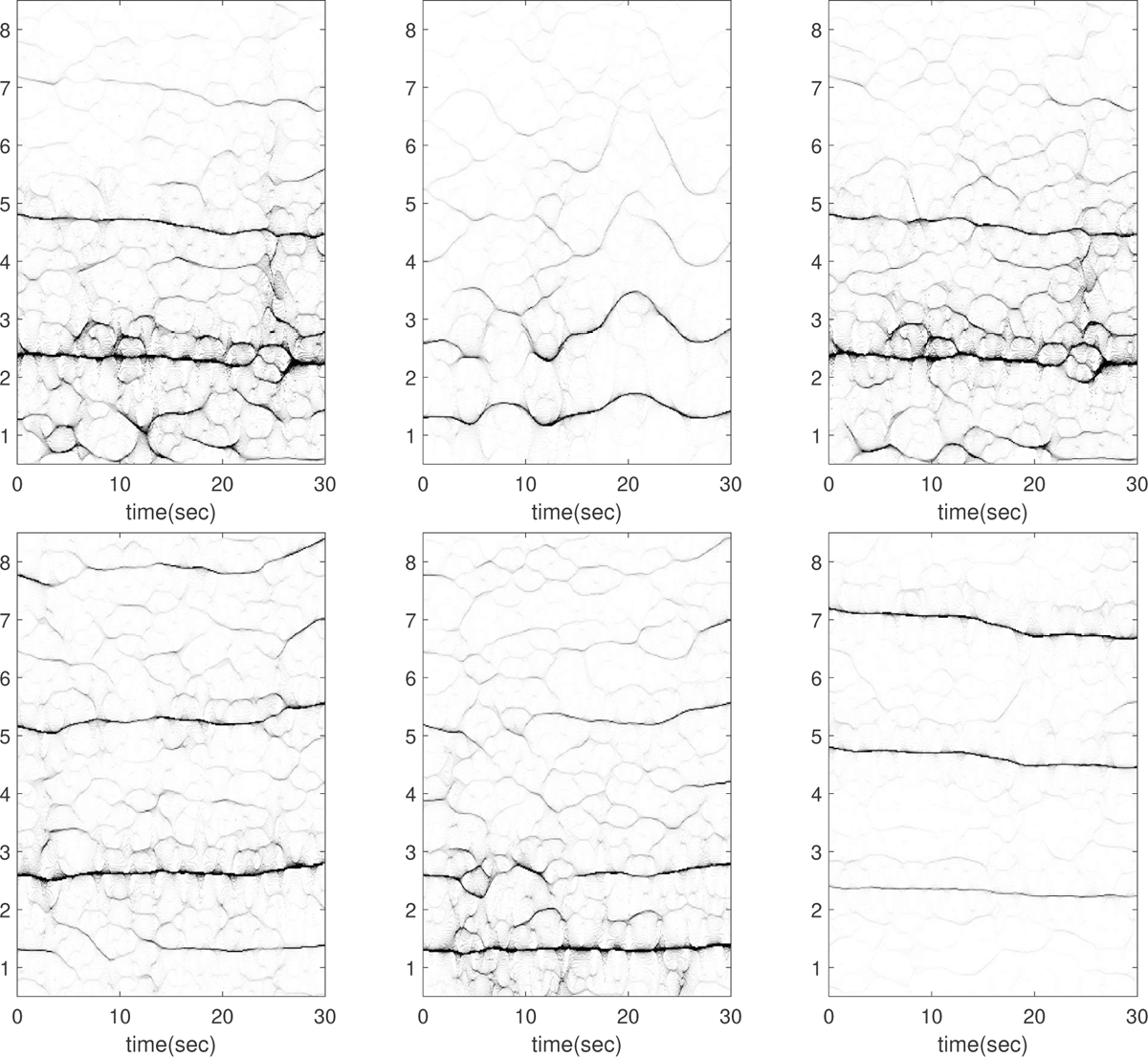
Subject 4 between 180 and 210 seconds in the TROIKA dataset. The subject is running at the speed of 6 kilometer per hour during this segment. The first row, from left to right: the time-frequency representations (TFRs) of the PPG, the decomposed motion rhythm component, and the PPG after removing the decomposed motion rhythm component. The second row, from left to right: the TFRs of the simultaneously recorded accelerometer magnitude signal, the simultaneously recorded x-axis of the accelerometer signal, and the simultaneously recorded ECG signal. Over this segment, the mean heart rate derived from the simultaneously recorded electrocardiogram (resp. raw PPG and extracted cardiac component) is 2.31 Hz (resp. 2.52 Hz, 2.35 Hz). In this case, it is visually obvious that the TFR of the decomposed motion rhythm is different from that of the IMU signal.

**Figure 47.**
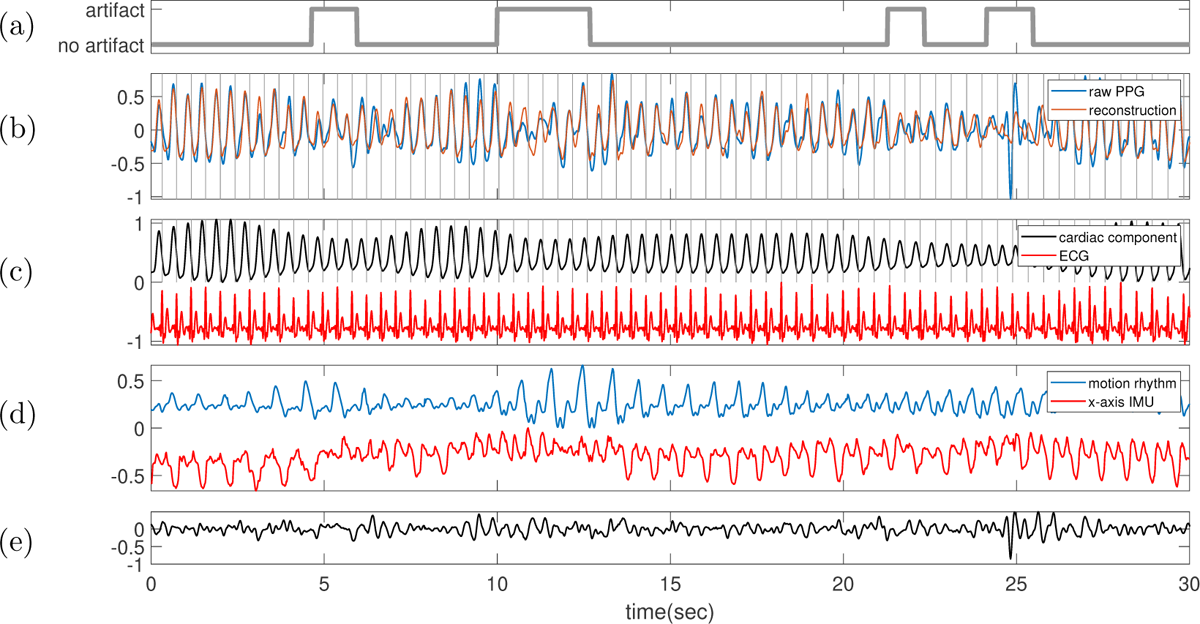
Subject 4 between 180 and 210 seconds in the TROIKA dataset. The subject is running at the speed of 6 kilometer per hour during this segment. **(a)** Label sequence (grey line) and the prediction result by the proposed SQA model (red-dashed line). **(b)** The raw PPG signal is shown in black, and the summation of the decomposed motion rhythm and cardiac component is superimposed in red. The detected R-peaks from the simultaneously recorded ECG are superimposed as vertical grey lines. **(c)** The decomposed cardiac component (black curve), the simultaneously recorded ECG (red curve) and the detected R-peaks (vertical grey lines). **(d)** The decomposed motion rhythm (blue curve) and the x-axis of the accelerometer signal recorded simultaneously (red curve). **(e)** The difference of the PPG signal and the summation of the decomposed motion rhythm and cardiac component. The normalized root mean square error (NRMSE) between the PPG signal and the summation of the decomposed motion rhythm and cardiac component reconstruction is 0.46.

**Figure 48.**
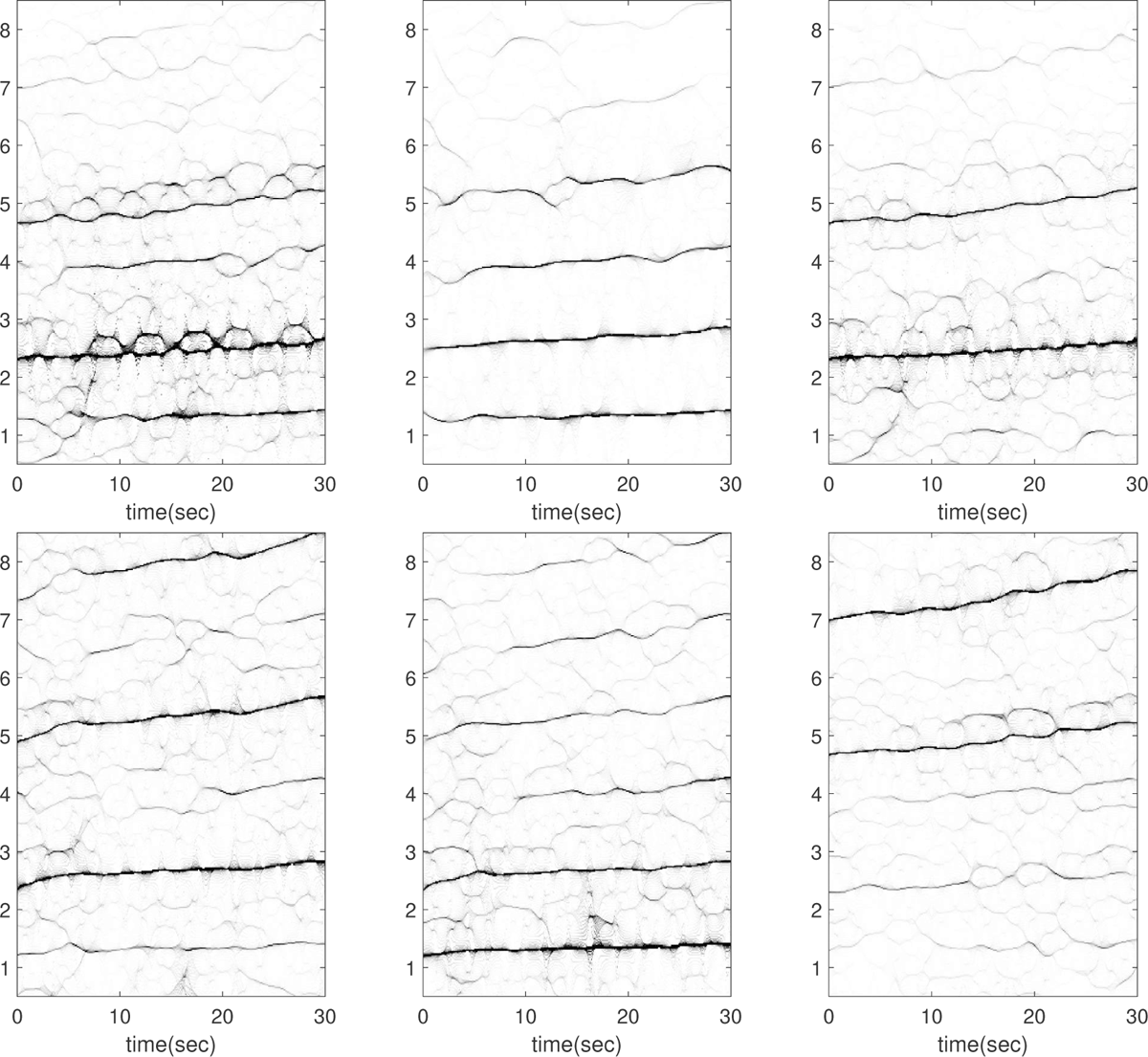
Subject 5 between 210 and 240 seconds in the TROIKA dataset. The subject is running at the speed of 12 kilometer per hour during this segment. The first row, from left to right: the time-frequency representations (TFRs) of the PPG, the decomposed motion rhythm component, and the PPG after removing the decomposed motion rhythm component. The second row, from left to right: the TFRs of the simultaneously recorded accelerometer magnitude signal, the simultaneously recorded x-axis of the accelerometer signal, and the simultaneously recorded ECG signal. Over this segment, the mean heart rate derived from the simultaneously recorded electrocardiogram (resp. raw PPG and extracted cardiac component) is 2.46 Hz (resp. 2.47 Hz, 2.46 Hz).

**Figure 49.**
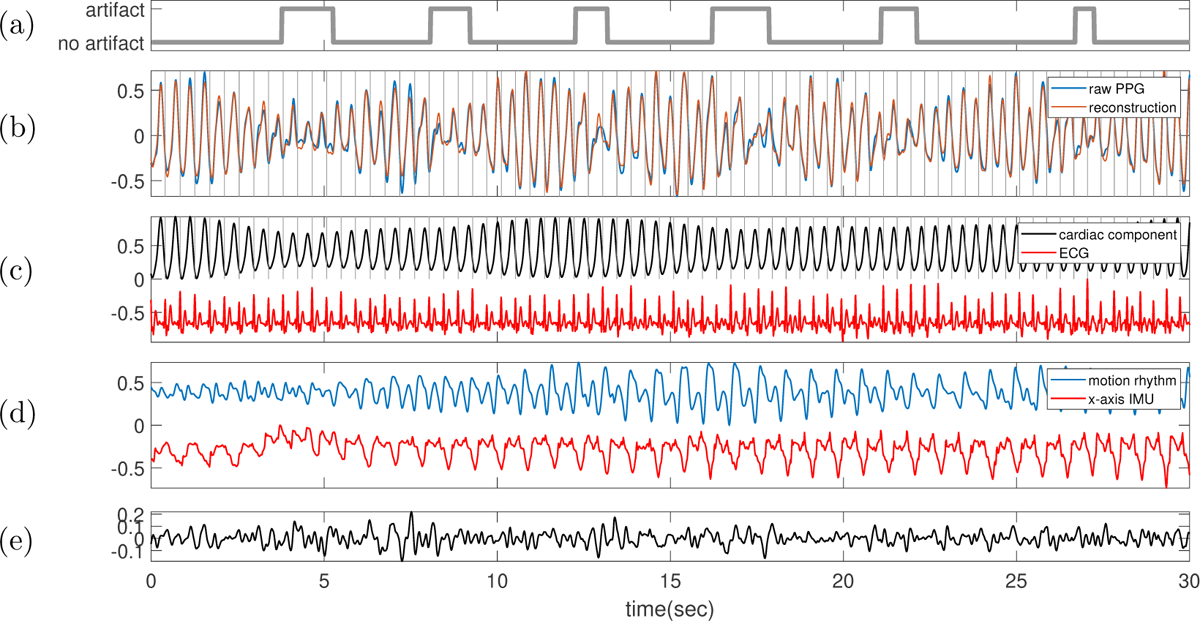
Subject 5 between 210 and 240 seconds in the TROIKA dataset. The subject is running at the speed of 12 kilometer per hour during this segment. **(a)** Label sequence (grey line) and the prediction result by the proposed SQA model (red-dashed line). **(b)** The raw PPG signal is shown in black, and the summation of the decomposed motion rhythm and cardiac component is superimposed in red. The detected R-peaks from the simultaneously recorded ECG are superimposed as vertical grey lines. **(c)** The decomposed cardiac component (black curve), the simultaneously recorded ECG (red curve) and the detected R-peaks (vertical grey lines). **(d)** The decomposed motion rhythm (blue curve) and the x-axis of the accelerometer signal recorded simultaneously (red curve). **(e)** The difference of the PPG signal and the summation of the decomposed motion rhythm and cardiac component. The normalized root mean square error (NRMSE) between the PPG signal and the summation of the decomposed motion rhythm and cardiac component reconstruction is 0.18.

**Figure 50.**
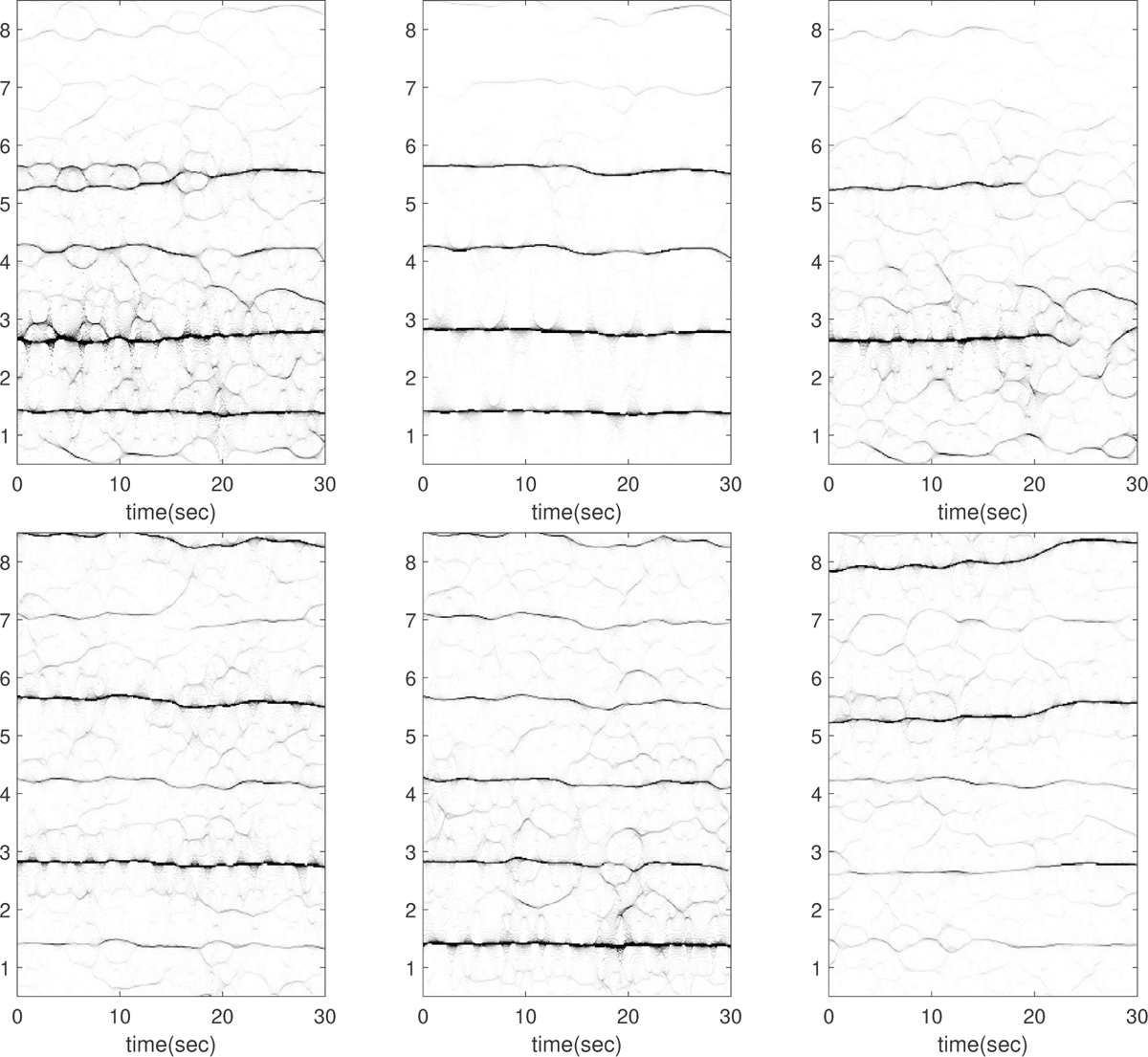
Subject 5 between 240 and 270 seconds in the TROIKA dataset. The subject is running at the speed of 12 kilometer per hour during this segment. The first row, from left to right: the time-frequency representations (TFRs) of the PPG, the decomposed motion rhythm component, and the PPG after removing the decomposed motion rhythm component. The second row, from left to right: the TFRs of the simultaneously recorded accelerometer magnitude signal, the simultaneously recorded x-axis of the accelerometer signal, and the simultaneously recorded ECG signal. Over this segment, the mean heart rate derived from the simultaneously recorded electrocardiogram (resp. raw PPG and extracted cardiac component) is 2.69 Hz (resp. 3.04 Hz, 2.61 Hz). This segment is noteworthy because the instantaneous frequency (IF) of the second harmonic of the IMU signal matches exactly with that of the fundamental component of ECG in the last 10 seconds (see the top row of Figure 51); indicating a strong 2:1 coupling between heart rate and motion rhythm. The current algorithm is limited in decomposing such signals, and the decomposition is not ideal in the last 10 seconds, as illustrated in Figure 51. Moreover, from the TFR of the ECG, there are plausible components oscillating at *∼*1.4 Hz and *∼*4.2 Hz in the ECG, which coincide with the first and third harmonics of the motion rhythm.

**Figure 51.**
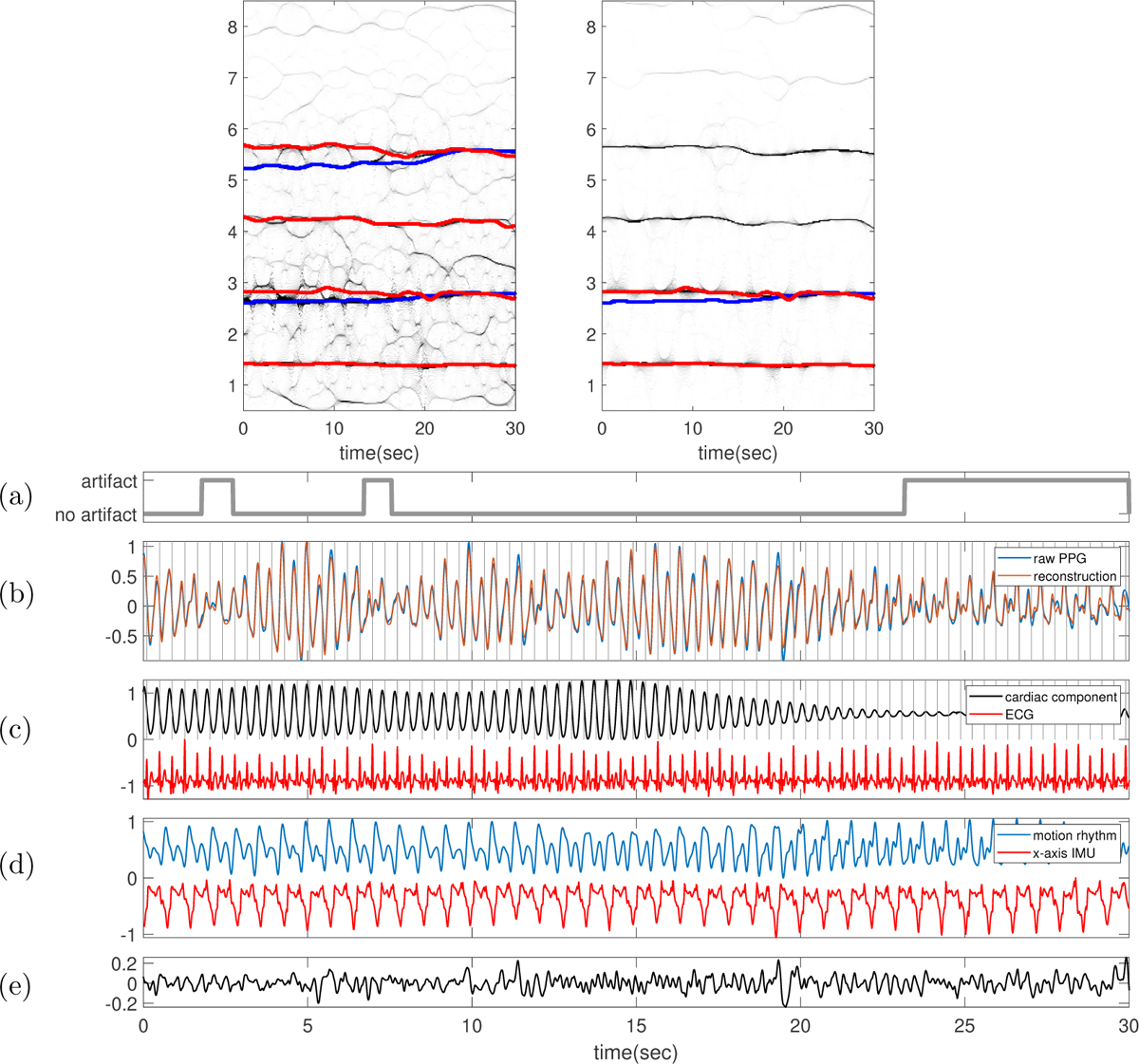
Subject 5 between 240 and 270 seconds in the TROIKA dataset. The subject is running at the speed of 12 kilometer per hour during this segment. In the top row, the TFR of the PPG with the IFs of the first two (four respectively) harmonics of ECG (IMU respectively) superimposed as blue (red respectively) curves are shown on the left-hand side. On the right-hand side, the TFR of the decomposed motion rhythm component with the IF of the first (first two respectively) harmonic of ECG (IMU respectively) superimposed as the blue (red respectively) curve are displayed. **(a)** Label sequence (grey line) and the prediction result by the proposed SQA model (red-dashed line). **(b)** The raw PPG signal is shown in black, and the summation of the decomposed motion rhythm and cardiac component is superimposed in red. The detected R-peaks from the simultaneously recorded ECG are superimposed as vertical grey lines. **(c)** The decomposed cardiac component (black curve), the simultaneously recorded ECG (red curve) and the detected R-peaks (vertical grey lines). **(d)** The decomposed motion rhythm (blue curve) and the x-axis of the accelerometer signal recorded simultaneously (red curve). **(e)** The difference of the PPG signal and the summation of the decomposed motion rhythm and cardiac component. The normalized root mean square error (NRMSE) between the PPG signal and the summation of the decomposed motion rhythm and cardiac component reconstruction is 0.18.

**Figure 52.**
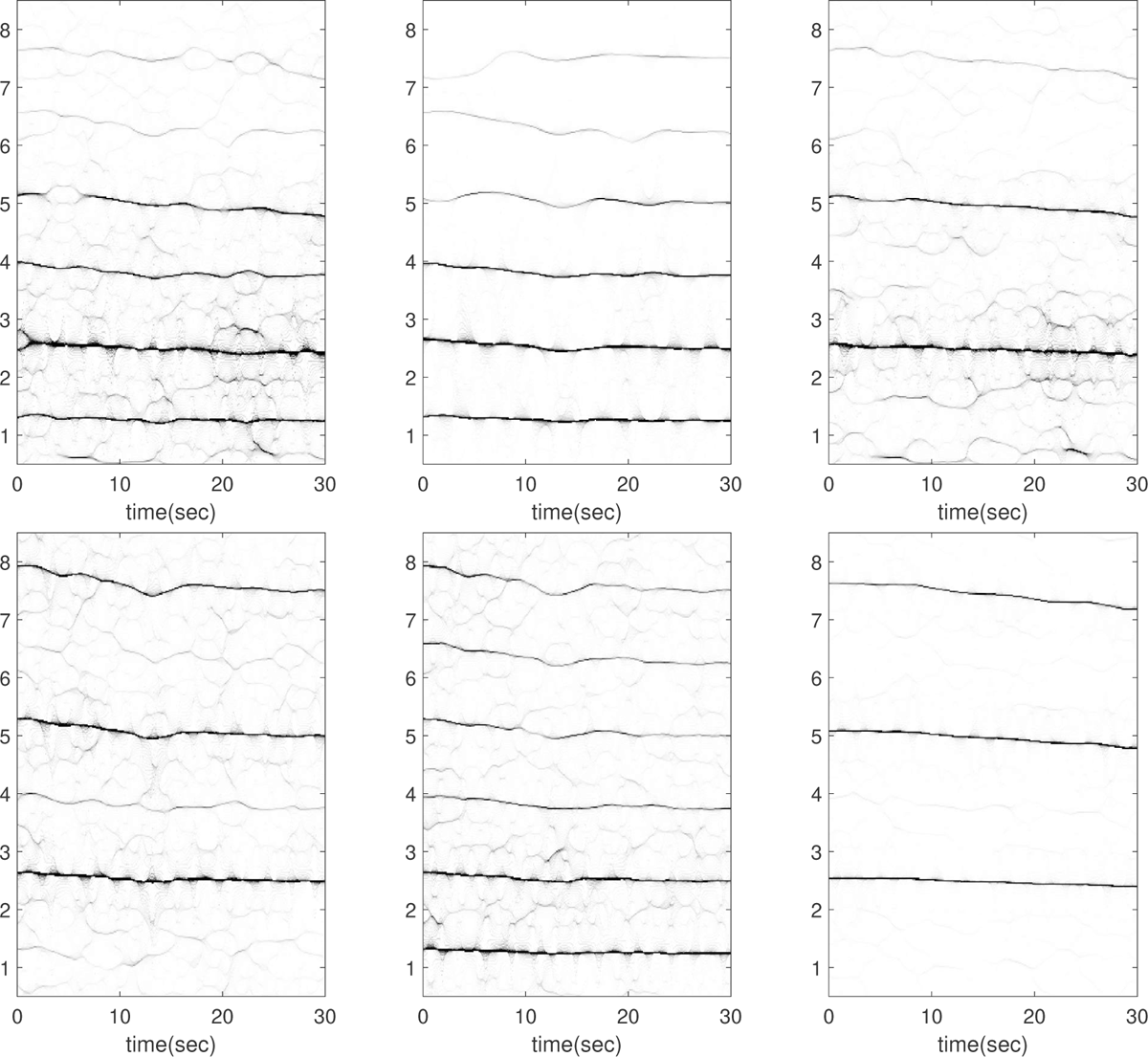
Subject 6 between 150 and 180 seconds in the TROIKA dataset. The subject is running at the speed of 6 kilometer per hour during this segment. The first row, from left to right: the time-frequency representations (TFRs) of the PPG, the decomposed motion rhythm component, and the PPG after removing the decomposed motion rhythm component. The second row, from left to right: the TFRs of the simultaneously recorded accelerometer magnitude signal, the simultaneously recorded x-axis of the accelerometer signal, and the simultaneously recorded ECG signal. Over this segment, the mean heart rate derived from the simultaneously recorded electrocardiogram (resp. raw PPG and extracted cardiac component) is 2.48 Hz (resp. 2.58 Hz, 2.48 Hz).

**Figure 53.**
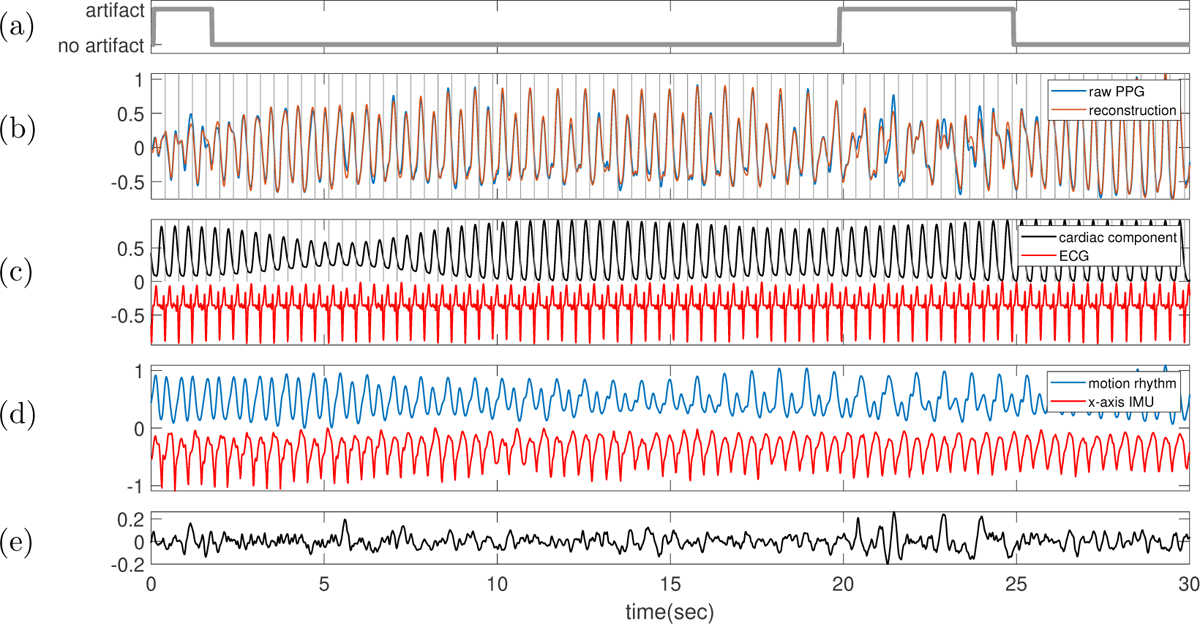
Subject 6 between 150 and 180 seconds in the TROIKA dataset. The subject is running at the speed of 6 kilometer per hour during this segment. **(a)** Label sequence (grey line) and the prediction result by the proposed SQA model (red-dashed line). **(b)** The raw PPG signal is shown in black, and the summation of the decomposed motion rhythm and cardiac component is superimposed in red. The detected R-peaks from the simultaneously recorded ECG are superimposed as vertical grey lines. **(c)** The decomposed cardiac component (black curve), the simultaneously recorded ECG (red curve) and the detected R-peaks (vertical grey lines). **(d)** The decomposed motion rhythm (blue curve) and the x-axis of the accelerometer signal recorded simultaneously (red curve). **(e)** The difference of the PPG signal and the summation of the decomposed motion rhythm and cardiac component. The normalized root mean square error (NRMSE) between the PPG signal and the summation of the decomposed motion rhythm and cardiac component reconstruction is 0.15. Department of Applied Mathematics, National Yang Ming Chiao Tung University, Hsinchu, Taiwan *E-mail address*: Data Science Degree Program, National Taiwan University and Academia Sinica, Taipei, Taiwan *E-mail address*: Department of Mathematics, National Chen-Kung University, Tainan, Taiwan and National Center for Theoretical Sciences, National Taiwan University, Taipei, Taiwan *E-mail address*: Department of Applied Mathematics, National Yang Ming Chiao Tung University, Hsinchu, Taiwan *E-mail address*: Courant Institute of Mathematical Sciences, New York University, New York, NY, 10012 USA *E-mail address*

**Figure 54.**
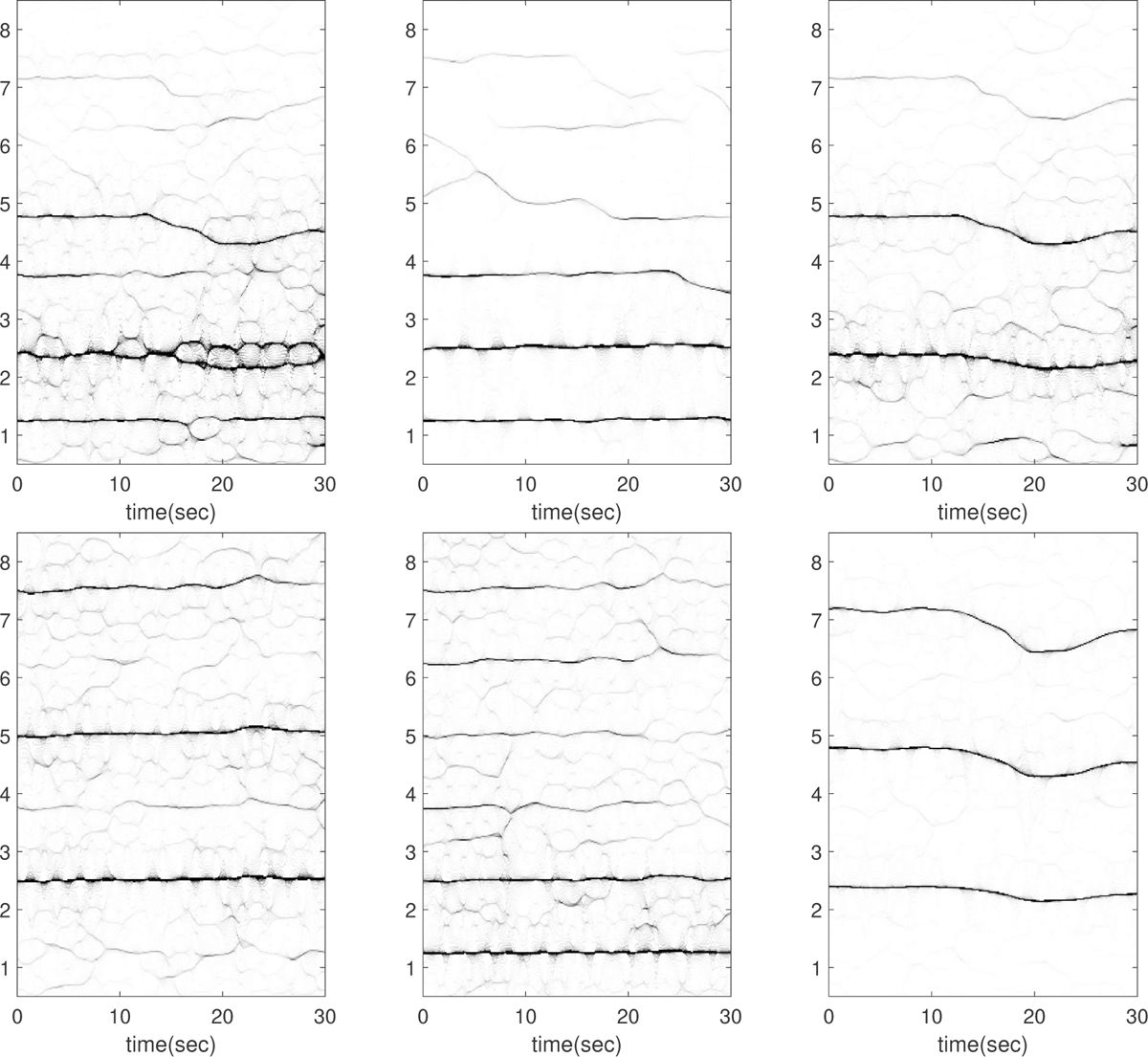
Subject 6 between 180 and 210 seconds in the TROIKA dataset. The subject is running at the speed of 6 kilometer per hour during this segment. The first row, from left to right: the time-frequency representations (TFRs) of the PPG, the decomposed motion rhythm component, and the PPG after removing the decomposed motion rhythm component. The second row, from left to right: the TFRs of the simultaneously recorded accelerometer magnitude signal, the simultaneously recorded x-axis of the accelerometer signal, and the simultaneously recorded ECG signal. Over this segment, the mean heart rate derived from the simultaneously recorded electrocardiogram (resp. raw PPG and extracted cardiac component) is 2.30 Hz (resp. 2.63 Hz, 2.30 Hz). In this case, the instantaneous heart rate changes fast at around 18th second.

**Figure 55.**
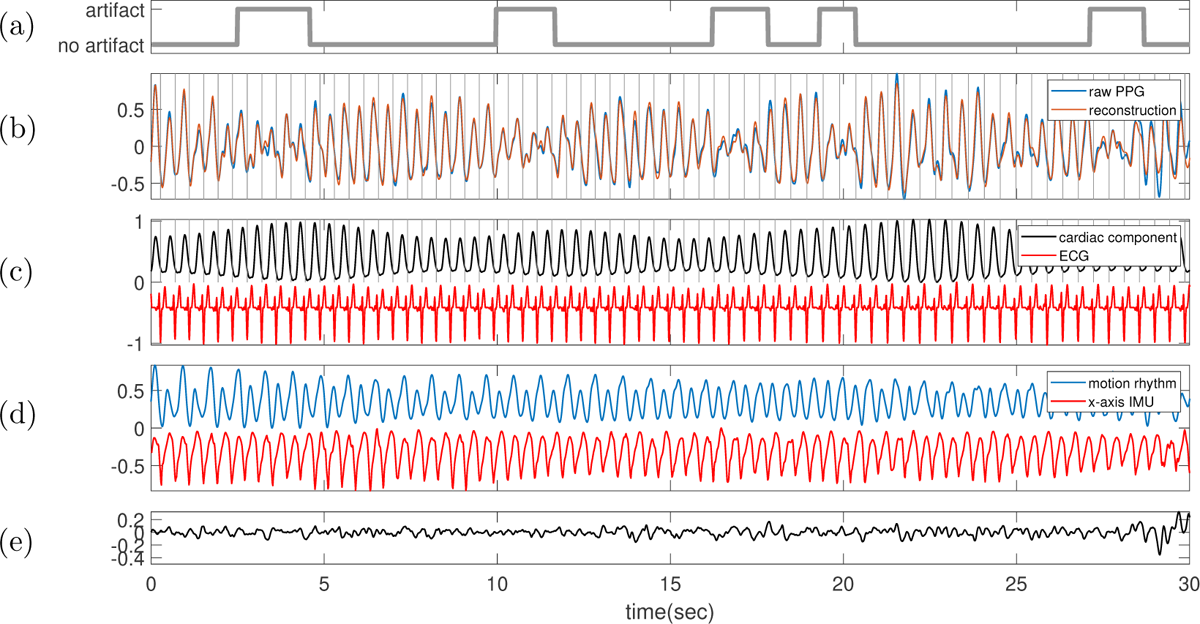
Subject 6 between 180 and 210 seconds in the TROIKA dataset. The subject is running at the speed of 6 kilometer per hour during this segment. **(a)** Label sequence (grey line) and the prediction result by the proposed SQA model (red-dashed line). **(b)** The raw PPG signal is shown in black, and the summation of the decomposed motion rhythm and cardiac component is superimposed in red. The detected R-peaks from the simultaneously recorded ECG are superimposed as vertical grey lines. **(c)** The decomposed cardiac component (black curve), the simultaneously recorded ECG (red curve) and the detected R-peaks (vertical grey lines). **(d)** The decomposed motion rhythm (blue curve) and the x-axis of the accelerometer signal recorded simultaneously (red curve). **(e)** The difference of the PPG signal and the summation of the decomposed motion rhythm and cardiac component. The normalized root mean square error (NRMSE) between the PPG signal and the summation of the decomposed motion rhythm and cardiac component reconstruction is 0.19.

**Figure 56.**
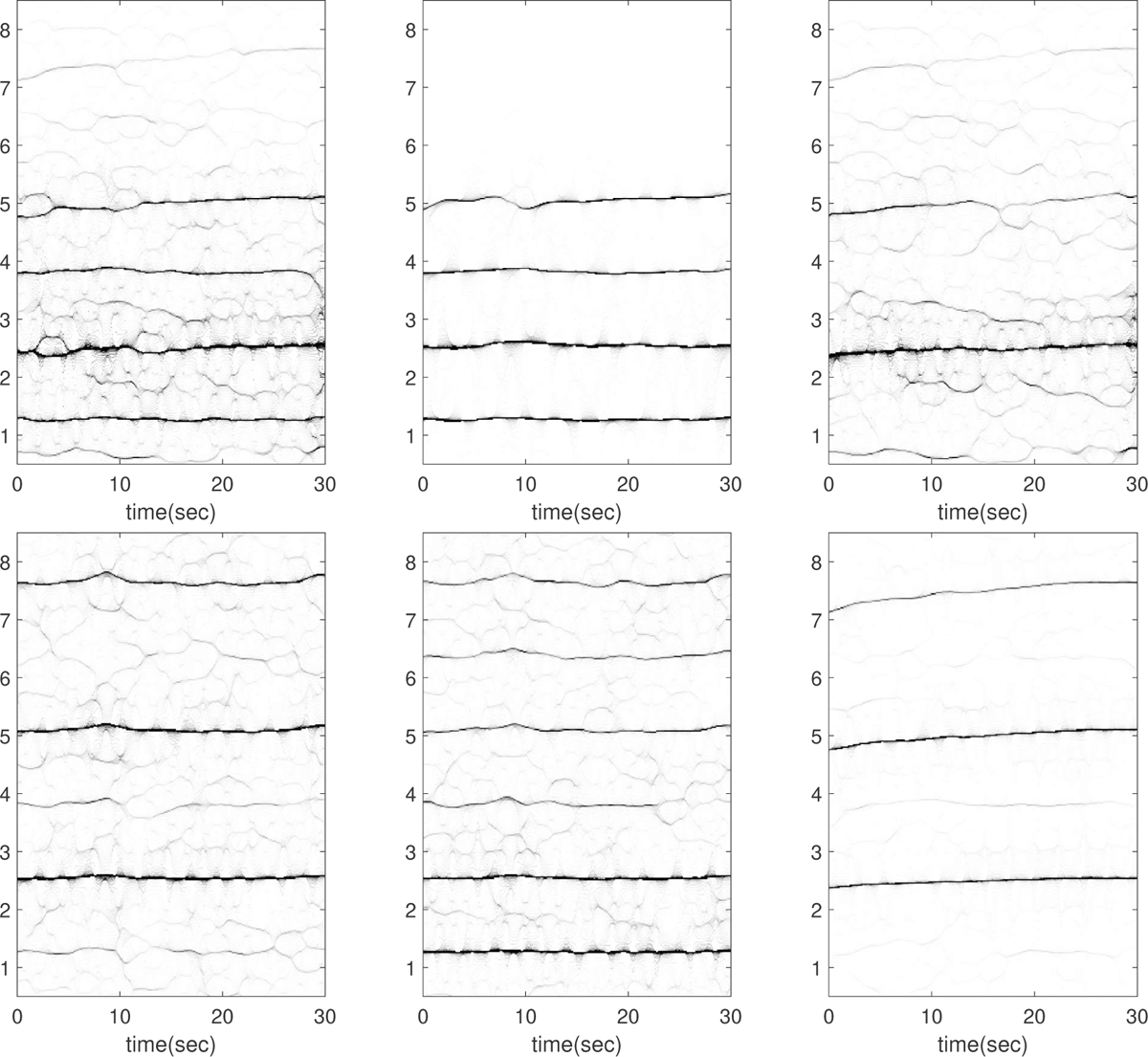
Subject 6 between 240 and 270 seconds in the TROIKA dataset. The subject is running at the speed of 12 kilometer per hour during this segment. The first row, from left to right: the time-frequency representations (TFRs) of the PPG, the decomposed motion rhythm component, and the PPG after removing the decomposed motion rhythm component. The second row, from left to right: the TFRs of the simultaneously recorded accelerometer magnitude signal, the simultaneously recorded x-axis of the accelerometer signal, and the simultaneously recorded ECG signal. Over this segment, the mean heart rate derived from the simultaneously recorded electrocardiogram (resp. raw PPG and extracted cardiac component) is 2.50 Hz (resp. 2.96 Hz, 2.49 Hz). Note that the fundamental component of the accelerometer magnitude signal (*∼* 1.2 Hz) is relatively weak in this segment.

**Figure 57.**
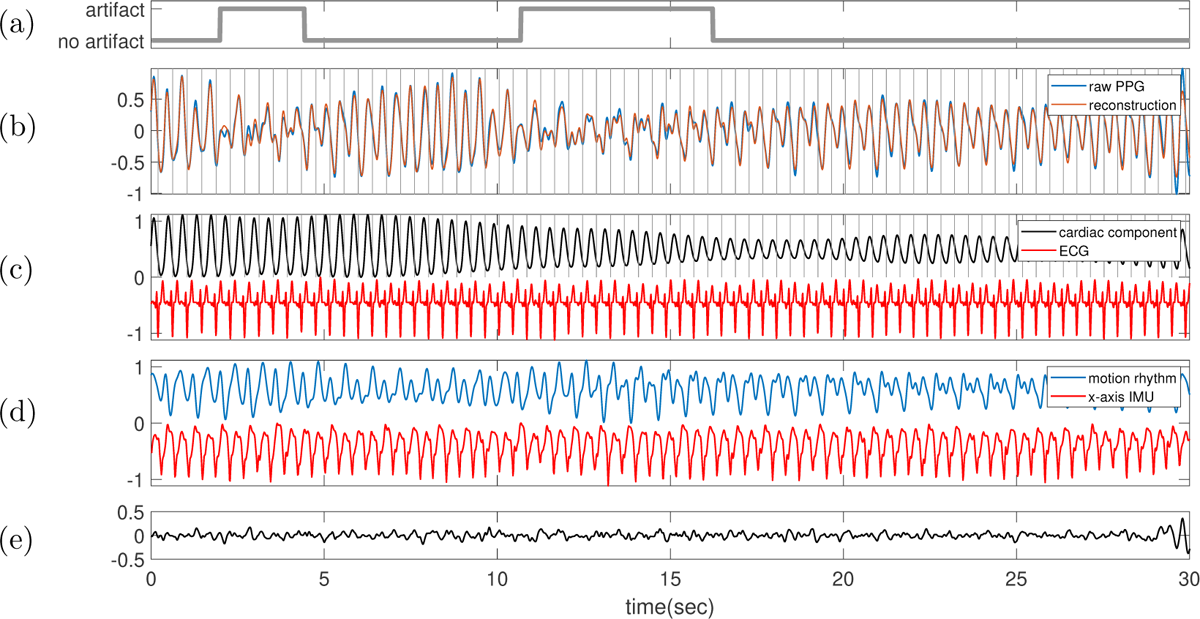
Subject 6 between 240 and 270 seconds in the TROIKA dataset. The subject is running at the speed of 12 kilometer per hour during this segment. **(a)** Label sequence (grey line) and the prediction result by the proposed SQA model (red-dashed line). **(b)** The raw PPG signal is shown in black, and the summation of the decomposed motion rhythm and cardiac component is superimposed in red. The detected R-peaks from the simultaneously recorded ECG are superimposed as vertical grey lines. **(c)** The decomposed cardiac component (black curve), the simultaneously recorded ECG (red curve) and the detected R-peaks (vertical grey lines). **(d)** The decomposed motion rhythm (blue curve) and the x-axis of the accelerometer signal recorded simultaneously (red curve). **(e)** The difference of the PPG signal and the summation of the decomposed motion rhythm and cardiac component. The normalized root mean square error (NRMSE) between the PPG signal and the summation of the decomposed motion rhythm and cardiac component reconstruction is 0.19.

**Figure 58.**
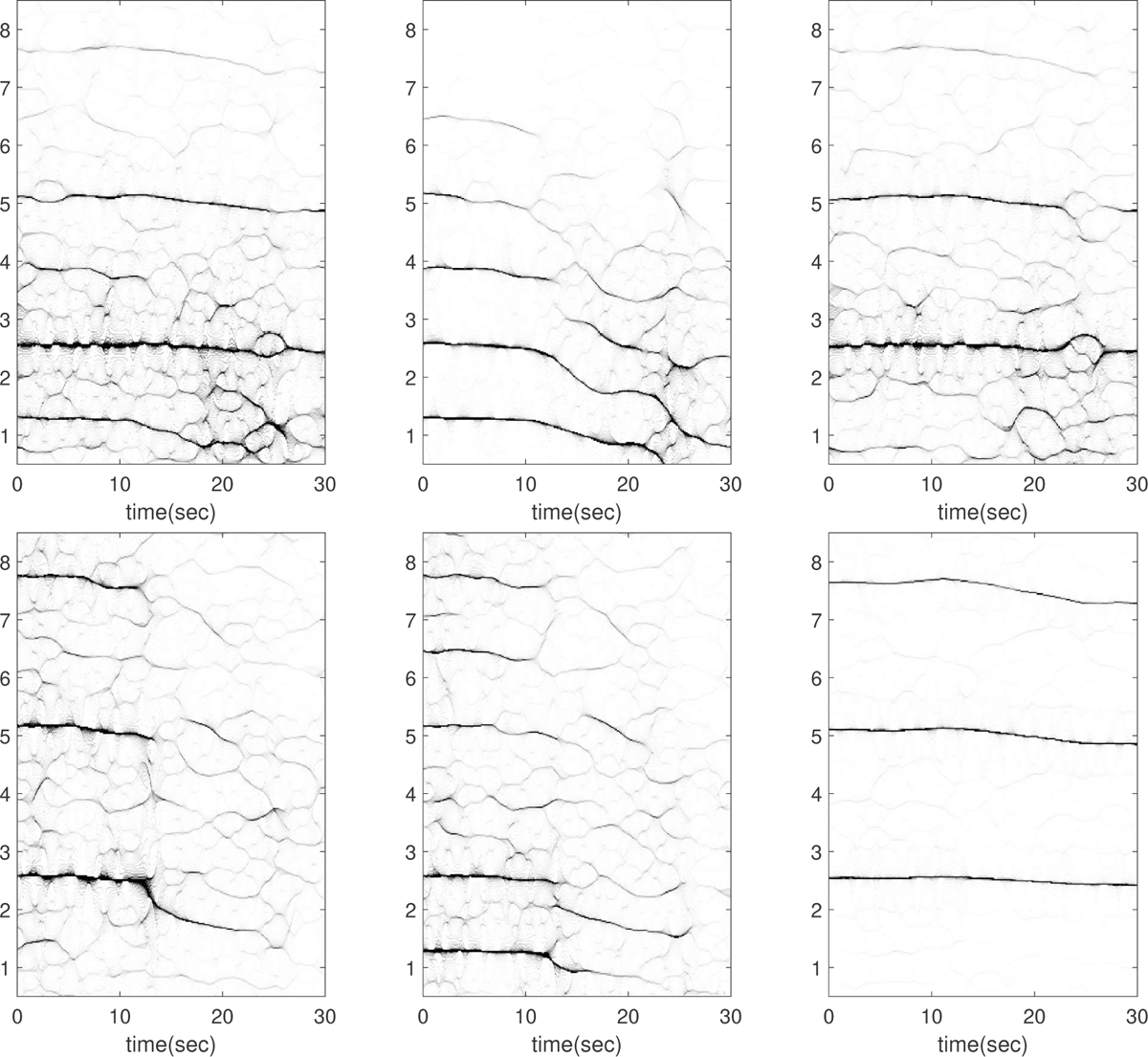
Subject 6 between 270 and 300 seconds in the TROIKA dataset. The subject is resting during this segment. The first row, from left to right: the time-frequency representations (TFRs) of the PPG, the decomposed motion rhythm component, and the PPG after removing the decomposed motion rhythm component. The second row, from left to right: the TFRs of the simultaneously recorded accelerometer magnitude signal, the simultaneously recorded x-axis of the accelerometer signal, and the simultaneously recorded ECG signal. Over this segment, the mean heart rate derived from the simultaneously recorded electro-cardiogram (resp. raw PPG and extracted cardiac component) is 2.51 Hz (resp. 2.54 Hz, 2.55 Hz). While the experts’ label indicates that the subject is resting, an obvious oscillation is observed in the accelerometer signals, which is shown in Figure 59. In this case, the instantaneous frequency of the accelerometer signal (also evident in the PPG signal) suddenly drops from *∼* 1.2 Hz, and the amplitude modulation (strength) also decreases. While detailed information about the recording environment is lacking, it is hypothesized that the subject might have been walking initially, possibly transitioning to a resting mode during the recording of this segment.

**Figure 59.**
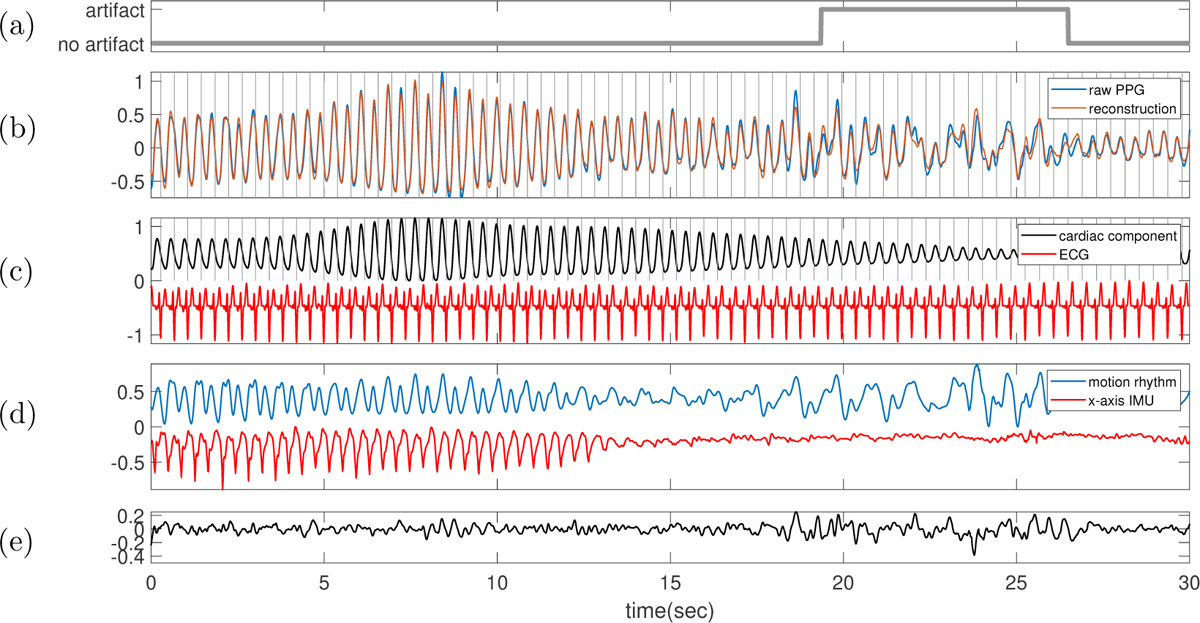
Subject 6 between 270 and 300 seconds in the TROIKA dataset. The subject is resting during this segment. **(a)** Label sequence (grey line) and the prediction result by the proposed SQA model (red-dashed line). **(b)** The raw PPG signal is shown in black, and the summation of the decomposed motion rhythm and cardiac component is superimposed in red. The detected R-peaks from the simultaneously recorded ECG are superimposed as vertical grey lines. **(c)** The decomposed cardiac component (black curve), the simultaneously recorded ECG (red curve) and the detected R-peaks (vertical grey lines). **(d)** The decomposed motion rhythm (blue curve) and the x-axis of the accelerometer signal recorded simultaneously (red curve). **(e)** The difference of the PPG signal and the summation of the decomposed motion rhythm and cardiac component. The normalized root mean square error (NRMSE) between the PPG signal and the summation of the decomposed motion rhythm and cardiac component reconstruction is 0.21.

**Figure 60.**
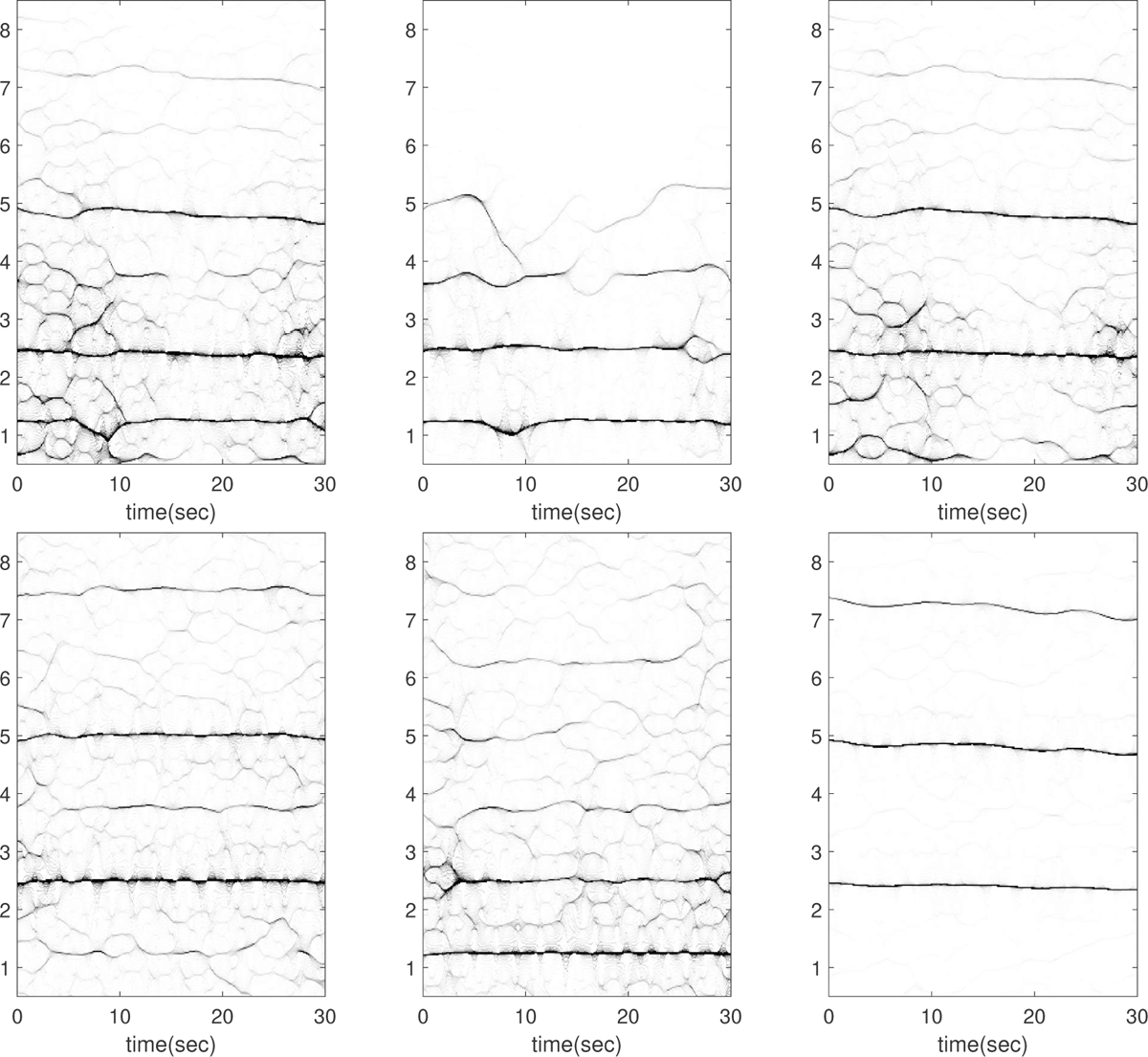
Subject 7 between 180 and 210 seconds in the TROIKA dataset. The subject is running at the speed of 6 kilometer per hour during this segment. The first row, from left to right: the time-frequency representations (TFRs) of the PPG, the decomposed motion rhythm component, and the PPG after removing the decomposed motion rhythm component. The second row, from left to right: the TFRs of the simultaneously recorded accelerometer magnitude signal, the simultaneously recorded x-axis of the accelerometer signal, and the simultaneously recorded ECG signal. Over this segment, the mean heart rate derived from the simultaneously recorded electrocardiogram (resp. raw PPG and extracted cardiac component) is 2.40 Hz (resp. 2.63 Hz, 2.40 Hz).

**Figure 61.**
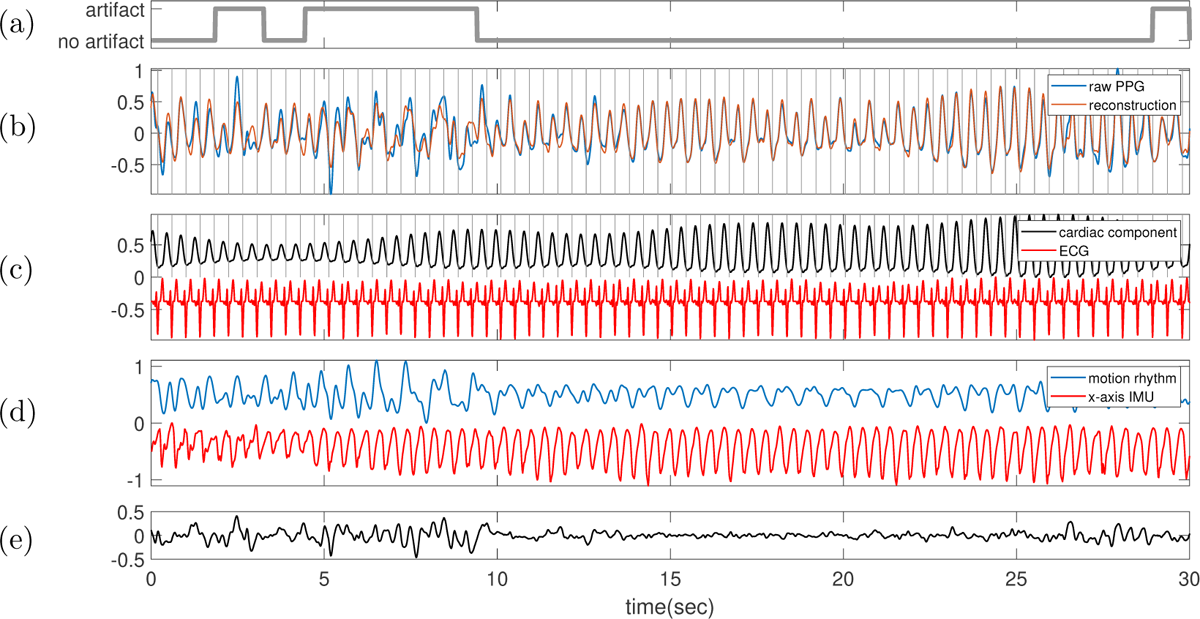
Subject 7 between 180 and 210 seconds in the TROIKA dataset. The subject is running at the speed of 6 kilometer per hour during this segment. **(a)** Label sequence (grey line) and the prediction result by the proposed SQA model (red-dashed line). **(b)** The raw PPG signal is shown in black, and the summation of the decomposed motion rhythm and cardiac component is superimposed in red. The detected R-peaks from the simultaneously recorded ECG are superimposed as vertical grey lines. **(c)** The decomposed cardiac component (black curve), the simultaneously recorded ECG (red curve) and the detected R-peaks (vertical grey lines). **(d)** The decomposed motion rhythm (blue curve) and the x-axis of the accelerometer signal recorded simultaneously (red curve). **(e)** The difference of the PPG signal and the summation of the decomposed motion rhythm and cardiac component. The normalized root mean square error (NRMSE) between the PPG signal and the summation of the decomposed motion rhythm and cardiac component reconstruction is 0.32.

**Figure 62.**
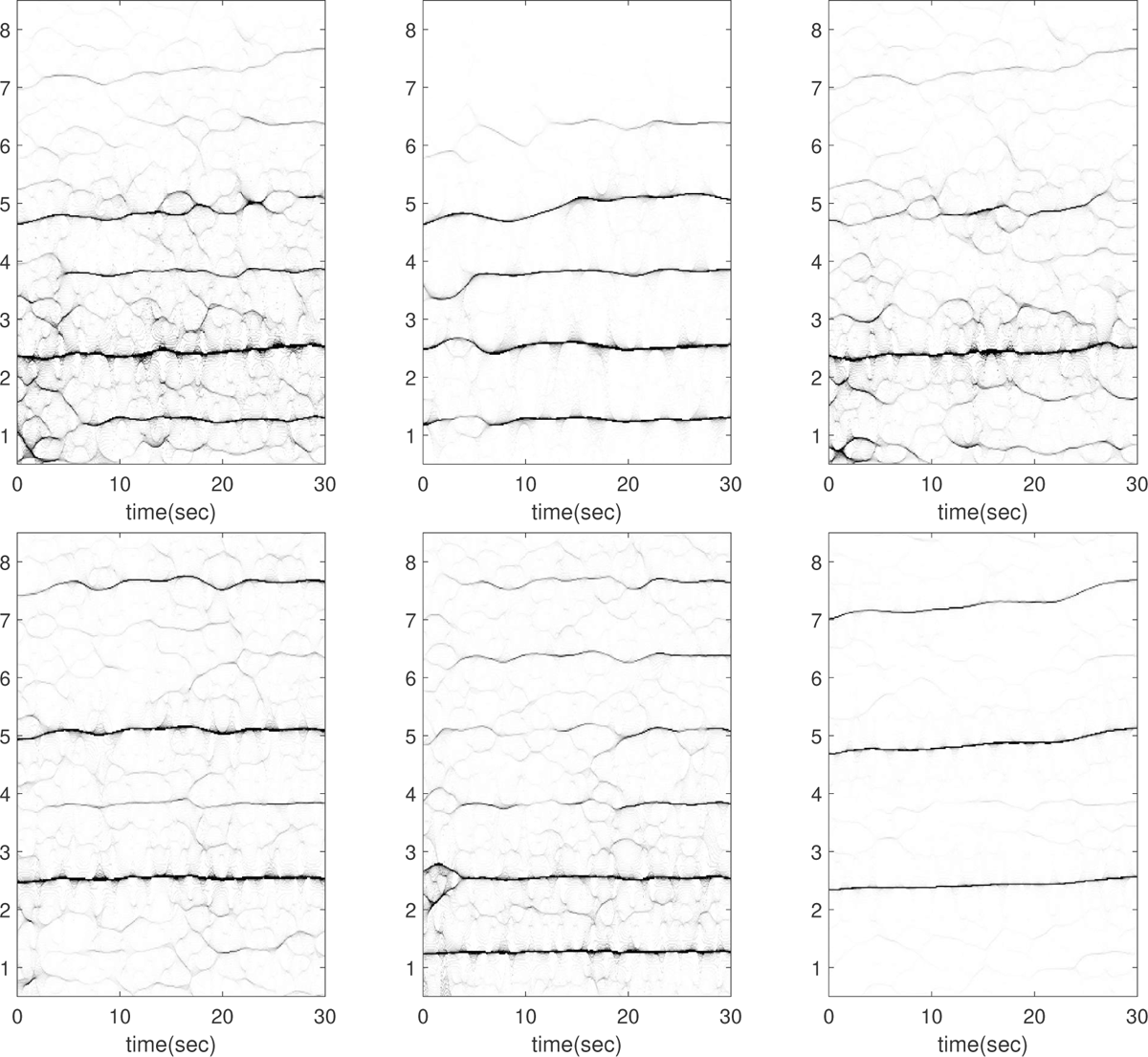
Subject 7 between 210 and 240 seconds in the TROIKA dataset. The subject is running at the speed of 12 kilometer per hour during this segment. The first row, from left to right: the time-frequency representations (TFRs) of the PPG, the decomposed motion rhythm component, and the PPG after removing the decomposed motion rhythm component. The second row, from left to right: the TFRs of the simultaneously recorded accelerometer magnitude signal, the simultaneously recorded x-axis of the accelerometer signal, and the simultaneously recorded ECG signal. Over this segment, the mean heart rate derived from the simultaneously recorded electrocardiogram (resp. raw PPG and extracted cardiac component) is 2.43 Hz (resp. 2.50 Hz, 2.43 Hz).

**Figure 63.**
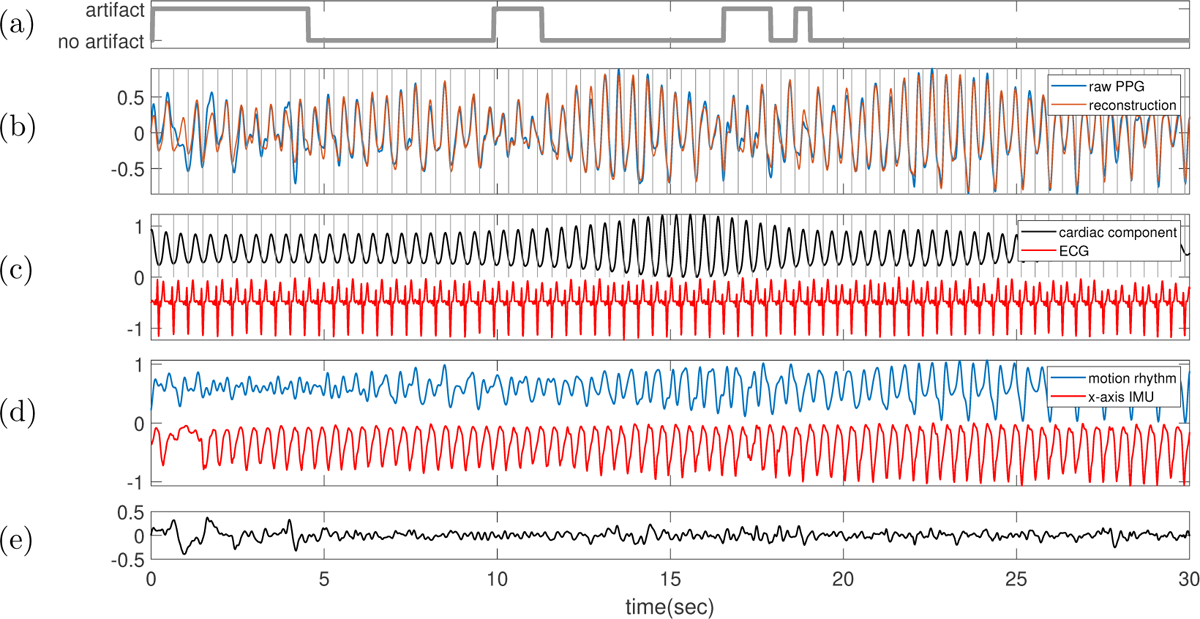
Subject 7 between 210 and 240 seconds in the TROIKA dataset. The subject is running at the speed of 12 kilometer per hour during this segment. **(a)** Label sequence (grey line) and the prediction result by the proposed SQA model (red-dashed line). **(b)** The raw PPG signal is shown in black, and the summation of the decomposed motion rhythm and cardiac component is superimposed in red. The detected R-peaks from the simultaneously recorded ECG are superimposed as vertical grey lines. **(c)** The decomposed cardiac component (black curve), the simultaneously recorded ECG (red curve) and the detected R-peaks (vertical grey lines). **(d)** The decomposed motion rhythm (blue curve) and the x-axis of the accelerometer signal recorded simultaneously (red curve). **(e)** The difference of the PPG signal and the summation of the decomposed motion rhythm and cardiac component. The normalized root mean square error (NRMSE) between the PPG signal and the summation of the decomposed motion rhythm and cardiac component reconstruction is 0.26.

**Figure 64.**
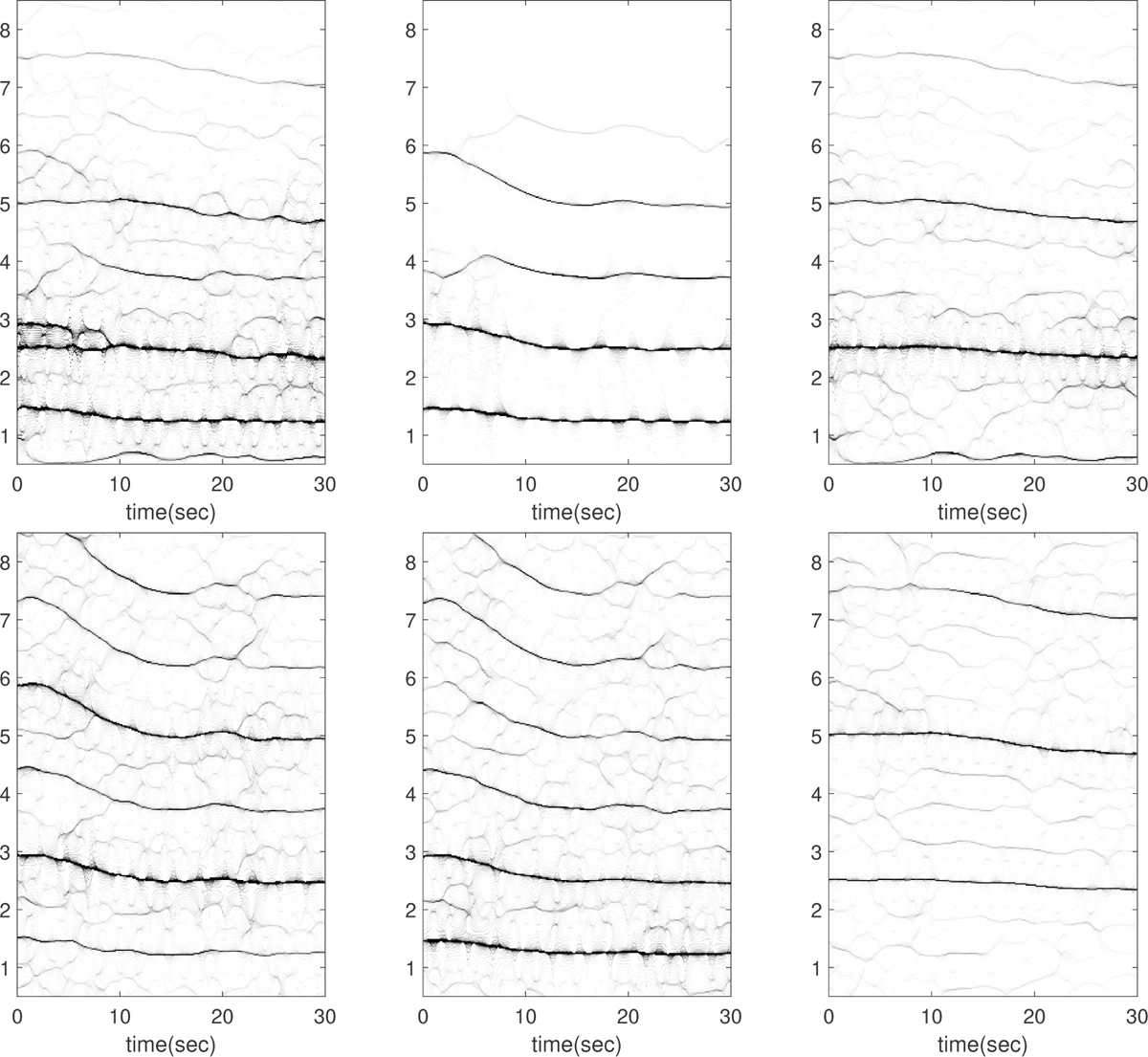
Subject 8 between 150 and 180 seconds in the TROIKA dataset. The subject is running at the speed of 6 kilometer per hour during this segment. The first row, from left to right: the time-frequency representations (TFRs) of the PPG, the decomposed motion rhythm component, and the PPG after removing the decomposed motion rhythm component. The second row, from left to right: the TFRs of the simultaneously recorded accelerometer magnitude signal, the simultaneously recorded x-axis of the accelerometer signal, and the simultaneously recorded ECG signal. Over this segment, the mean heart rate derived from the simultaneously recorded electrocardiogram (resp. raw PPG and extracted cardiac component) is 2.45 Hz (resp. 2.46 Hz, 2.45 Hz). In this case, the instantaneous frequency of the motion rhythm changes decreases from 1.5Hz in the beginning to around 1.25Hz in the end.

**Figure 65.**
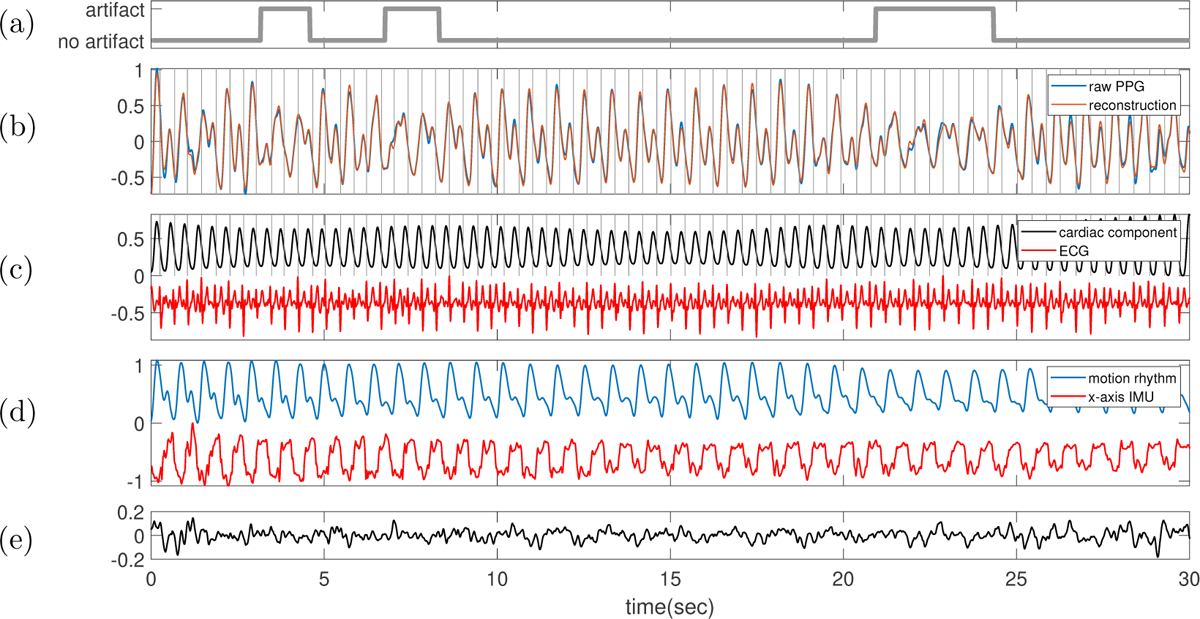
Subject 8 between 150 and 180 seconds in the TROIKA dataset. The subject is running at the speed of 6 kilometer per hour during this segment. **(a)** Label sequence (grey line) and the prediction result by the proposed SQA model (red-dashed line). **(b)** The raw PPG signal is shown in black, and the summation of the decomposed motion rhythm and cardiac component is superimposed in red. The detected R-peaks from the simultaneously recorded ECG are superimposed as vertical grey lines. **(c)** The decomposed cardiac component (black curve), the simultaneously recorded ECG (red curve) and the detected R-peaks (vertical grey lines). **(d)** The decomposed motion rhythm (blue curve) and the x-axis of the accelerometer signal recorded simultaneously (red curve). **(e)** The difference of the PPG signal and the summation of the decomposed motion rhythm and cardiac component. The normalized root mean square error (NRMSE) between the PPG signal and the summation of the decomposed motion rhythm and cardiac component reconstruction is 0.13. This example is noteworthy since the wave-shape function of the decomposed motion rhythm is “similar” to the usually seen PPG signal in the resting status.

**Figure 66.**
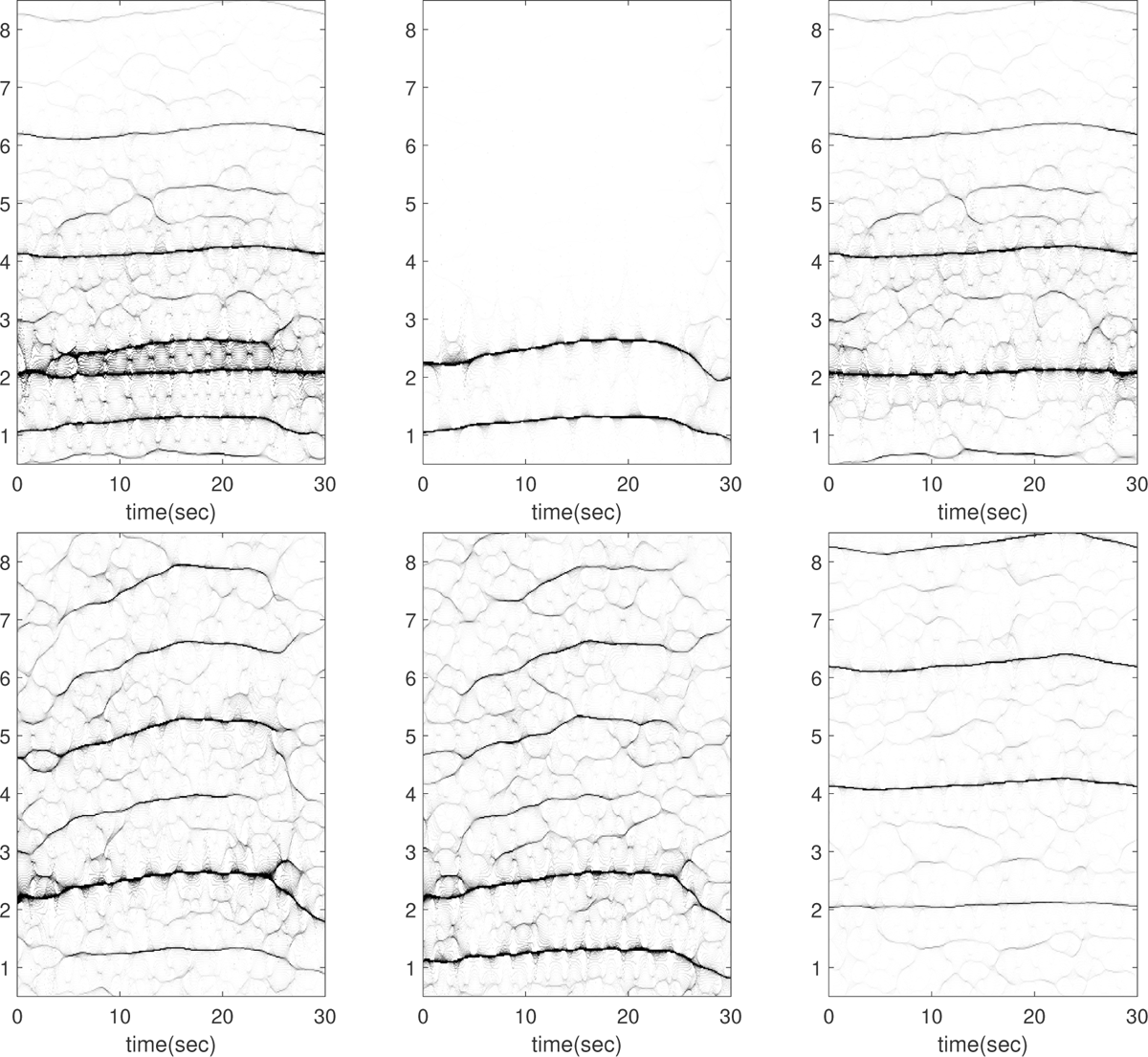
Subject 8 between 270 and 300 seconds in the TROIKA dataset. The subject is resting during this segment. The first row, from left to right: the time-frequency representations (TFRs) of the PPG, the decomposed motion rhythm component, and the PPG after removing the decomposed motion rhythm component. The second row, from left to right: the TFRs of the simultaneously recorded accelerometer magnitude signal, the simultaneously recorded x-axis of the accelerometer signal, and the simultaneously recorded ECG signal. Over this segment, the mean heart rate derived from the simultaneously recorded electrocardiogram (resp. raw PPG and extracted cardiac component) is 2.08 Hz (resp. 2.62 Hz, 2.08 Hz). In this case, the instantaneous frequency of the motion rhythm increases from about 1.1 Hz initially to about 1.25 Hz around the 20th second, and then decreases to around 0.9 Hz towards the end.

**Figure 67.**
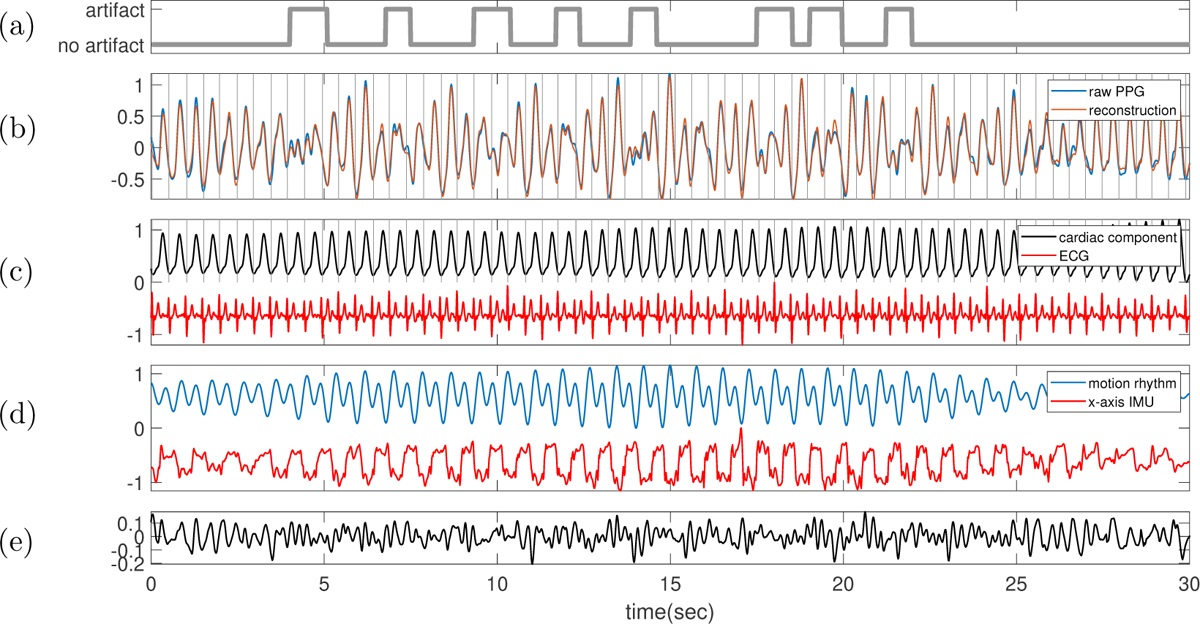
Subject 8 between 270 and 300 seconds in the TROIKA dataset. The subject is resting during this segment. **(a)** Label sequence (grey line) and the prediction result by the proposed SQA model (red-dashed line). **(b)** The raw PPG signal is shown in black, and the summation of the decomposed motion rhythm and cardiac component is superimposed in red. The detected R-peaks from the simultaneously recorded ECG are superimposed as vertical grey lines. **(c)** The decomposed cardiac component (black curve), the simultaneously recorded ECG (red curve) and the detected R-peaks (vertical grey lines). **(d)** The decomposed motion rhythm (blue curve) and the x-axis of the accelerometer signal recorded simultaneously (red curve). **(e)** The difference of the PPG signal and the summation of the decomposed motion rhythm and cardiac component. The normalized root mean square error (NRMSE) between the PPG signal and the summation of the decomposed motion rhythm and cardiac component reconstruction is 0.16.

**Figure 68.**
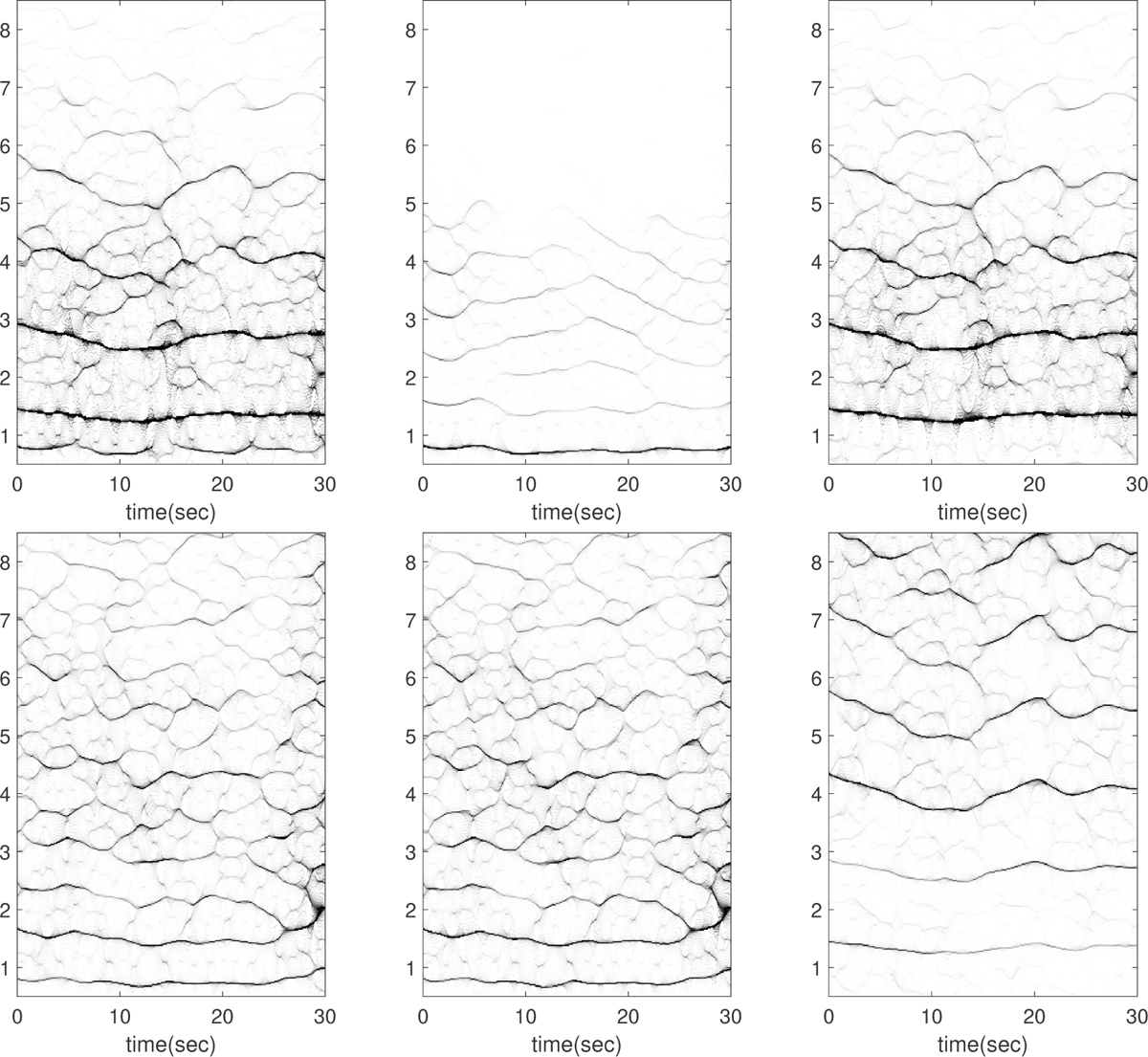
Subject 9 between 0 and 30 seconds in the TROIKA dataset. The subject is resting during this segment. The first row, from left to right: the time-frequency representations (TFRs) of the PPG, the decomposed motion rhythm component, and the PPG after removing the decomposed motion rhythm component. The second row, from left to right: the TFRs of the simultaneously recorded accelerometer magnitude signal, the simultaneously recorded x-axis of the accelerometer signal, and the simultaneously recorded ECG signal. Over this segment, the mean heart rate derived from the simultaneously recorded electrocardiogram (resp. raw PPG and extracted cardiac component) is 1.35 Hz (resp. 1.35 Hz, 1.35 Hz). This is an intriguing case. While the experts’ label suggests that the subject is resting, there is a noticeable regular oscillation with an instantaneous frequency around 0.8 Hz in the accelerometer signal, as well as in the motion rhythm component in the PPG signal, as shown in Figure 69. Despite the lack of detailed information about the recording environment, we are unable to explain, or even hypothesize, the observed signal behavior.

**Figure 69.**
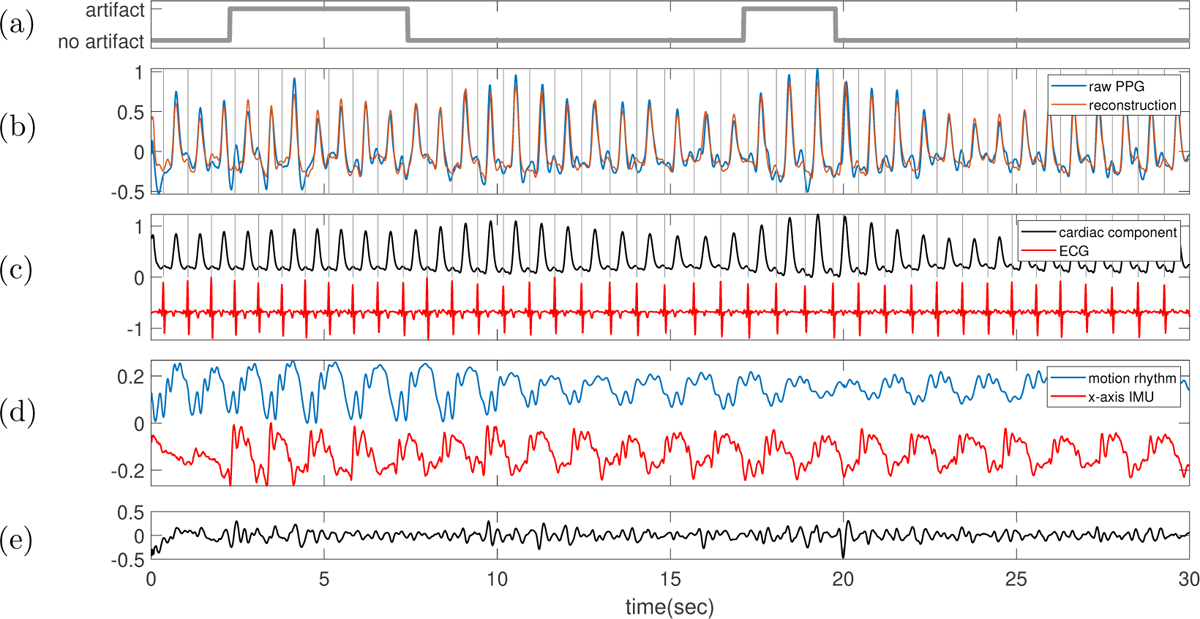
Subject 9 between 0 and 30 seconds in the TROIKA dataset. The subject is resting during this segment. **(a)** Label sequence (grey line) and the prediction result by the proposed SQA model (red-dashed line). **(b)** The raw PPG signal is shown in black, and the summation of the decomposed motion rhythm and cardiac component is superimposed in red. The detected R-peaks from the simultaneously recorded ECG are superimposed as vertical grey lines. **(c)** The decomposed cardiac component (black curve), the simultaneously recorded ECG (red curve) and the detected R-peaks (vertical grey lines). **(d)** The decomposed motion rhythm (blue curve) and the x-axis of the accelerometer signal recorded simultaneously (red curve). **(e)** The difference of the PPG signal and the summation of the decomposed motion rhythm and cardiac component. The normalized root mean square error (NRMSE) between the PPG signal and the summation of the decomposed motion rhythm and cardiac component reconstruction is 0.33.

**Figure 70.**
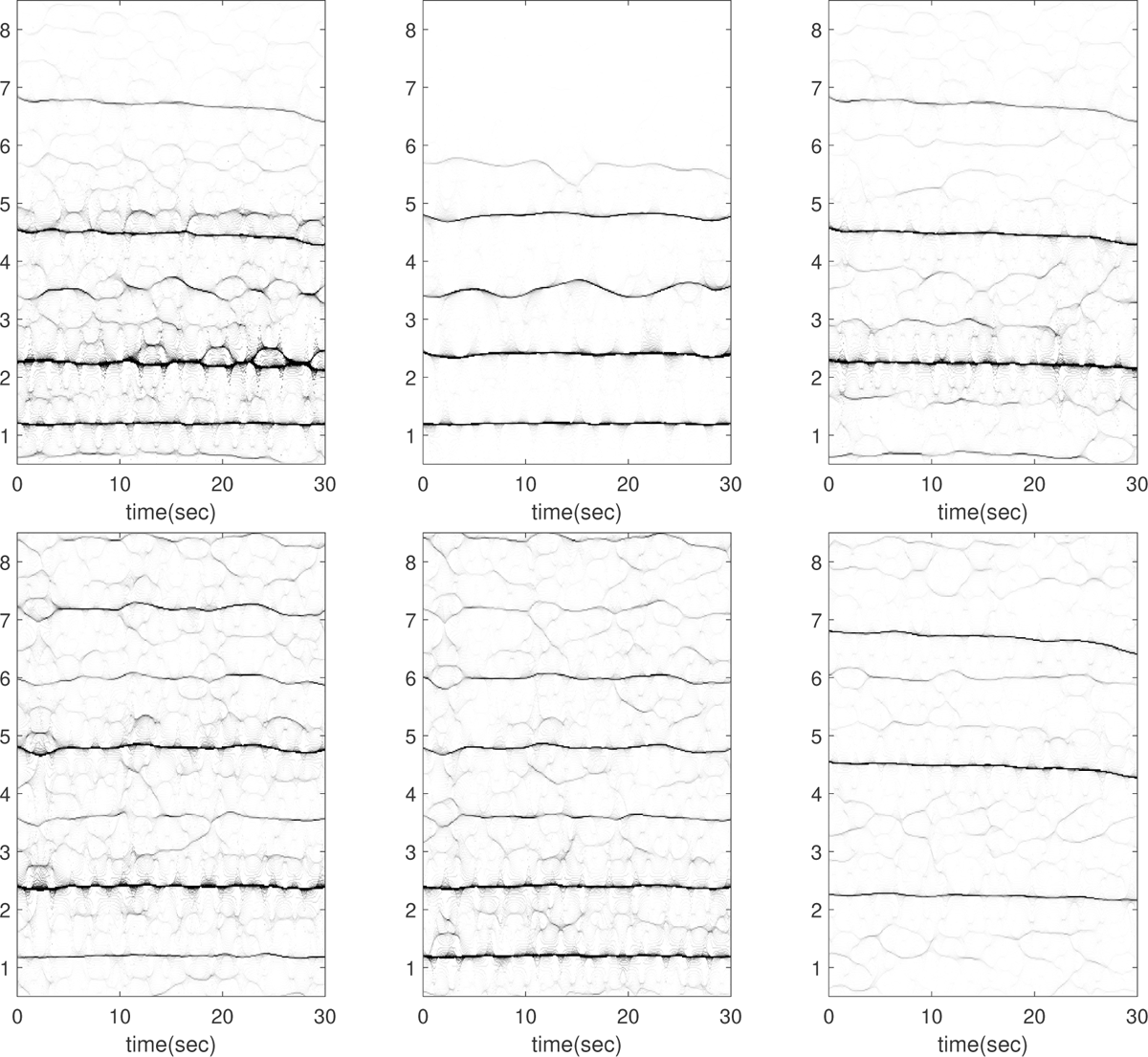
Subject 9 between 180 and 210 seconds in the TROIKA dataset. The subject is running at the speed of 6 kilometer per hour during this segment. The first row, from left to right: the time-frequency representations (TFRs) of the PPG, the decomposed motion rhythm component, and the PPG after removing the decomposed motion rhythm component. The second row, from left to right: the TFRs of the simultaneously recorded accelerometer magnitude signal, the simultaneously recorded x-axis of the accelerometer signal, and the simultaneously recorded ECG signal. Over this segment, the mean heart rate derived from the simultaneously recorded electrocardiogram (resp. raw PPG and extracted cardiac component) is 2.23 Hz (resp. 2.43 Hz, 2.23 Hz).

**Figure 71.**
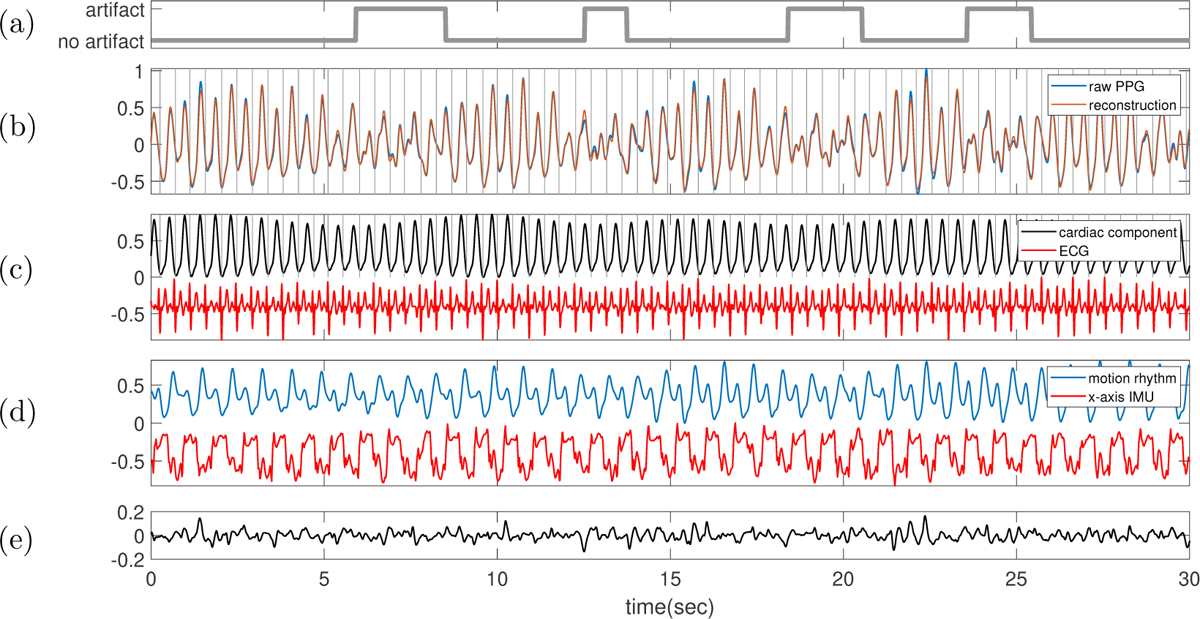
Subject 9 between 180 and 210 seconds in the TROIKA dataset. The subject is running at the speed of 6 kilometer per hour during this segment. **(a)** Label sequence (grey line) and the prediction result by the proposed SQA model (red-dashed line). **(b)** The raw PPG signal is shown in black, and the summation of the decomposed motion rhythm and cardiac component is superimposed in red. The detected R-peaks from the simultaneously recorded ECG are superimposed as vertical grey lines. **(c)** The decomposed cardiac component (black curve), the simultaneously recorded ECG (red curve) and the detected R-peaks (vertical grey lines). **(d)** The decomposed motion rhythm (blue curve) and the x-axis of the accelerometer signal recorded simultaneously (red curve). **(e)** The difference of the PPG signal and the summation of the decomposed motion rhythm and cardiac component. The normalized root mean square error (NRMSE) between the PPG signal and the summation of the decomposed motion rhythm and cardiac component reconstruction is 0.11.

**Figure 72.**
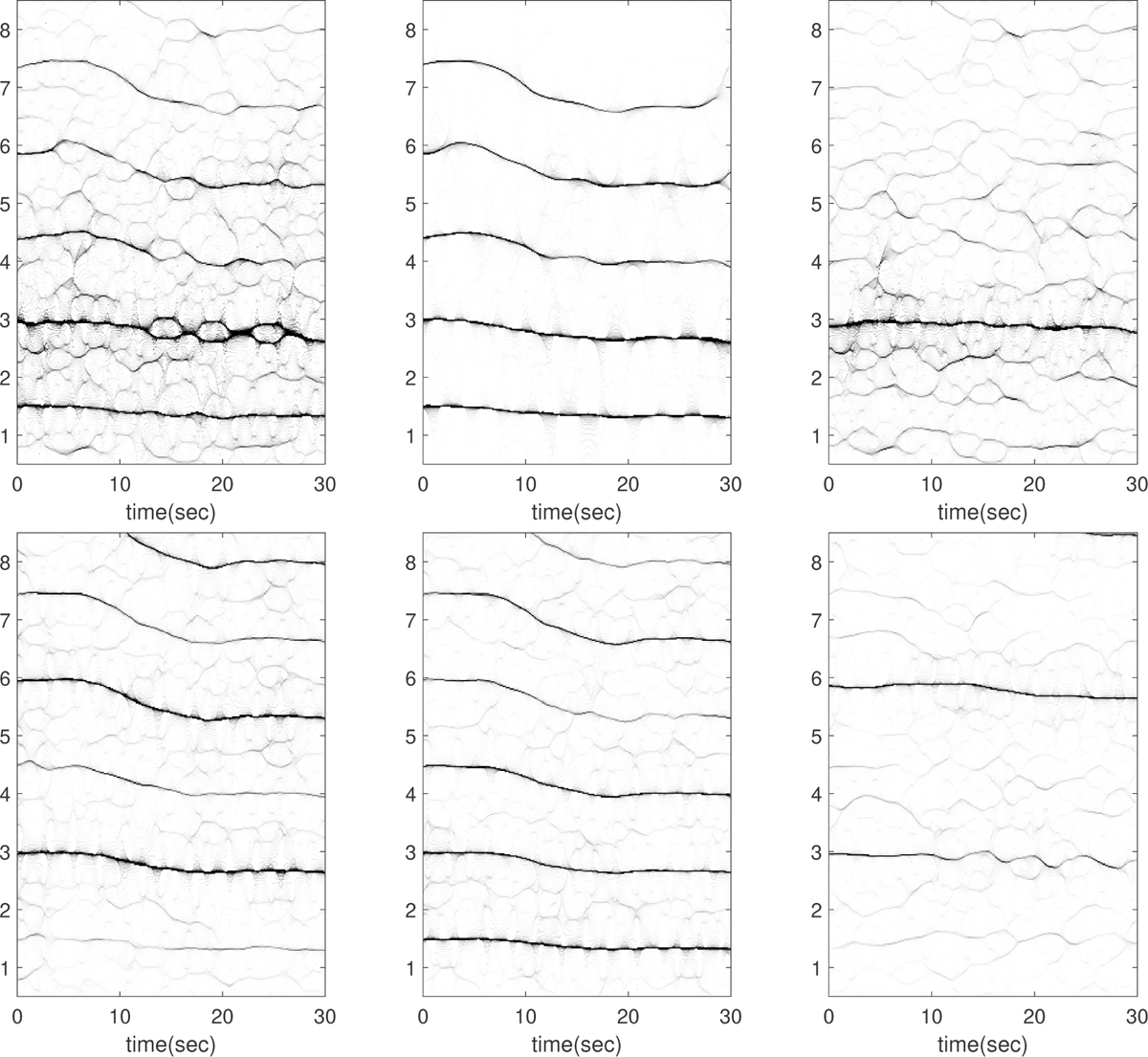
Subject 10 between 270 and 300 seconds in the TROIKA dataset. The subject is resting during this segment. The first row, from left to right: the time-frequency representations (TFRs) of the PPG, the decomposed motion rhythm component, and the PPG after removing the decomposed motion rhythm component. The second row, from left to right: the TFRs of the simultaneously recorded accelerometer magnitude signal, the simultaneously recorded x-axis of the accelerometer signal, and the simultaneously recorded ECG signal. Over this segment, the mean heart rate derived from the simultaneously recorded electrocardiogram (resp. raw PPG and extracted cardiac component) is 2.89 Hz (resp. 3.12 Hz, 2.90 Hz).

**Figure 73.**
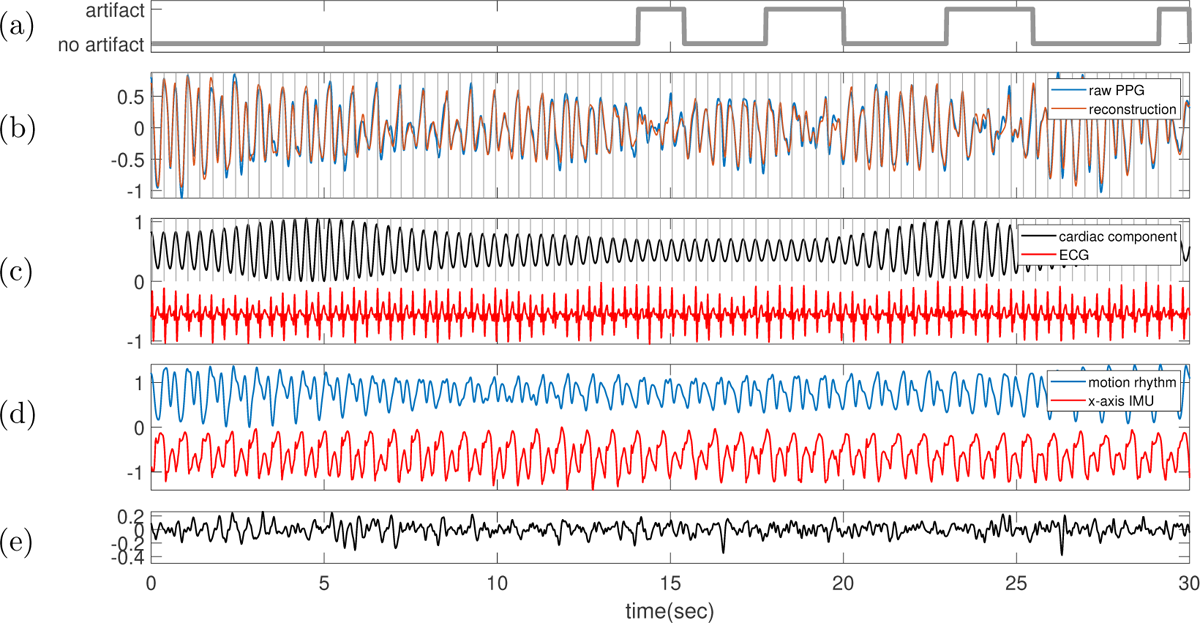
Subject 10 between 270 and 300 seconds in the TROIKA dataset. The subject is resting during this segment. **(a)** Label sequence (grey line) and the prediction result by the proposed SQA model (red-dashed line). **(b)** The raw PPG signal is shown in black, and the summation of the decomposed motion rhythm and cardiac component is superimposed in red. The detected R-peaks from the simultaneously recorded ECG are superimposed as vertical grey lines. **(c)** The decomposed cardiac component (black curve), the simultaneously recorded ECG (red curve) and the detected R-peaks (vertical grey lines). **(d)** The decomposed motion rhythm (blue curve) and the x-axis of the accelerometer signal recorded simultaneously (red curve). **(e)** The difference of the PPG signal and the summation of the decomposed motion rhythm and cardiac component. The normalized root mean square error (NRMSE) between the PPG signal and the summation of the decomposed motion rhythm and cardiac component reconstruction is 0.23.

**Figure 74.**
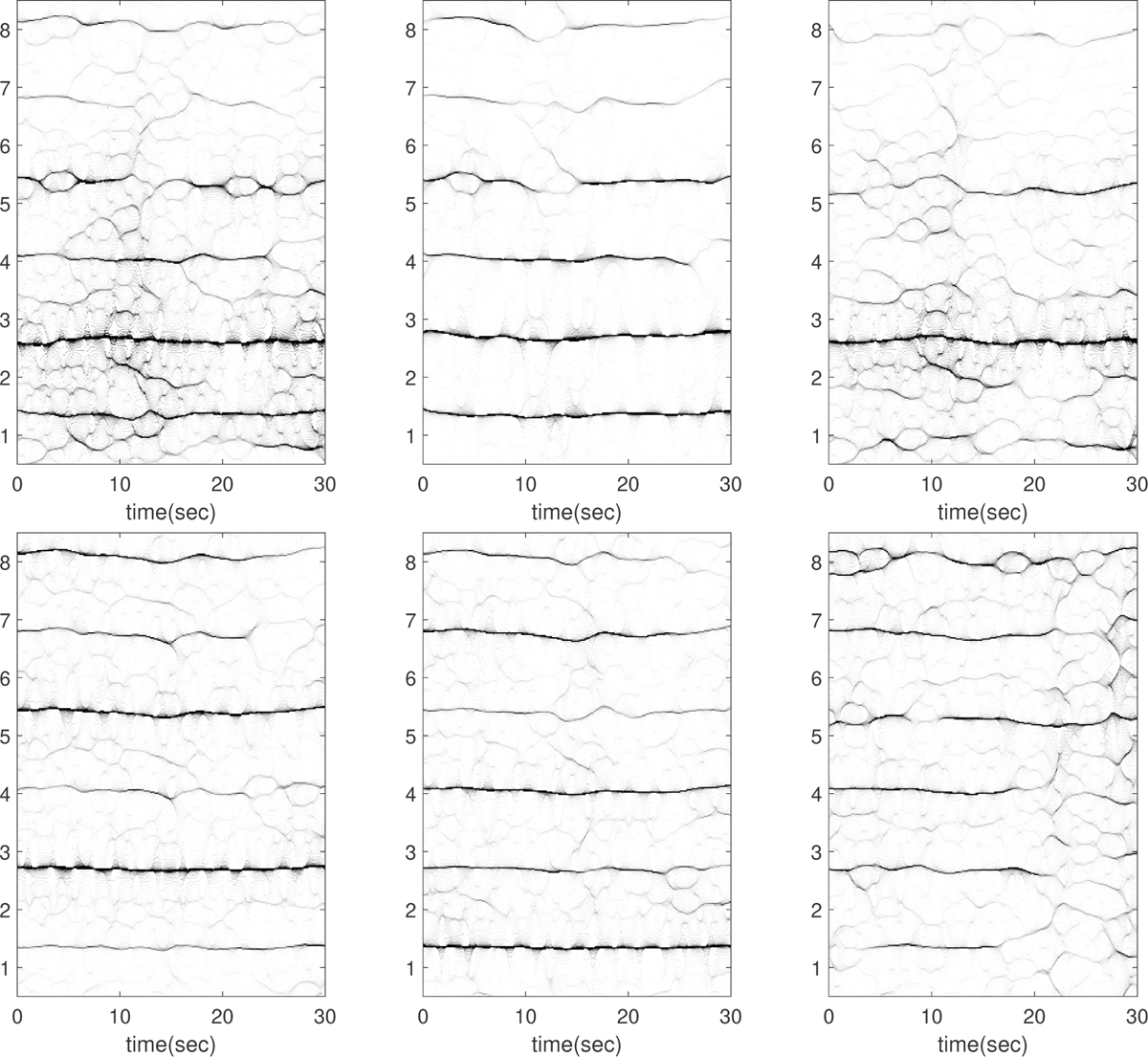
Subject 11 between 180 and 210 seconds in the TROIKA dataset. The subject is running at the speed of 6 kilometer per hour during this segment. The first row, from left to right: the time-frequency representations (TFRs) of the PPG, the decomposed motion rhythm component, and the PPG after removing the decomposed motion rhythm component. The second row, from left to right: the TFRs of the simultaneously recorded accelerometer magnitude signal, the simultaneously recorded x-axis of the accelerometer signal, and the simultaneously recorded ECG signal. Over this segment, the mean heart rate derived from the simultaneously recorded electrocardiogram (resp. raw PPG and extracted cardiac component) is 2.61 Hz (resp. 2.76 Hz, 2.62 Hz). This segment is also noteworthy because the instantaneous frequency (IF) of the second harmonic of the IMU signal closely matches and intertwines with that of the fundamental component of ECG in the first 10 seconds (see the top row of Figure 75); indicating a rough 2:1 coupling between heart rate and motion rhythm. The current algorithm is limited in decomposing such signals. Moreover, the TFR of the ECG reveals a pronounced oscillation that aligns with the motion rhythm, contributing to the ECG’s volatile appearance (see Figure 75(c)).

**Figure 75.**
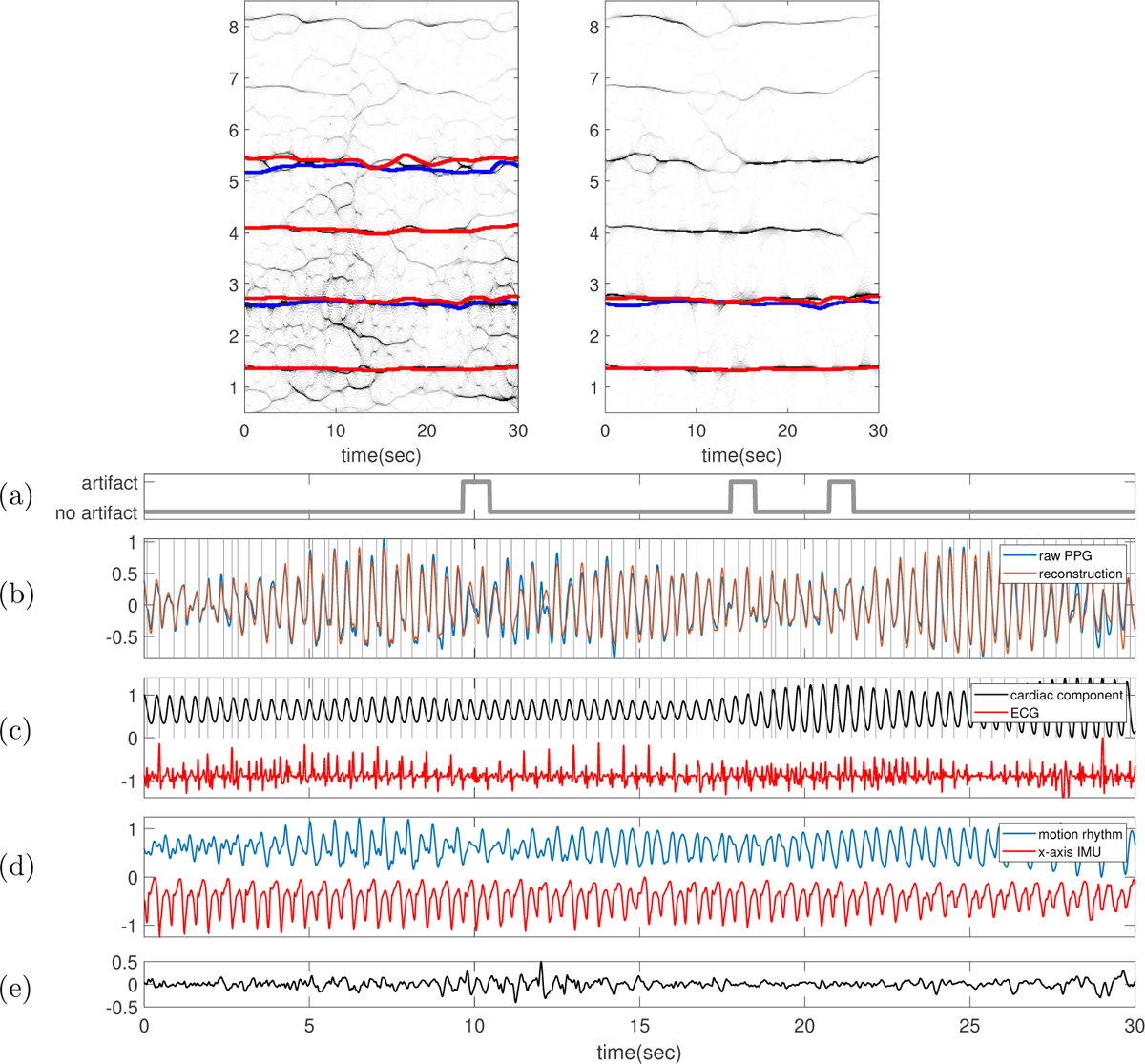
Subject 11 between 180 and 210 seconds in the TROIKA dataset. The subject is running at the speed of 6 kilometer per hour during this segment. In the top row, the TFR of the PPG with the IFs of the first two (four respectively) harmonics of ECG (IMU respectively) superimposed as blue (red respectively) curves are shown on the left-hand side. On the right-hand side, the TFR of the decomposed motion rhythm component with the IF of the first (first two respectively) harmonic of ECG (IMU respectively) superimposed as the blue (red respectively) curve are displayed. The IF of ECG is extracted on the TFR of the rectified and motion rhythm-free ECG; i.e. the motion rhythm is removed by SAMD. **(a)** Label sequence (grey line) and the prediction result by the proposed SQA model (red-dashed line). **(b)** The raw PPG signal is shown in black, and the summation of the decomposed motion rhythm and cardiac component is superimposed in red. The detected R-peaks from the simultaneously recorded ECG are superimposed as vertical grey lines. **(c)** The decomposed cardiac component (black curve), the simultaneously recorded ECG (red curve) and the detected R-peaks (vertical grey lines). **(d)** The decomposed motion rhythm (blue curve) and the x-axis of the accelerometer signal recorded simultaneously (red curve). **(e)** The difference of the PPG signal and the summation of the decomposed motion rhythm and cardiac component. The normalized root mean square error (NRMSE) between the PPG signal and the summation of the decomposed motion rhythm and cardiac component reconstruction is 0.24.

**Figure 76.**
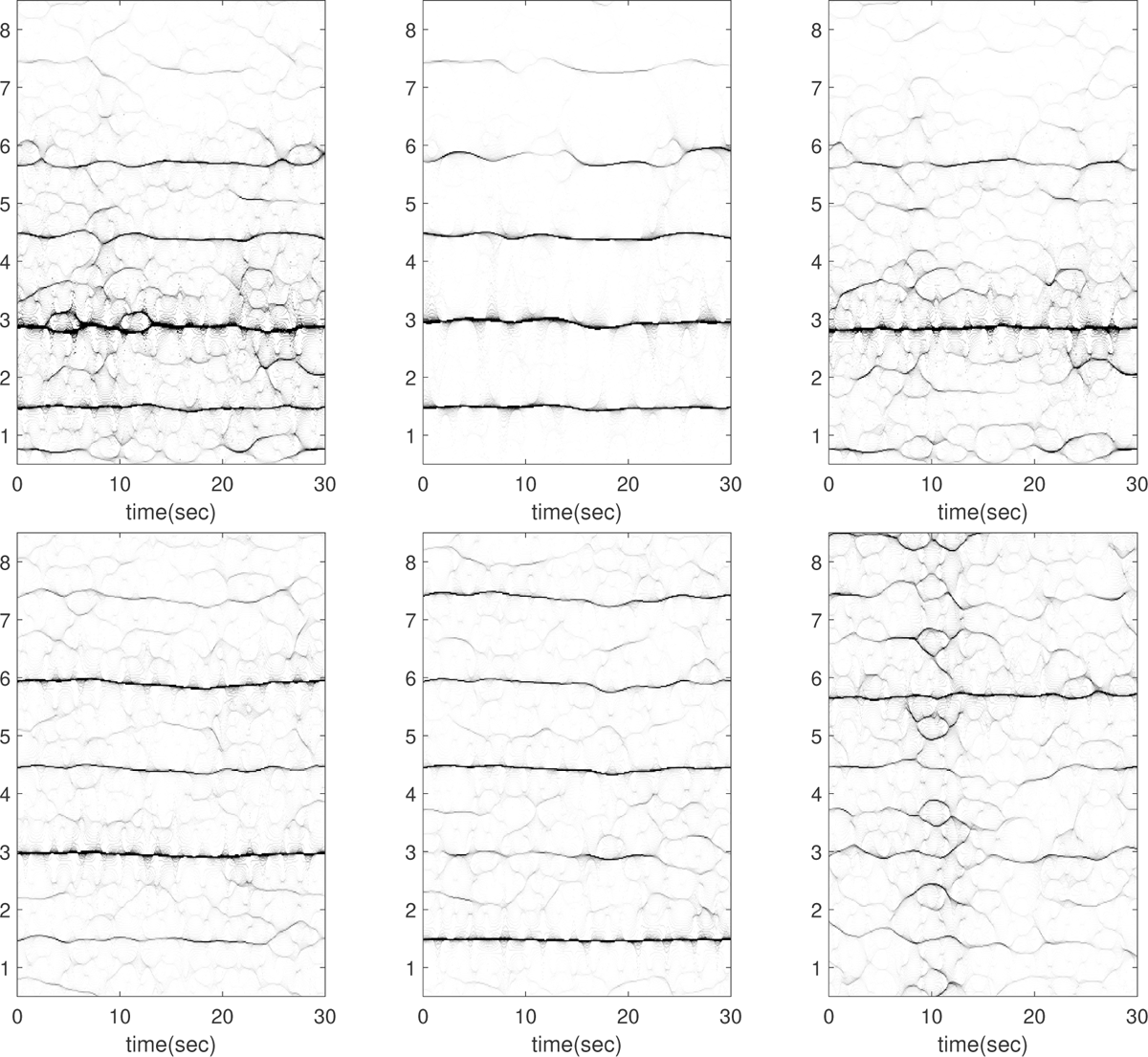
Subject 11 between 240 and 270 seconds in the TROIKA dataset. The subject is running at the speed of 12 kilometer per hour during this segment. The first row, from left to right: the time-frequency representations (TFRs) of the PPG, the decomposed motion rhythm component, and the PPG after removing the decomposed motion rhythm component. The second row, from left to right: the TFRs of the simultaneously recorded accelerometer magnitude signal, the simultaneously recorded x-axis of the accelerometer signal, and the simultaneously recorded ECG signal. Over this segment, the mean heart rate derived from the simultaneously recorded electrocardiogram (resp. raw PPG and extracted cardiac component) is 2.99 Hz (resp. 2.86 Hz, 2.84 Hz). This segment is also noteworthy because the instantaneous frequency (IF) of the second harmonic of the IMU signal closely matches and intertwines with that of the fundamental component of ECG in the first 10 seconds (see the top row of Figure 77); indicating a rough 2:1 coupling between heart rate and motion rhythm. The current algorithm is limited in decomposing such signals.

**Figure 77.**
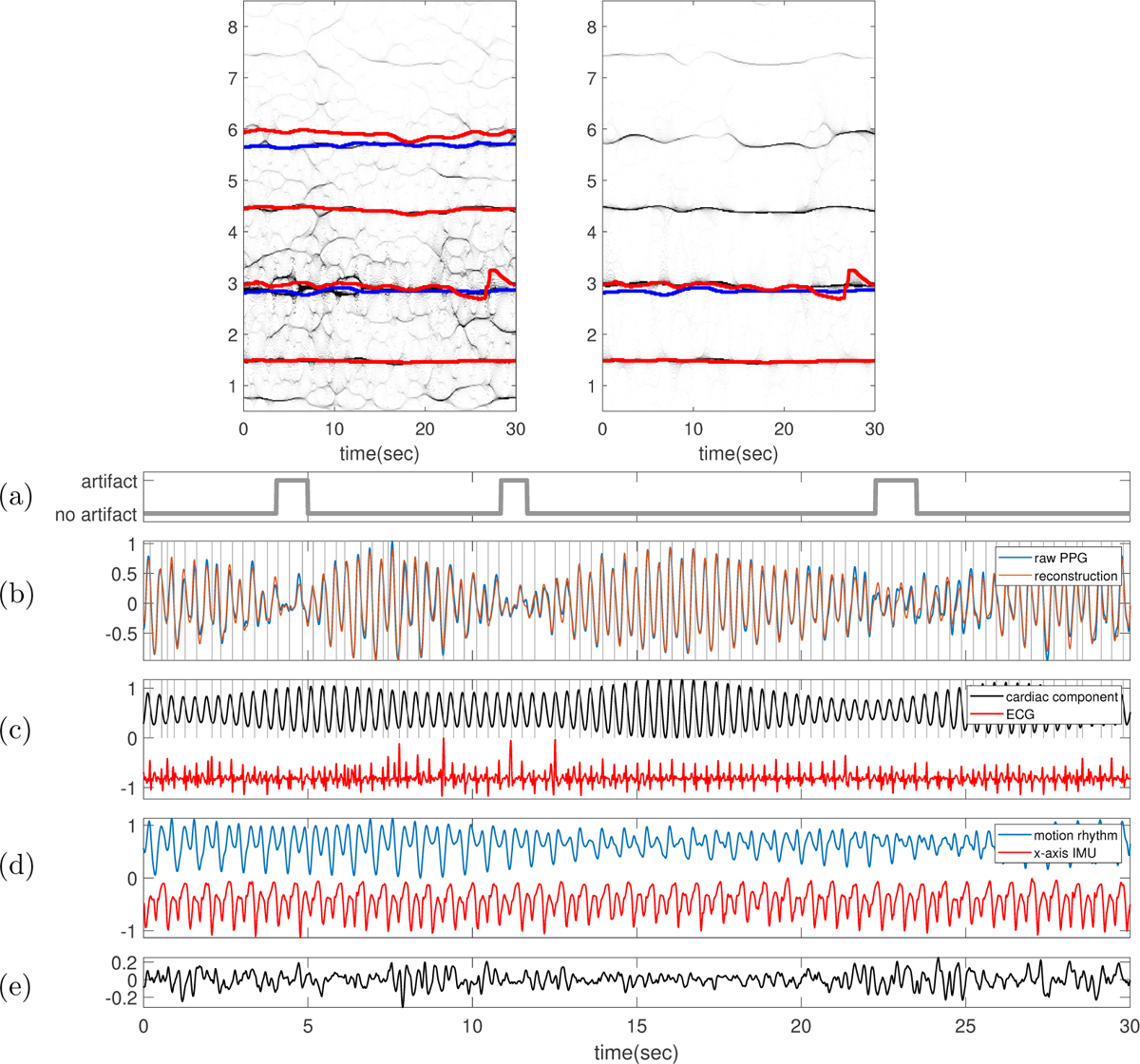
Subject 11 between 240 and 270 seconds in the TROIKA dataset. The subject is running at the speed of 12 kilometer per hour during this segment. In the top row, the TFR of the PPG with the IFs of the first two (four respectively) harmonics of ECG (IMU respectively) superimposed as blue (red respectively) curves are shown on the left-hand side. On the right-hand side, the TFR of the decomposed motion rhythm component with the IF of the first (first two respectively) harmonic of ECG (IMU respectively) superimposed as the blue (red respectively) curve are displayed. The IF of ECG is extracted on the TFR of the rectified and motion rhythm-free ECG; i.e. the motion rhythm is removed by SAMD. **(a)** Label sequence (grey line) and the prediction result by the proposed SQA model (red-dashed line). **(b)** The raw PPG signal is shown in black, and the summation of the decomposed motion rhythm and cardiac component is superimposed in red. The detected R-peaks from the simultaneously recorded ECG are superimposed as vertical grey lines. **(c)** The decomposed cardiac component (black curve), the simultaneously recorded ECG (red curve) and the detected R-peaks (vertical grey lines). **(d)** The decomposed motion rhythm (blue curve) and the x-axis of the accelerometer signal recorded simultaneously (red curve). **(e)** The difference of the PPG signal and the summation of the decomposed motion rhythm and cardiac component. The normalized root mean square error (NRMSE) between the PPG signal and the summation of the decomposed motion rhythm and cardiac component reconstruction is 0.20.

## Footnotes

1 See [9] and the description in [35] for details.

## References

[1] Neng-Tai Chiu, Beau Chuang, Suthawan Anakmeteeprugsa, Kirk H. Shelley, Aymen Awad Alian, and Hau-Tieng Wu. Signal quality assessment of peripheral venous pressure. J. Clin. Monit. Comput., 2023.

[2] Marcelo A Colominas and Hau-Tieng Wu. Decomposing non-stationary signals with time-varying wave-shape functions. IEEE Trans. Signal Process, 69:5094–5104, 2021.

[3] Ricardo Couceiro, P Carvalho, RP Paiva, J Henriques, I Quintal, M Antunes, J Muehlsteff, C Eickholt, C Brinkmeyer, M Kelm, et al. Assessment of cardiovascular function from multi-gaussian fitting of a finger photoplethysmogram. Physiological measurement, 36(9):1801, 2015.

[4] Ingrid Daubechies, Jianfeng Lu, and Hau-Tieng Wu. Synchrosqueezed wavelet transforms: an empirical mode decomposition-like tool. Appl. Comput. Harmon. Anal., 30(2):243–261, 2011.

[5] Nathalie Delprat, Bernard Escudíe, Philippe Guillemain, Richard Kronland-Martinet, Philippe Tchamitchian, and Bruno Torresani. Asymptotic wavelet and gabor analysis: Extraction of instantaneous frequencies. IEEE Trans. Inf. Theory, 38(2):644–664, 1992.

[6] Mohamed Elgendi. Optimal signal quality index for photoplethysmogram signals. Bioengineering, 3, 2016.

[7] P. Flandrin. Time-frequency/time-scale analysis, volume 10. Academic Press Inc., San Diego, 1999.

[8] Ashan Gunarathne, Jeetesh V Patel, Elizabeth A Hughes, and Gregory YH Lip. Measurement of stiffness index by digital volume pulse analysis technique: clinical utility in cardiovascular disease risk stratification. American journal of hypertension, 21(8):866–872, 2008.

[9] Zhicheng Guo, Cheng Ding, Xiao Hu, and Cynthia Rudin. A supervised machine learning semantic segmentation approach for detecting artifacts in plethysmography signals from wearables. Physiological Measurement, 42(12):125003, dec 2021.

[10] B.S. Kim and S.K. Yoo. Motion artifact reduction in photoplethysmography using independent component analysis. IEEE Transactions on Biomedical Engineering, 53(3):566–568, 2006.

[11] Yuriy Kurylyak, Francesco Lamonaca, and Domenico Grimaldi. A neural network-based method for continuous blood pressure estimation from a ppg signal. In 2013 IEEE International instrumentation and measurement technology conference (I2MTC), pages 280–283. IEEE, 2013.

[12] Panicos A Kyriacou and John Allen. Photoplethysmography: Technology, Signal Analysis and Applications. Academic Press, 2021.

[13] Han-Wook Lee, Ju-Won Lee, Won-Geun Jung, and Gun-Ki Lee. The periodic moving average filter for removing motion artifacts from ppg signals. *International Journal of Control*, Automation and Systems, 5, 12 2007.

[14] Chen-Yun Lin, Li Su, and Hau-Tieng Wu. Wave-shape function analysis–when cepstrum meets timefrequency analysis. J. Fourier Anal. Appl., 24(2):451–505, 2018.

[15] Sheng Lu, He Zhao, Kihwan Ju, Kunson Shin, Myoungho Lee, Kirk Shelley, and Ki H Chon. Can photoplethysmography variability serve as an alternative approach to obtain heart rate variability information? Journal of clinical monitoring and computing, 22:23–29, 2008.

[16] Elisa Mejia-Mejia, John Allen, Karthik Budidha, Chadi El-Hajj, Panicos A. Kyriacou, and Peter H. Charlton. 4 - photoplethysmography signal processing and synthesis. In John Allen and Panicos Kyriacou, editors, Photoplethysmography, pages 69–146. Academic Press, 2022.

[17] Sandrine C Millasseau, Ronan P Kelly, James M Ritter, and Philip J Chowienczyk. The vascular impact of aging andvasoactive drugs: Comparison of two digital volume pulse measurements. American journal of hypertension, 16(6):467–472, 2003.

[18] Serena Moscato, Stella Lo Giudice, Giulia Massaro, and Lorenzo Chiari. Wrist photoplethysmography signal quality assessment for reliable heart rate estimate and morphological analysis. Sensors, 22(15), 2022.

[19] Patrick Mullan, Christoph M. Kanzler, Benedikt Lorch, Lea Schroeder, Ludwig Winkler, Larissa Laich, Frederik Riedel, Robert Richer, Christoph Luckner, Heike Leutheuser, Bjoern M. Eskofier, and Cristian Pasluosta. Unobtrusive heart rate estimation during physical exercise using photoplethysmographic and acceleration data. In 2015 37th Annual International Conference of the IEEE Engineering in Medicine and Biology Society (EMBC), pages 6114–6117, 2015.

[20] Emad Kasaeyan Naeini, Iman Azimi, Amir M. Rahmani, Pasi Liljeberg, and Nikil Dutt. A real-time ppg quality assessment approach for healthcare internet-of-things. Procedia Computer Science, 151:551–558, 2019. The 10th International Conference on Ambient Systems, Networks and Technologies (ANT 2019)/ The 2nd International Conference on Emerging Data and Industry 4.0 (EDI40 2019) / Affiliated Workshops.

[21] Thomas Oberlin, Sylvain Meignen, and Vaĺerie Perrier. Second-order synchrosqueezing transform or invertible reassignment? towards ideal time-frequency representations. IEEE Trans. Signal Process, 63(5):1335–1344, 2015.

[22] Michael J Oppenheim and Dean F Sittig. An innovative dicrotic notch detection algorithm which combines rule-based logic with digital signal processing techniques. Computers and biomedical research, 28(2):154–170, 1995.

[23] Christina Orphanidou. Signal Quality Assessment in Physiological Monitoring: State of the Art and Practical Considerations. 01 2018.

[24] Christina Orphanidou, Timothy Bonnici, Peter Charlton, David Clifton, David Vallance, and Lionel Tarassenko. Signal-quality indices for the electrocardiogram and photoplethysmogram: Derivation and applications to wireless monitoring. IEEE Journal of Biomedical and Health Informatics, 19(3):832– 838, 2015.

[25] Birutė Paliakaitė, Andrius Petrėnas, Andrius Solšenko, and Vaidotas Marozas. Modeling of artifacts in the wrist photoplethysmogram: Application to the detection of life-threatening arrhythmias. Biomedical Signal Processing and Control, 66:102421, 2021.

[26] Fulai Peng, Hongyun Liu, and Weidong Wang. A comb filter based signal processing method to effectively reduce motion artifacts from photoplethysmographic signals. Physiological Measurement, 36(10):2159, sep 2015.

[27] S. M. A. Salehizadeh, Duy K. Dao, Jo Woon Chong, David McManus, Chad Darling, Yitzhak Mendelson, and Ki H. Chon. Photoplethysmograph signal reconstruction based on a novel motion artifact detection-reduction approach. part ii: Motion and noise artifact removal. Annals of Biomedical Engineering, 42(11):2251–2263, Nov 2014.

[28] Jean Schmith, Carolina Kelsch, Beatriz Cappelozza Cunha, Lucio Rene Prade, Eduardo Augusto Martins, Armando Leopoldo Keller, and Rodrigo Marques de Figueiredo. Photoplethysmography signal quality assessment using attractor reconstruction analysis. Biomedical Signal Processing and Control, 86:105142, 2023.

[29] Kirk H Shelley. Photoplethysmography: beyond the calculation of arterial oxygen saturation and heart rate. Anesthesia & Analgesia, 105(6):S31–S36, 2007.

[30] Kirk H Shelley, Doris Tamai, Denis Jablonka, Michael Gesquiere, Robert G Stout, and David G Silverman. The effect of venous pulsation on the forehead pulse oximeter wave form as a possible source of error in spo2 calculation. Anesthesia & Analgesia, 100(3):743–747, 2005.

[31] Hangsik Shin. Deep convolutional neural network-based signal quality assessment for photoplethysmogram. Computers in Biology and Medicine, 145:105430, 2022.

[32] Yan-Wei Su, Gi-Ren Liu, Yuan-Chung Sheu, and Hau-Tieng Wu. Ridge detection for nonstationary multicomponent signals with time-varying wave-shape functions and its applications. *arXiv preprint arXiv:2309.06673*, 2023.

[33] J Abdul Sukor, S J Redmond, and N H Lovell. Signal quality measures for pulse oximetry through waveform morphology analysis. Physiological Measurement, 32(3):369, feb 2011.

[34] H.-T. Wu. Instantaneous frequency and wave shape functions (I). Appl. Comput. Harmon. Anal., 35:181–199, 2013.

[35] Zhilin Zhang, Zhouyue Pi, and Benyuan Liu. Troika: A general framework for heart rate monitoring using wrist-type photoplethysmographic signals during intensive physical exercise. IEEE Transactions on Biomedical Engineering, 62(2):522–531, 2015.

